# Glymphatic flow resistance in humans measured by multifrequency impedance dispersion

**DOI:** 10.1101/2024.01.06.24300933

**Authors:** Paul Dagum, Laurent Giovangrandi, Swati Rane Levendovszky, Jake J. Winebaum, Tarandeep Singh, Yeilim Cho, Robert M. Kaplan, Michael S. Jaffee, Miranda M. Lim, Carla Vandeweerd, Jeffrey J. Iliff

## Abstract

Glymphatic function in animal models supports the clearance of brain proteins whose mis-aggregation is implicated in neurodegenerative conditions including Alzheimer’s and Parkinson’s disease. The measurement of glymphatic function in the human brain has been elusive, limiting its potential in translational research. Here we described a non-invasive multimodal device for the first-in-human continuous measurement of sleep-active glymphatic flow resistance using repeated electrical impedance spectroscopy measurements through two separate clinical validation studies. Device measurements successfully (i) paralleled sleep-associated changes in extracellular volume that regulate glymphatic function, (ii) replicated preclinical findings showing glymphatic function is increased with increasing sleep EEG delta power, and is decreased with increasing sleep EEG beta power and heart rate, and (iii) predicted glymphatic solute exchange measured by contrast-enhanced MRI. The present investigational device permits the continuous and time-resolved assessment of glymphatic flow resistance in naturalistic settings necessary to determine the contribution of glymphatic impairment to risk and progression of Alzheimer’s disease, and to enable target-engagement studies that modulate glymphatic function in humans.

## INTRODUCTION

The glymphatic system is a brain-wide network of perivascular pathways along which the cerebrospinal fluid (CSF) surrounding the brain exchanges with brain interstitial fluid, supporting nutrient distribution and waste clearance (**Figure 1a-b**). In animal models, glymphatic function supports the clearance of amyloid beta^1,2^, tau^3–5^, and alpha synuclein^6,7^, whose mis-aggregation is implicated in the pathogenesis of neurodegenerative conditions including Alzheimer’s and Parkinson’s disease. Glymphatic function is further postulated to be a mechanism for volume transmission of the neurotransmitters acetylcholine, serotonin and norepinephrine, and of neurohormones regulating brain-body homeostasis^8^. While this system is foundational to brain health and disease, measurement of glymphatic function in the human brain, principally by magnetic resonance imaging (MRI), is conducted in only a handful of academic neuroimaging centers^9^. Without a consistent, reproducible and accessible way of measuring this important neurobiology in humans, the promise of developing therapeutic interventions that target glymphatic function to treat and prevent neurological and psychiatric diseases remains elusive.

**Figure 1.**
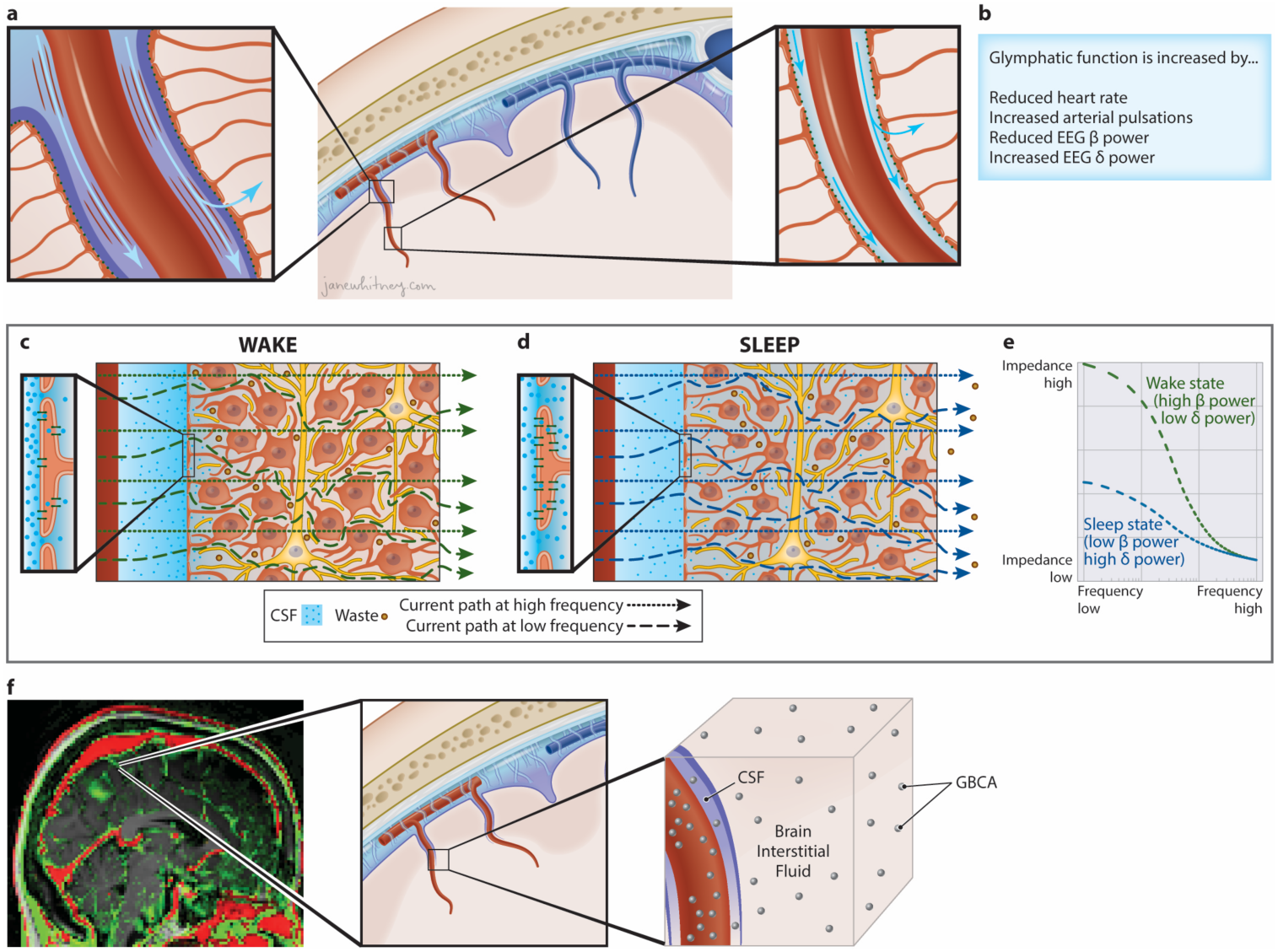
Detection of sleep-wake changes in glymphatic flow resistance by impedance spectroscopy. **a**, Glymphatic function involves the influx of cerebrospinal fluid (CSF) influx along perivascular spaces (PVSs) surrounding penetrating arteries (center). Glymphatic flow is driven by arterial pulsations and vasomotor oscillations (right). Astrocytic endfoot processes create the barrier between the PVS and the brain parenchyma, with gaps allowing CSF to exchange through the brain interstitial space. The endfoot processes are studded with perivascular aquaporin-4 water channels that also enable water to move into the astrocytes and out to the interstitial space. **b,** Data from physiological studies in rodents suggest that glymphatic function is increased in conditions of reduced heart rate, increased vasomotor pulsations, reduced EEG beta power, and increased EEG delta power (Hablitz et al. 2019). **c,** The interstitial space is dynamically regulated. Under conditions of low EEG delta power and high EEG beta power common to the awake state, it is narrow and tortuous, forming a high-resistance pathway that suppresses glymphatic function. When alternating current is injected into the brain parenchyma, at low frequencies the current cannot penetrate the capacitive cell membranes and its propagation depends primarily on the resistive pathway of the interstitial fluid. At high current frequencies, the current readily penetrates the capacitive cell membranes and its propagation depends on the resistance of the total tissue volume. This frequency difference is the β-dielectric dispersion of the underlying tissue. If the intracranial volume is constant, a change in the dielectric dispersion reflects a change in the low-frequency resistance pathway of the interstitial fluid. **d,** Under high EEG delta power and low EEG beta power common to the sleep state, fluid shifts from the intracellular compartment into the interstitial space, enhancing glymphatic function by ∼60% in rodent studies (Xie et al. 2013). This widening of interstitial pathways reduces the current resistance at low-frequencies which reduces the measured dielectric dispersion. **e**, An impedance-frequency graph shows the change in dielectric dispersion between the two sleep/wake EEG states. The change in glymphatic flow resistance between these two states is inversely proportional to the relative change in the dielectric dispersion. **f,** Contrast-enhanced MRI following intravenous gadolinium-based contrast agent (GBCA) injection shows vascular regions that enhance immediately (t=30 min) upon GBCA injection (red), and CSF and brain parenchymal regions that enhance late (t=3 hrs) after leakage of GBCA first into the CSF and thence into the brain interstitium (green). A given MRI voxel includes GBCA within the blood, CSF and brain interstitial fluid compartments.

Studies in rodents have demonstrated that glymphatic function is more rapid during sleep compared to the awake state^2,10^. During periods of wakefulness, the interstitial space is narrow and tortuous, forming a high-resistance pathway that suppresses glymphatic flow, while during sleep fluid shifts from the intracellular compartment into the extracellular (interstitial) space, widening these pathways, and enhancing glymphatic function by ∼60% (**Figure 1c-d**).

Measuring glymphatic function across a range of physiological conditions demonstrated that faster glymphatic exchange is associated with increased electroencephalogram (EEG) delta band power, decreased EEG beta power, and reduced heart rate^11^. Studies utilizing contrast-enhanced (CE)-MRI to measure solute exchange between the CSF and brain interstitial compartments have confirmed in human participants more rapid glymphatic function during sleep^12^, although the poor temporal resolution of this approach^13–15^ has not permitted the relationship between EEG or cardiovascular features and glymphatic function to be defined in the human brain.

Electrical impedance spectroscopy (EIS) is widely used in medicine for body composition analysis and for monitoring cellular changes in settings including tumors and edema^16–19^. When alternating current is injected into tissue, at low frequencies the voltage drop depends primarily on the resistive pathway of the extracellular (interstitial) fluid, while at high frequencies cell membranes behave like capacitors allowing current to also pass relatively freely through the intracellular space (**Figure 1c-e**). This frequency dependence is the β-dielectric dispersion of the underlying tissue and its repeated measurement permits the detection of shifts in the extracellular and intracellular distribution of water within tissue^17,20^.

We propose that the biophysics of EIS can be leveraged to measure the fluid shift that occurs with changes in sleep-active glymphatic function. Because intracranial volume is constant, any change in this frequency dependence across repeated EIS measures reflects changes in the resistance of the extracellular space, which is inversely proportional to the extracellular fluid volume (**Figure 1c-e**). Within the brain, the extracellular compartment includes the brain interstitial space (∼16% of total volume) and a combination of CSF (∼10%) and blood plasma (∼6%)^21^. Small diurnal fluctuations occur in blood volume and in the CSF compartments, however these changes are small and offset one another across physiological states^22,23^. In contrast, sleep-wake differences in glymphatic function are associated with much larger shifts in the intracellular and particularly interstitial volumes^2^. Thus, sleep-wake changes in EIS measures will be sensitized to changes in brain intracellular and interstitial volume, and EIS-based measures of parenchymal resistance (R_P_) may represent a novel approach to track sleep-active glymphatic function.

We developed a wearable investigational device (**Figure 2a-f**) that overcomes several technical challenges surrounding the implementation of EIS as a measure of glymphatic flow resistance. To define the dynamic relationship between R_P_, sleep EEG features, and cardiovascular parameters through the course of an overnight period, EIS measurements must be made with high accuracy and temporal resolution. The investigational device performs measurements at frequencies between 1kHz and 256kHz and has a ∼3% measurement error in R_P_ (**Supplemental 4)** enabling it to detect small changes in dispersion between repeated measures. It is important to ensure that device measurements are not affected by changes in electrode impedances that can occur over time, including those caused by the effects of motion on the electrode-skin interface. A non-traditional four-electrode impedance configuration was used with multiple excitation and sense configurations in combination with low-impedance, non-polarizable electrodes. This ensured high-fidelity measurements including at low frequencies due to the reduced sensitivity to motion artifacts, low noise, and stability of the skin-gel-electrode interface. A signal processing approach enabled identification of signal from noise using the real and imaginary parts of the impedance to establish if the measured response is related to the excitation signal or has been distorted by artifact. The device further integrated shielded cables to reduce effects of electromagnetic interferences, both external and internal to the device, and the design minimized stray capacitances affecting higher frequencies. The device is described in more detail in **Methods** and **Supplemental Information** including **Supplemental Figure 2, Supplemental Figure 3** and **Supplemental Tables 7 – 12**. We note that glymphatic flow is determined by the parenchymal resistance to flow and the motive force driving CSF along the perivascular spaces (PVS) into the interstitial compartment. The resistance measured by the device is transcranial with a high temporal resolution of changes in global intracellular to interstitial fluid shifts.

**Figure 2.**
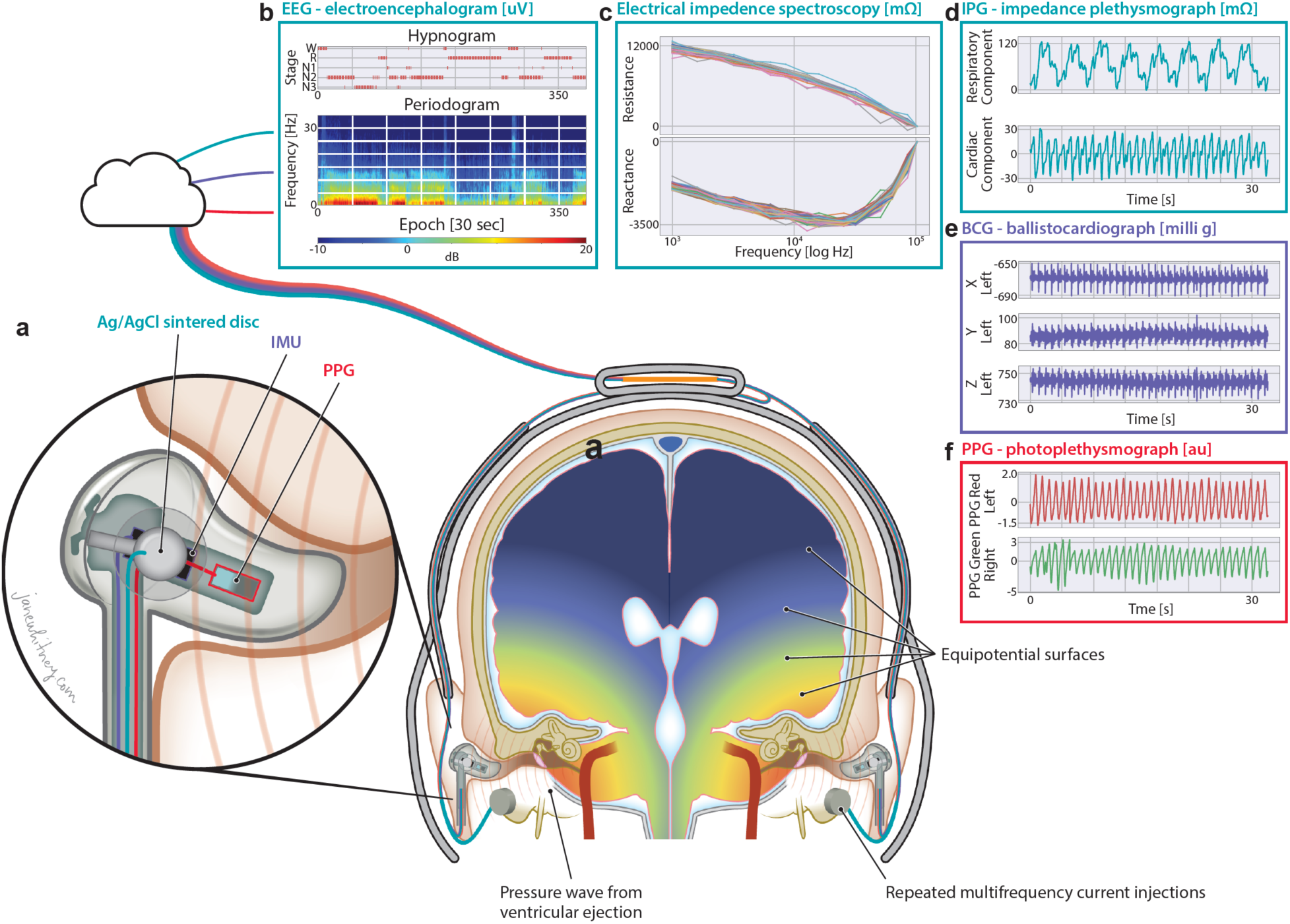
Technical schematic of investigational device and its output signals. Data for this study was acquired with a non-invasive multimodal skin-interfaced wireless device for continuous measurement of brain parenchymal resistance using repeated electrical impedance spectroscopy EIS time-multiplexed with EEG and cardiovascular measurements. EEG differential measurements were made between the two in-ear electrodes and a left mastoid was used to drive the common mode. These electrodes were shared with the EIS and the two measurements were time-multiplexed with a pair of analog multiplexers that decoupled EEG and EIS. The transcranial multi-frequency alternating current injections created an electric field through the brain and orthogonal equipotential surfaces. **a**, Each in-ear electronics housed PPG and IMU sensors. **b**, Data from the sensors were stored on the device’s FLASH memory and transferred via a USB port to a cloud signal processing pipeline for offline analysis. Data from a participant visit is shown, which includes the device EEG periodogram and hypnogram. **c**, The EIS resistance and reactance frequency plots for this participant reveal an approximate 2000 mOhm change in the dielectric dispersion during the sleep visit. **d**, The IPG respiratory and cardiac impedance variations are 120 mn and 50 mn, respectively. **e**, Cardiac ejection of blood is detected by the in-ear IMUs in the ballistocardiogram. The J peak of the ballistocardiogram marks the aortic valve opening. **f**, Time delay between the ballistocardiogram and in-ear PPG measures the pulse-transit time (PTT) to the that ear.

## RESULTS

Data were acquired using the investigational medical device developed by Applied Cognition Inc^24,25^. Transcranial impedance dispersion is estimated from each EIS measure using a dielectric relaxation model. In addition to EIS measurements, the device measures EEG, photoplethysmogram (PPG), impedance plethysmogram (IPG) and head motion (**Figure 2a-f**). We propose that the change in transcranial impedance dispersion, which we will henceforth term parenchymal resistance R_P_ (*R_P_* = *R*_0_ − *R*_∞_), reflects shifts in brain intracellular and interstitial compartment fluid volumes that underlie changes in glymphatic function (**Figure 1c-e**). **Supplemental 3** contains an intuitive justification of the physiological connection between EIS change and changes in glymphatic function. To test this, we evaluated whether R_P_ measured with this device in human participants aligned with the central features of glymphatic function described in rodent models^2,11^.

### Study Design

We conducted two clinical studies (denoted as “Benchmarking Study” and “Replication Study”) to validate the performance of the device’s R_P_ measure as a surrogate of glymphatic flow resistance during sleep and wake (**Figure 3**, top). Both studies were cross-over trials where participants wearing the investigational device were subjected to one night of natural sleep and one night spent awake, separated by two or more weeks. The Benchmarking Study was conducted in The Villages® community, an active-lifestyle senior living community in Central Florida where the University of Florida maintains a satellite academic research center, The University of Florida Health Precision Health Research Center (UF Health PHRC). The Benchmarking Study was designed to define the effect of sleep on overnight R_P_ measurements, as well as the relationship between R_P_ and glymphatic function. It included overnight device recordings, overnight gold-standard polysomnography (PSG), and morning contrast-enhanced magnetic resonance imaging (CE-MRI) following intravenous gadolinium-based contrast agent (GBCA) administration as a measure of glymphatic function. During the morning period, following pre-contrast MRI scanning, GBCA injection, and immediate post-contrast MRI scanning (totaling 2.5 hours spent awake), participants that were sleep deprived overnight were provided a 1.5 hr sleep opportunity prior to the final MRI scan, while those that slept normally overnight were kept awake during the 1.5 hr period (**Figure 3**, top). We evaluated whether, as predicted from prior rodent studies on the sleep-modulation of glymphatic function^2,11^, **a)** R_P_ is reduced in sleeping compared to awake participants, **b)** whether reduced R_P_ is associated with more rapid CSF-brain interstitial solute exchange measured by CE-MRI, **c)** whether R_P_ is associated with increasing sleep EEG delta power and decreasing sleep EEG beta power, and **d)** whether reduced R_P_ is associated with decreasing heart rate. The Replication Study conducted at the University of Washington did not include overnight PSG or morning CE-MRI and had as a primary outcome confirming the effect of sleep state on device-measured R_P_, and secondary outcomes confirming the associations between R_P_ and sleep stages, heart rate, and EEG spectral band power. As noted in the detailed study description in the Methods section, these studies also evaluated novel non-contrast MRI measures of glymphatic function, sleep-sensitive changes in cognitive function, and sleep-sensitive changes in plasma Alzheimer’s disease biomarkers across a range of physiological sleep conditions. Those findings will be reported elsewhere.

**Figure 3.**
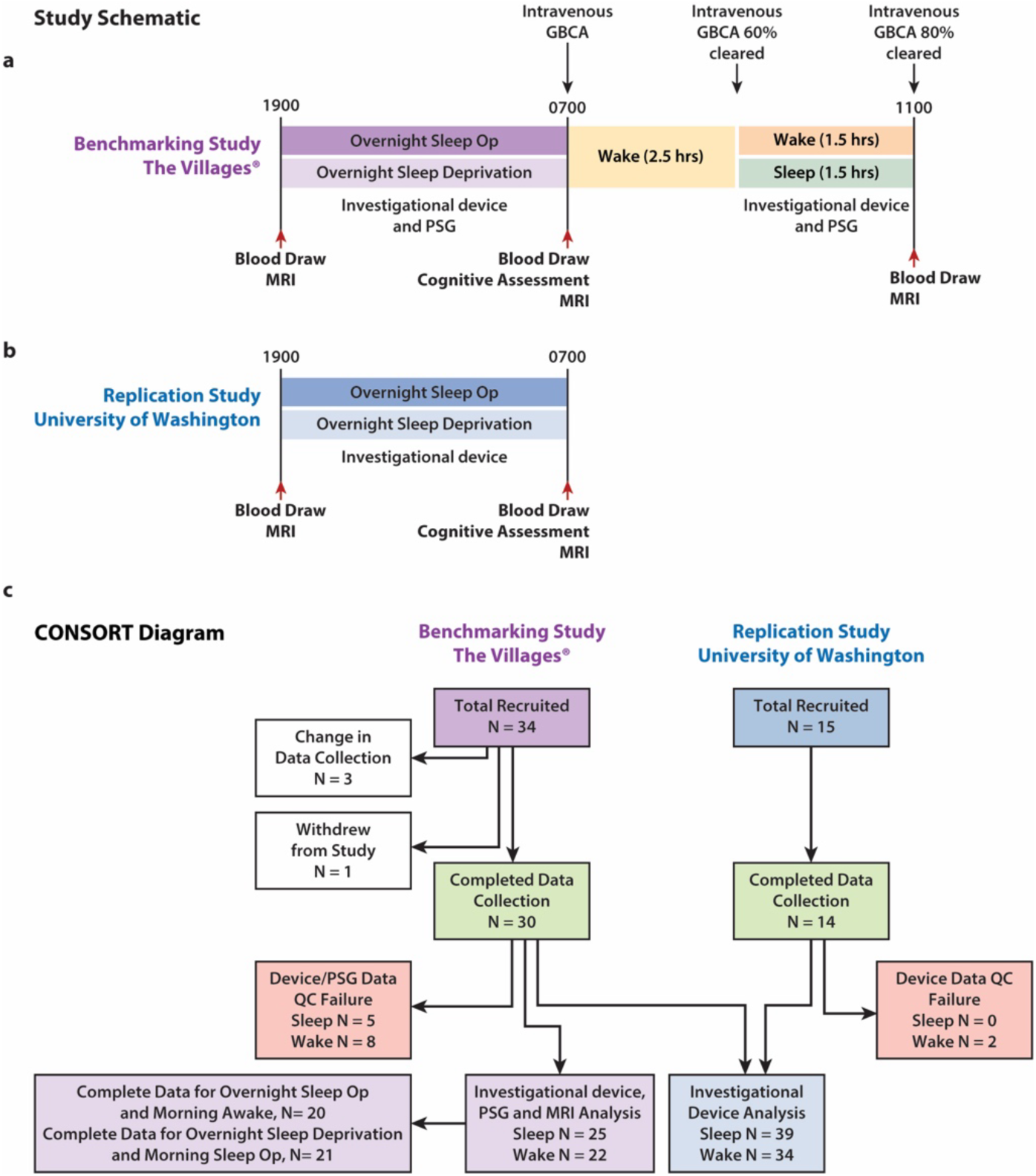
Study schematic and CONSORT diagram for Benchmarking Study and Replication Study. **a**, The Benchmarking Study conducted at The Villages® was designed to define the relationship between parenchymal resistance (R_P_) and glymphatic function. Reported here are the overnight and morning device recordings, overnight and morning gold-standard polysomnography (PSG), and morning contrast-enhanced magnetic resonance imaging (CE-MRI) following intravenous gadolinium-based contrast agent (GBCA) administration as a measure of glymphatic function. Primary and secondary outcome data not reported here included blood analysis of amyloid β and tau levels, cognitive assessment, and non-contrast MRI. **b**, The Replication Study conducted at the University of Washington had a primary outcome of confirming the effect of sleep state on device-measured R_P_, and secondary outcomes of confirming the associations between R_P_ and sleep stages, heart rate, and EEG spectral band power. The Replication Study also included secondary outcomes on blood analyses for amyloid β and tau levels, cognitive assessment and non-contrast MRI. **c**, The Benchmarking Study enrolled 34 participants of which 30 completed both visits. Three were censored due to changes in device data collection and sensor locations. One withdrew following the first MRI scan. Of the 30 that completed the study, 5 overnight sleep visits and 8 overnight wake visits failed the data quality control (QC) criteria to provide sufficient artifact free data to yield results. This resulted in 25 sleep and 22 wake device, PSG and MRI complete data sets. Of these, 20 sleep and 21 wake device participants had complete data from both the overnight and morning sleep/wake periods. The Replication Study enrolled 14 participants. All 14 completed the study, of which 3 wake visits failed the data QC criteria and no sleep visit failed. (sleep op, sleep opportunity)

### Inclusion and Exclusion Criteria

All studies were performed between October 2022 – June 2023 and were reviewed and approved by University of Florida Institutional Review Board (IRB No. 202201364, Benchmarking Study) and Western Institutional Review Board (IRB No. 20225818, Replication Study). The studies have been registered at ClinicalTrials.gov (https://clinicaltrials.gov/study/NCT06222385 and https://clinicaltrials.gov/study/NCT06060054). Written informed consent was obtained from all study participants during a screening visit, prior to any study activities. Studies were carried out in accordance with the principles of the Belmont Report. The Benchmarking Study enrolled 34 healthy participants 56-66 years of age. The Replication Study enrolled 14 healthy participants 49-63 years of age. Participants were excluded if they had cognitive impairment or clinical depression. Cognitive impairment was assessed using the Montreal Cognitive Assessment^26^ (MoCA, 28.1 +/− 1.2; range 26, 30) and depression was evaluated using the 15-item Geriatric Depression Scale^27^ (GDS, 0.7 +/− 1.1; range 0, 4). Participants with a self-reported history of diabetes, hypertension, coronary artery disease, pulmonary disease, neurological disease, depression or anxiety were also excluded from the study, as were participants planning travel to alternate time zones within two weeks of study participation. Participant demographics, MoCA and GDS scores are listed for each study, and for the combined dataset in **Supplemental Table 1**.

A Consolidated Standards of Reporting Trials (CONSORT) diagram for the Benchmarking Study and Replication Study is provided in **Figure 3** (bottom). Within the Benchmarking Study, the first three participants were removed from analysis because of a design change in the investigational device sensor positions. One participant was unable to complete the first MRI session and withdrew from the study. Of the remaining 30 participants (61.8 ± 2.7 years of age; 14 female, 16 male) that completed the Benchmarking Study, five overnight sleep studies and eight overnight wake studies failed data quality control due to excessive artifacts in the recordings, leaving 25 overnight sleep studies and 22 overnight wake studies in the Benchmarking Study with complete device, PSG and MRI data. Excessive movement artifacts during the morning recording periods resulted in the exclusion of data from five additional morning wake and one additional morning sleep opportunity periods, leaving 20 overnight sleep/morning wake and 21 overnight wake/morning sleep opportunity datasets with complete overnight and morning data. All participants enrolled in the Replication Study (55.6 ± 4.6 years of age; 7 female, 7 male) completed the protocol. All overnight sleep data were usable, but three overnight wake studies were removed, two because of excessive artifact and one because of non-compliance with the wake protocol. The higher number of quality control failures in the Benchmarking Study was largely due to the physical challenges caused by simultaneous data acquisition from both PSG and investigational device on the same night. Note that when the investigational device was used in the absence of PSG in the Replication Study, quality control failures were less frequent and restricted to motion artifacts in the awake condition.

### Assessment of sleep and wake changes in parenchymal resistance

Study participants underwent one night of normal sleep and one night of sleep deprivation separated by 2-4 weeks. Sleep/wake or wake/sleep visit order was determined by random assignment following informed consent. In the Benchmarking Study, sleep and wake status was confirmed by PSG with anticipated differences in rapid eye movement (REM), and non-REM (NREM: N1, N2, N3) sleep stages observed between sleep and wake conditions. Similar differences were observed in the Replication Study, in which investigational device-based EEG was used to evaluate sleep parameters. EEG was assessed with the investigational device in both the Benchmarking and Replication Studies; these measures were used when combining EEG parameters between both studies. Averaged hypnograms showing sleep stage distribution throughout the overnight sleep period for the Benchmarking and Replication Studies are shown in **Figure 4a-b** (top), respectively. The averaged hypnogram for the overnight and morning periods for the Benchmarking study participants with complete data during both periods is shown in **Figure 4c** (left).

**Figure 4.**
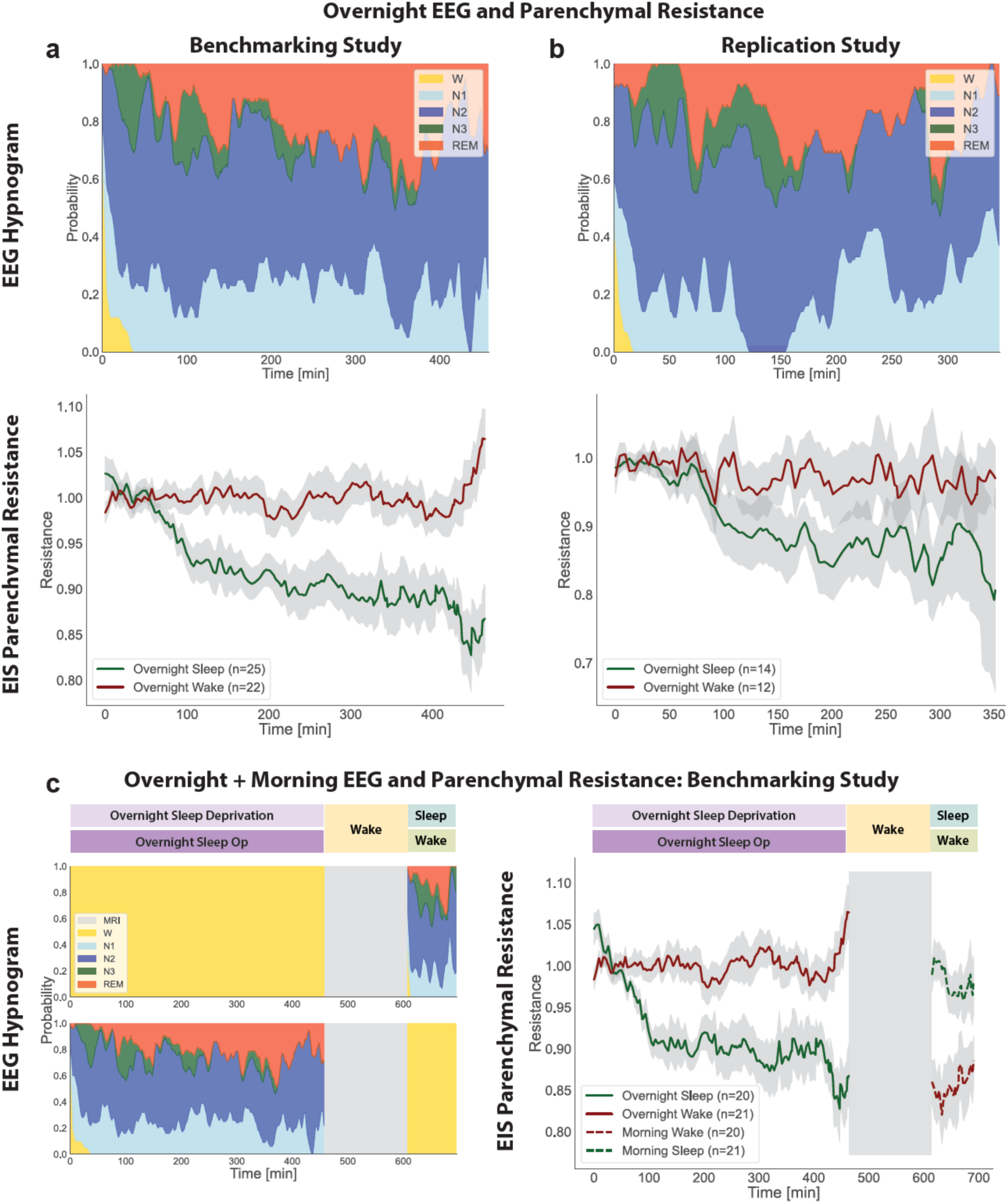
Brain parenchymal resistance is reduced periods of sleep. **a.** Averaged EEG hypnograms from the sleep condition are shown for the overnight period of the Benchmarking Study with WASO excluded in both the hypnograms and sleep R_P_ (top). Over that period, parenchymal resistance (R_P_, bottom) remained constant during the awake state (red) but declined gradually in the sleep state (green). **b.** Similar trends were observed in the Replication Study. **c.** During the morning period of the Benchmarking Study, participants underwent a 1.5 hr sleep opportunity or period of wakefulness. Averaged EEG hypnograms for the overnight and morning periods are shown at left. At right, during the morning awake period, R_P_ increased gradually (dashed red), while it declined gradually during the sleep state (dashed green). Parenchymal resistances R_P_ shown **(a)**, **(b)** and **(c)** are the average across the participants after normalizing each participant’s R_P_ measurements by their value at onset of the sleep or wake period. R_P_ in **(c)** for morning wake is plotted starting at the ending value of overnight R_P_ for illustration purposes. Standard error of the means are shown for each plot in light grey. The combined overnight and morning Benchmarking Study **(c)** includes participants who’s overnight and morning data passed quality controls.

During the overnight period R_P_ was continuously monitored. The results from the Benchmarking Study are provided in **Figure 4a** (bottom), which depicts the mean and standard errors of overnight R_P_. Each participant’s R_P_ was normalized to the R_P_ at onset of the observation period, and linear interpolation was used for missing R_P_ values within each participant. Compared to the wake condition, overall R_P_ during sleep was reduced by 8% (Wake 1.00 ± 0.06, Sleep 0.92 ± 0.06, T-test p < 0.001). When evaluated through the course of the overnight period, the R_P_ during the wake period remained largely constant. In contrast, during the sleep period the mean R_P_ decreased monotonically, reaching at the end of the sleep period a nadir ∼20% lower than at onset or under the awake condition. Similar results were observed the Replication Study (**Figure 4b**, bottom). Within the Benchmarking study, participants that were sleep deprived overnight received a 1.5 hr sleep opportunity in the morning. Over this period, R_P_ began at a higher value (resulting from overnight sleep deprivation) but declined over the 1.5 hr sleep opportunity (**Figure 4c**, right). Among participants that slept overnight but were kept awake in the morning, R_P_ during the morning period began at a low value (resulting from overnight sleep) but increased over the 1.5 hr period of wakefulness. These data demonstrate across two study populations that R_P_ remains constant or increases during periods of waking but declines steadily through periods of sleep.

### Association between sleep parameters and glymphatic function

Following the overnight sleep periods, participants in the Benchmarking Study underwent CE-MRI for the assessment of glymphatic function. Beginning at 7am, participants underwent a pre-contrast T1-weighted MRI scan, received intravenous GBCA (Gadavist, 0.1 mmol/kg), followed by T1-weighted MRI scans at 7-10 min (T_10_) and 240 min (T_240_) post-injection. We evaluated contrast enhancement in eight regions of interest (ROIs): frontal cortical gray and white matter, parietal cortical gray and white matter, temporal cortical gray and white matter, and occipital gray and white matter. Contrast enhancement at the T_10_ time point primarily reflects GBCA within the blood column, while enhancement at the T_240_ time point reflects GBCA leakage into the cerebrospinal fluid (CSF) and into the brain interstitial compartments^14,15^. Glymphatic function was defined by measuring brain parenchymal contrast enhancement, the % change in T1-weighted signal intensity between 10-240 min post-GBCA injection (100% * (T_240_-T_10_)/T_0_) within each ROI.

To evaluate different sleep-related contributions to glymphatic function, we developed a general random intercept mixed model with eight ROI groups fit to the T_10_-T_240_ change in brain parenchymal enhancement. Our **null hypothesis** was that parenchymal contrast, including contrast in the interstitial compartment, was not influenced by sleep variables. Glymphatic function supports the movement of solutes into and out of the brain interstitial compartment, yet brain parenchymal enhancement includes contrast signal from the blood compartment (the initial source of the GBCA) and the CSF compartment (the intermediate space linking the blood and brain interstitium) in addition to the interstitial compartment (**Figure 1f**). Thus, we defined the **null model** to include regressors for vascular and CSF contrast in the mixed model: the T_10_-T_240_ change in blood signal within the internal carotid artery, and the T_10_-T_240_ change in CSF signal within the cerebral lateral ventricles. We also included the confounding biological variables of age, gender, and APOE ε4 status in the null model *shown in* **Supplemental** (bottom). The analyses of EEG powerbands, heart rate and sleep staging were performed on the overnight sleep period recordings. Pulse plethysmography and EEG were acquired concurrently and heart rate, EEG powerbands, sleep stage durations (REM; N1, N2, N3 NREM), number of awakenings and wake after sleep onset (WASO) were computed and included independently in the null model (**Supplemental Table 2**), and likelihood ratio tests used to compare the individual predictor model with the null model. Relative EEG delta, theta, alpha and beta powerbands were each expressed as the overnight mean of the powerband normalized to the average powerband value of the first three sleep epochs. Higher EEG delta power (mean 19.12, 95% CI 5.4819, 32.7599; P=0.0072), lower EEG alpha (mean −7.28, 95% CI −12.2638, −2.2935; P=0.0052) and beta (mean −5.33, 95% CI −7.7859, −2.8728; P<0.001) power were significantly associated with greater contrast enhancement. We next evaluated EEG-derived sleep stages within individual mixed models. Neither number of awakenings during the sleep period, nor wake after sleep onset (WASO) were associated with parenchymal contrast enhancement. Lower N1 sleep time was significantly associated with greater parenchymal contrast enhancement (mean −0.1274, 95% CI −0.2130, −0.0420; P=0.0043), while N2, N3 and REM sleep time had no significant association with parenchymal enhancement within these individual models (**Supplemental Table 2**). Thus, within individual mixed models, increased overnight EEG delta band power, reduced alpha and beta band power, and reduced time in N1 sleep are each associated with greater glymphatic function, each rejecting the null hypothesis that parenchymal contrast was not influenced by sleep variables (**Supplemental Table 2**).

### Direct association between parenchymal resistance and glymphatic function

The device R_P_ values were computed repeatedly throughout the overnight sleep period from each EIS scan. During sleep recordings, the R_P_ was non-stationary, decreasing throughout the night as shown in **Figure 4a-c**. We observed that lower overnight R_P_ was significantly associated with higher glymphatic contrast enhancement (mean −38.08, 95% CI −61.34805, - 14.8115; P=0.0018, **Supplemental Table 2**).

### Combined effects of overnight parenchymal resistance, EEG spectral power, and heart rate on glymphatic function

We next utilized a comprehensive mixed model to define the contributions of R_P_, EEG spectral band power, and heart rate to parenchymal contrast enhancement. As in the individual mixed models above, this comprehensive model extended the null model that contains only contrast enhancement in the blood and cerebral ventricles as regressors, and age, APOE4 status, and gender as confounding biological variables. We evaluated the relative delta, theta and beta powerbands, omitting the alpha powerband to avoid multicollinearity in the model. We subjected the model to a backward elimination algorithm with a Wald’s p-value threshold of 0.05. **Table 1** provides the full outputs of the analyses for each of the four intervention periods of overnight sleep opportunity, morning sleep opportunity, overnight sleep deprivation and morning awake of the study (**Figure 3**, top).

**Table 1.**
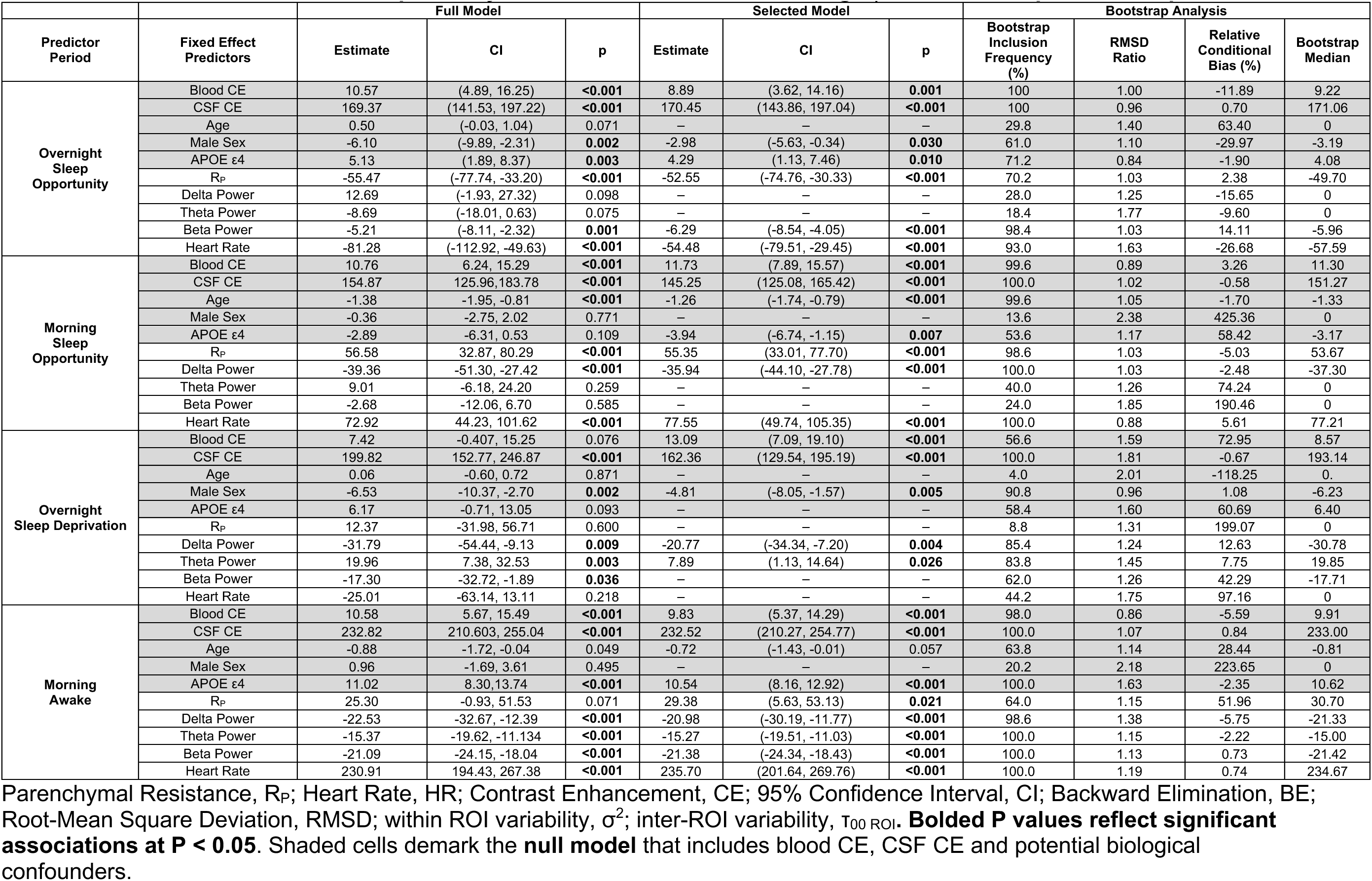
Multivariate models of brain parenchymal contrast enhancement using R_p_, HR and EEG power band predictors.

In reference to the models selected by backward elimination shown in **Table 1**, as expected, both blood and ventricular CSF contrast enhancement were strongly associated with parenchymal enhancement following the overnight sleep opportunity (blood mean 8.89; 95% CI 3.62, 14.16; P = 0.001; CSF mean 170.45; 95% CI 143.86, 197.04; P<0.001) and overnight sleep deprivation (blood mean 13.09; 95% CI 7.09, 19.10; P < 0.001; CSF mean 162.36; 95% CI 129.54, 195.19; P<0.001). For the overnight sleep opportunity, lower heart rate (mean - 54.48, 95% CI −79.51, −29.45; P<0.001) and lower R_P_ (mean −52.55, 95% CI −74.76, −33.33; P<0.001) predicted greater contrast enhancement. Reduced EEG beta power predicted greater parenchymal enhancement (mean −6.29, 95% CI −8.54, −4.05; P < 0.001), while neither delta nor theta power exhibited significant associations within the multivariate model. In the overnight sleep deprivation condition, reduced EEG delta power (mean −20.77, 95% CI −34.34, −7.20; P = 0.004) and increased theta power (mean 7.89, 95% CI 1.13, 14.64; P = 0.026) predicted greater parenchymal enhancement, while neither beta power, heart rate nor R_P_ showed significant associations within the multivariate model.

### Combined effects of morning parenchymal resistance, EEG spectral power, and heart rate on glymphatic function

As shown in the study schematic (**Figure 3**, top), between 7:10 am and 9:30am (140 min post-injection, T_140_) participants underwent MRI scanning followed by instrumentation with the investigational device and PSG for the morning sleep opportunity or awake intervention. Because the mean terminal half-life of intravenous GBCA in adults with normal renal function is 109 min^28^, the post-injection blood contrast during the morning recordings was 60% cleared by T_140_ and 80% cleared by T_240_. Glymphatic function in the period between T_140_ and T_240_, would therefore dilute existing parenchymal contrast with CSF having 60% to 80% lower contrast concentration, a process analogous to ‘clearance’ of interstitial contrast. Thus, we would expect that an increase in glymphatic function during the period between T_140_ and T_240_ would be reflected as reduced parenchymal contrast enhancement at T_240_ (**Supplemental Figure 1**).

In reference to the models selected by backward elimination in **Table 1**, once again both blood and ventricular CSF contrast enhancement were strongly associated with parenchymal enhancement following the morning sleep opportunity (blood mean 11.73; 95% CI 7.89, 15.57; P < 0.001; CSF mean 145.25; 95% CI 125.08, 165.42; P<0.001) and awake intervention (blood mean 9.83; 95% CI 5.37, 14.29; P < 0.001; CSF mean 232.52; 95% CI 210.27, 254.77; P<0.001). During the morning sleep opportunity, R_P_ begins to decrease (**Figure 4c**). Higher EEG delta power (mean −35.94, 95% CI −44.10, −27.78; P < 0.001) and lower R_P_ (mean 55.35, 95% CI 33.01, 77.70; P < 0.001) predicted reduced parenchymal enhancement (greater contrast clearance).

During the morning awake period, R_P_ increases from its overnight sleep ending level (**Figure 4c**). During this intervention, higher EEG delta power (mean −20.98, 95% CI −30.19, −11.77; P < 0.001), theta power (mean −15.27, 95% CI −19.51, −11.03; P < 0.001) and beta power (mean - 21.38, 95% CI −24.34, −18.43; P < 0.001) predicted greater contrast clearance as did lower heart rate (mean 235.70, 95% CI 201.64, 269.76; P < 0.001) and lower R_P_ (mean 29.38, 95% CI 5.63, 53.13; P < 0.021). These findings are similar to those observed for the morning sleep opportunity and consistent with the explanation that an increase in glymphatic influx of CSF during this period dilutes the ISF contrast concentration and reduces the T_240_ MRI signal intensity.

Parenchymal Resistance, R_P_; Heart Rate, HR; Contrast Enhancement, CE; 95% Confidence Interval, CI; Backward Elimination, BE; Root-Mean Square Deviation, RMSD; within ROI variability, σ^2^; inter-ROI variability, τ_00 ROI_**. Bolded P values reflect significant associations at P < 0.05**. Shaded cells demark the **null model** that includes blood CE, CSF CE and potential biological confounders.

We used bootstrap analysis of the backward elimination algorithm for stability investigations on the impact of variable selection on bias and variance of the regression coefficient and bootstrap inclusion frequencies. We estimated the multivariate model using the backward elimination algorithm on 500 bootstrap data replicates with replacement for each intervention period (**Table 1**). The bootstrap inclusion frequency represents the percent of the 500 backward elimination models that retained each variable, while the bootstrap median is the median estimate for that variable. The relative conditional bias quantified the omitted variable selection bias of each variable from inclusion/non-inclusion of other variables from the full model. In the overnight sleep opportunity, in addition to vascular and CSF contrast enhancement (each with an inclusion frequency of 100%), R_P_ (70.2%), EEG beta power (98.4%), and heart rate (93.0%) had high bootstrap inclusion frequencies and bootstrap median estimates comparable to the selected model estimates. Device-measured R_P_ and ventricular contrast enhancement had the lowest relative conditional bias, and therefore their predictions of parenchymal contrast uptake were least affected by inclusion of the other variables. Findings in the morning sleep opportunity also showed high inclusion frequencies of the variables retained in the backward elimination algorithm on the full data set and bootstrap median estimates comparable to the selected model estimates, with vascular contrast enhancement (99.6%), CSF contrast enhancement (100%), R_P_ (98.6%), EEG delta power (100%), and heart rate (100%). These regressors also had low relative conditional bias. These analyses suggest high model stability to perturbations of the data set^29^.

Lastly, we computed the percent of the residual variance of the null model that was explained by including EEG power band, heart rate and R_P_ regressors for each intervention and computed the LRT between the null model and the intervention models. For the overnight sleep and sleep deprivation interventions, 24.5% and 12.7% of the residual null model variance was explained by including these regressors, respectively. The null hypothesis that parenchymal contrast enhancement is not influenced by these variables was rejected for the overnight sleep model (likelihood ratio 52.6, p-value < 0.001) and the sleep deprivation model (likelihood ratio 14.1, p-value = 0.015). Including the regressors in morning sleep and awake models explained 38.3% and 74.8% of the residual null model variance, respectively. The null hypothesis that the parenchymal contrast enhancement is not influenced by these variables during the morning sleep (likelihood ratio 79.5, p-value < 0.001) and awake condition (likelihood ratio 149.6, p-value < 0.001) were also both rejected.

### Combined effects of parenchymal resistance and sleep stages on glymphatic function

We developed a second comprehensive mixed model to define the relationship among R_P_, sleep stages, heart rate and parenchymal contrast enhancement. As above, this comprehensive model extended the null model that contained contrast enhancement in the blood and cerebral ventricles as regressors, and age, APOE4 status, and gender as confounding biological variables. We evaluated WASO time, and time in N1, N2, N3 and REM sleep stages. **Supplemental Table 3** provides the model output for the overnight sleep opportunity (top) and the morning sleep opportunity (bottom). In the overnight sleep opportunity model selected by backward elimination, as before lower R_P_ was significantly associated with greater parenchymal contrast enhancement (mean −67.17; 95% CI −87.78, −46.56; P<0.001). In this model, more time in REM sleep (mean 0.16; 95% CI 0.09, 0.23; P<0.001), and less time in N1 (mean −0.32; 95% CI −0.43, −0.21; P<0.001), N2 (mean −0.10; 95% CI −0.15, −0.06; P<0.001), N3 (mean −0.07; 95% CI −0.12, 0.02; P=0.007) sleep and WASO (mean −0.07; 95% CI −0.12, −0.02; P=0.006) were each associated with greater contrast enhancement. Bootstrap analysis showed that blood and CSF contrast enhancement (each with an inclusion frequency of 100%), as well as R_P_ (100%), REM sleep time (99.0%), N1 sleep time (98.8%) and N2 sleep time (98.0%) were the most stable elements of the model relating sleep stages to parenchymal glymphatic function, while the RMSD ratio for N2 sleep time, N3 sleep time and WASO showed substantial variance deflation following backward elimination (**Supplemental Table 3**) that is associated with weak or noise predictors.

In the morning sleep opportunity model selected by backward elimination, lower R_P_ was significantly associated with greater parenchymal contrast clearance (mean 34.42; 95% CI 5.25, 63.59; P = 0.025), similar to the EEG band power model for this intervention. In this model, less time in REM sleep (mean 0.41; 95% CI 0.20, 0.62; P<0.001), and more time in N1 (mean −0.26; 95% CI −0.50, −0.02; P=0.04) and N3 (mean −0.17; 95% CI −0.33, −0.021; P=0.031) were associated with less contrast enhancement, or greater contrast clearance. N3 sleep did not survive bootstrap analysis while R_P_, REM and N1 sleep had inclusion frequencies of 65.4%, 88.2% and 61.6%, respectively (**Supplemental Table 3).**

### Effects of sleep stages on brain parenchymal resistance

To understand if sleep stages had a differential effect on the overnight change in R_P_, we computed the mean of the first-order differences ΔR_P_ for each sleep stage for the Benchmarking and Replication studies separately and combined (**Figure 5**). The mean ΔR_P_ was negative and largest for N2, N3 and REM sleep stages reaching significance when the values for both studies where pooled together. This suggests that R_P_ in **Figure 4a-c** decreases during N2, N3 and REM sleep.

**Figure 5.**
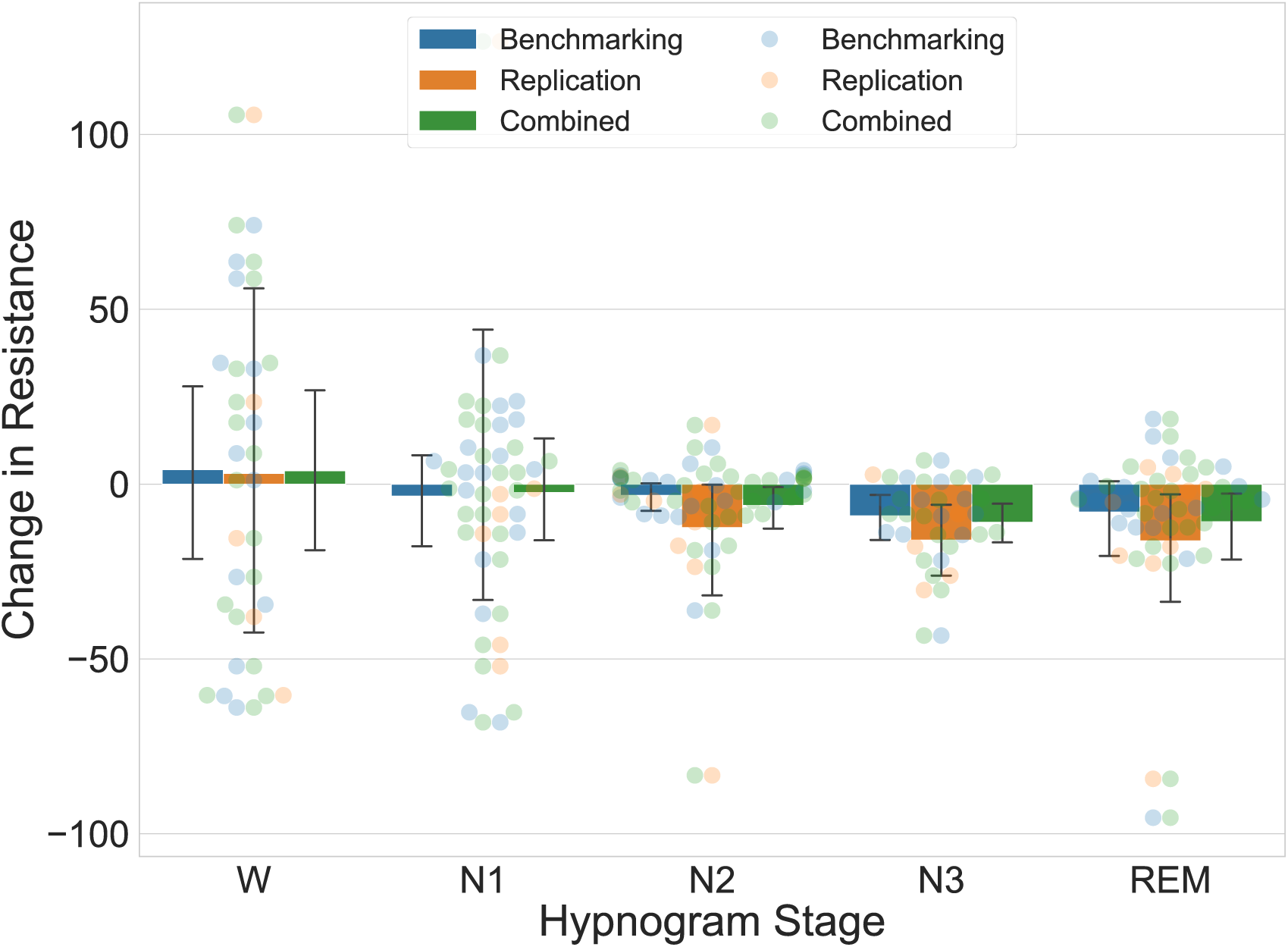
Effect of sleep stages on R_P_. The mean first-order difference ΔR_P_ is shown by sleep stage and study for the overnight sleep period with 95% confidence intervals. During N2, N3 and REM sleep ΔR_P_ is negative for each study and when combined reaches significance. Benchmarking N2 (mean −3.21, 95% CI −9.07, −0.33; P = 0.113), N3 (mean −9.07, 95% CI - 17.04, −3.75; P = 0.005), REM (mean −8.01, 95% CI −25.64, −1.18; P = 0.133). Replication N2 (mean −12.46, 95% CI −34.94, −1.49; P = 0.115), N3 (mean −16.04, 95% CI −25.32, −5.32; P = 0.002), REM (mean −16.24, 95% CI −42.19, −4.65; P = 0.065). Combined N2 (mean −6.10, 95% CI −14.78, −1.71; P = 0.04), N3 (mean −10.91, 95% CI −17.35, −5.93; P < 0.001), REM (mean - 10.75, 95% CI −23.86, −3.91; P = 0.023). Units of ΔR_P_ are mΩ.

### Effects of large changes in EEG spectral power on brain parenchymal resistance

To define the relationships between sleep EEG powerbands, heart rate and R_P_, we modeled the relationship between the first-order difference ΔR_P_ and the difference in spectral band power (Δdelta, Δtheta, Δalpha, Δbeta) and changes in heart rate ΔHR, during NREM, REM and Wake (**Table 2**). We utilized threshold regression to model the relationships between ΔR_P_ and large changes in (Δ) band power or heart rate. A custom estimation procedure described in **Supplemental Information** was used to jointly estimate the threshold regression for the full positive and negative ranges of β band power and ΔHR. The full output of the threshold regression models is provided for the combined study data in **Table 2**. In both REM and NREM sleep, a large increase in Δdelta was significantly associated with a decrease in ΔR_P_ (**Figure 6a**). In contrast during REM and NREM sleep, a large increase in Δbeta was significantly associated with an increase in ΔR_P_. The threshold models for the significant estimated effects of β band power on ΔR_P_ are plotted in **Figure 6b**. The full output of the threshold regression model is provided for each site individually in **Supplemental Table 4**. Because the variables were standardized before model estimation, the regression estimates in **Table 2** and **Supplemental Table 4** can be interpreted as standardized effect sizes. Note that because of the slope ratio *r*, changes that exceed the positive change point *t_p_* have effect sizes given by the regression estimate multiplied by the slope ratio. The positive and negative thresholds are in units of standard deviation.

**Figure 6.**
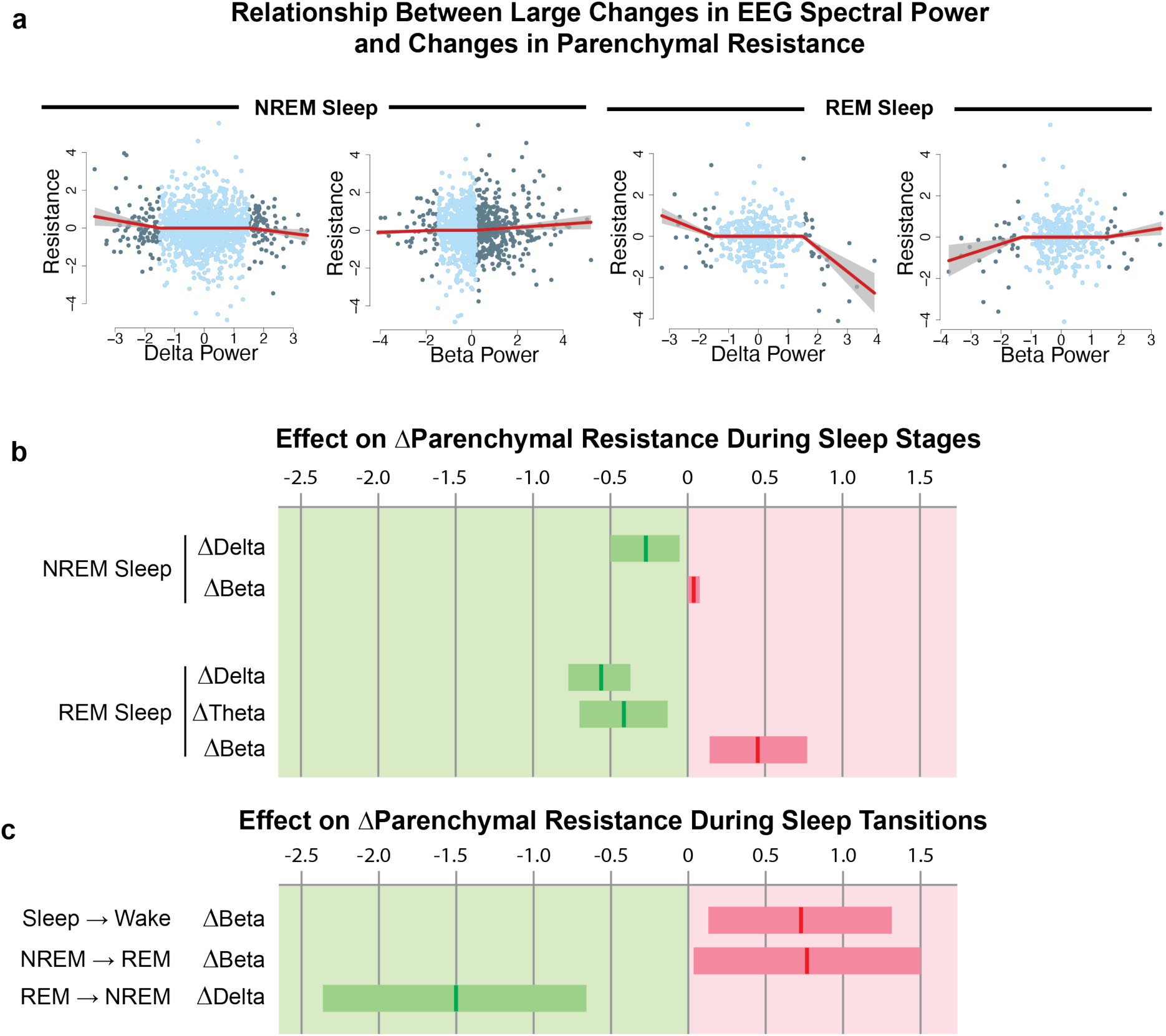
Brain parenchymal resistance is increased by large changes in EEG beta power and reduced by large changes in EEG delta power. **a**, Threshold linear mixed regression model of R_P_ against EEG delta and beta power for REM and NREM sleep shown with standard error after differencing both R_P_ and powerband values to make data stationary, standardizing the ΔR_P_ and Δ powerband values to zero mean and unit standard deviation, and adjusting for site, age, gender, APOe4 and site-age interaction confounders. Units of ΔR_P_ and Δ powerband are in standard deviations. Data in light blue are inside the change points and data in dark blue are outside the change points. When delta and beta power changes between successive measurements exceed 1.0 to 1.5 stand deviations, we observe significant changes in ΔR_P_**. b**, 95% CI of regression coefficients show that large changes in REM delta, theta and beta, and NREM delta and beta are significant predictors of ΔR_P_. For a 1 standard deviation increase in Δ powerband, an increase (decrease) in ΔR_P_ is illustrated in red (green) with units along the top in standard deviation of ΔR_P._ **c**, Large changes in beta and delta powerbands at sleep-wake and NREM-REM transitions show significant effect on ΔR_P_ consistent with the step change in beta and delta power at these transitions. For a 1 standard deviation increase in β powerband across a sleep-wake or NREM-REM transition, an increase (decrease) in ΔR_P_ is illustrated in red (green) with units along the top in standard deviation of ΔR_P._ these data show that glymphatic function is enhanced by declining R_P_, and that R_P_ in turn declines with reduced EEG beta power and heart rate, and with increasing EEG delta power.

**Table 2.** The change in parenchymal resistance resulting from large changes in EEG band power and heart rate occurring during REM sleep, NREM sleep, wake, and at NREM-REM and sleep-wake transitions.

| Study Condition | Period | Predictors | Fixed Effects |  |  |  | Threshold Model Parameters |  |  |
| --- | --- | --- | --- | --- | --- | --- | --- | --- | --- |
|  |  |  | Estimate | CI | p | DF | Positive Threshold | Negative Threshold | Slope Ratio |
| Overnight Sleep Op | NREM | Δdelta | -0.28 | (-0.50, -0.058) | **0.014** | 1380 | 1.5 | -1.5 | 0.7 |
| | | Δtheta | -0.08 | (-0.23, 0.063) | 0.265 | 1380 | 1.5 | -0.98 | 0.5 |
| | | Δalpha | -0.14 | (-0.30, 0.022) | 0.091 | 1380 | 1.5 | -1.25 | 0.65 |
| | | Δbeta | 0.04 | (0.0022, 0.080) | **0.039** | 1380 | 0.18 | -1.5 | 2 |
| | | ΔHR | 0.10 | (-0.0033, 0.19) | 0.059 | 1115 | 0.43 | -0.43 | 0.5 |
| | REM | Δdelta | -0.57 | (-0.77, -0.37) | **<0.001** | 319 | 1.5 | -1.5 | 2 |
| | | Δtheta | -0.41 | (-0.70, -0.13) | **0.005** | 319 | 0 | -1.5 | 0.5 |
| | | Δalpha | -0.09 | (-0.21, 0.041) | 0.189 | 319 | 1.45 | 0 | 2 |
| | | Δbeta | 0.45 | (0.14, 0.77) | **0.006** | 319 | 1.5 | -1.3 | 0.5 |
| | | ΔHR | 0.31 | (-0.057, 0.68) | 0.101 | 271 | 1.5 | -1.15 | 0.5 |
| | NREM to REM | Δdelta | -0.29 | (-0.82, 0.24) | 0.293 | 67 | 1.5 | -1.05 | 2 |
| | | Δbeta | 0.77 | (0.04, 1.49) | **0.046** | 67 | 0.6 | -1.5 | 0.5 |
| | REM to NREM | Δdelta | -1.50 | (-2.35, -0.66) | **0.001** | 58 | 1 | -1.5 | 0.5 |
| | | Δbeta | -0.05 | (-0.24, 0.14) | 0.615 | 58 | 0.1 | -1.5 | 1.85 |
| | Sleep to Wake | Δdelta | 0.03 | (-0.34, 0.4) | 0.885 | 66 | 1.5 | -0.3 | 1.95 |
| | | Δbeta | 0.73 | (0.13, 1.32) | **0.021** | 66 | 0.35 | -1.23 | 0.5 |
| | Wake to Sleep | Δdelta | -0.20 | (-0.64, 0.25) | 0.392 | 113 | 0.05 | -1.35 | 0.6 |
| | | Δbeta | 0.37 | (-0.01, 0.76) | 0.065 | 113 | 1.5 | -1.05 | 2 |
| Overnight Sleep Deprivation | WAKE | Δdelta | -0.06 | (-0.15, 0.034) | 0.216 | 1137 | 1.5 | 0 | 0.5 |
| | | Δtheta | -0.07 | (-0.17, 0.035) | 0.198 | 1137 | 1.4 | -1.45 | 2 |
| | | Δalpha | 0.06 | (-0.030, 0.14) | 0.206 | 1137 | 1.15 | 0 | 0.73 |
| | | Δbeta | 0.12 | (0.0075, 0.23) | **0.037** | 1137 | 1.5 | -1.43 | 1.93 |
| | | ΔHR | -0.10 | (-0.20, -0.011) | **0.029** | 1002 | 0.25 | -0.25 | 0.5 |

Rapid eye movement, REM; non-REM, NREM; Confidence Interval, CI; Degrees of Freedom, DF; EEG band power predictor units in percent of total band power; HR predictor units beats-per-minute. **Bolded P values reflect significant associations at $P < 0.05$ .** Predictor and parenchymal resistance values in regression models were scaled to zero mean and unit standard deviation with coefficient estimates representing effect size in standard deviations. Positive and negative thresholds are in units of predictor standard deviation. Models included confounding biological variables age, gender, and APOE $\epsilon 4$ status.

The asymmetric effect of EEG band power changes on R_P_ shown in **Figure 5** accounts for the progressive decline in R_P_ that occurs through the course of the overnight sleep period, shown in **Figure 4a-c**. In the overnight wake state, Δbeta had a significant but small effect on ΔR_P_. ΔHR in NREM and wake had a small effect on ΔR_P_ and a larger but non-significant effect in REM. The threshold model findings in **Table 2** and plots in **Figure 6a** demonstrate that large changes in spectral band power are needed to alter R_P_. Because such large changes in EEG powerbands and HR occur at sleep stage transitions, we explored the associations between Δdelta and Δbeta and ΔR_P_ at NREM-REM and REM-NREM transitions, and at Sleep-Wake and Wake-Sleep transitions. **Table 2** provides the full model output on sleep stage transitions for data pooled from both study sites, while the significant estimated effects of Δdelta and Δbeta on ΔR_P_ are plotted in **Figure 6c**. The full output of the threshold regression model on sleep stage transitions is provided for each site individually in **Supplemental Table 5**. During the NREM-REM, Wake-Sleep, and Sleep-Wake transitions, changes in EEG beta power and not EEG delta power underlie the observed changes in ΔR_P_. In contrast, during the REM-NREM transition, changes in EEG delta power and not beta power were associated with changes in ΔR_P_. In total,

## DISCUSSION

We have described a non-invasive multimodal skin-interfaced wireless device for continuous measurement of brain parenchymal resistance R_P_ using repeated EIS time-multiplexed with EEG and cardiovascular measurements. Through two separate clinical validation studies, we observe that brain R_P_ measured by the device faithfully captures sleep-active changes in glymphatic flow resistance based on three complementary criteria established from studies in the murine and human brain. Brain R_P_ declines overnight, paralleling sleep-associated changes in extracellular volume that regulate glymphatic function^2^. Brain R_P_ is reduced with increasing sleep EEG delta power, and is increased with increasing sleep EEG beta power and heart rate^11^. Lastly, brain R_P_ is a robust predictor of glymphatic function, measured by CE-MRI^12–15,30^. These findings demonstrate that this investigational device provides the first remote, non-invasive, and time-resolved measure of glymphatic flow resistance suitable for defining this sleep-active biology in naturalistic settings.

Several technological advances have enabled this work. We have developed a technique for low-noise EIS measurements using multiple excitation and sense configurations to determine the brain parenchymal impedances accurately. A signal processing approach enabled the segregation of signal and noise, using the Kramers-Kronig relations^31,32^ between the real and imaginary parts of the impedance to establish if the measured response is related to the excitation signal or has been distorted by noise or other artifact. Unlike applications of EIS to neurological conditions such as seizures, tumors or cerebral edema^33–36^, the rapid evolution of glymphatic function during sleep and its nightly variability make it suitable for monitoring with longitudinal repeated assessments. Data collected under a variety of scenarios in benchtop testing, agar phantom testing with biological interfaces and in-human testing indicate that this system yields consistent, repeatable results and shows longitudinal stability of the measured impedances.

A key first step in validating any potential measure of glymphatic function is assessing its performance against established, principally contrast-based, measures of glymphatic exchange. Initial studies characterizing glymphatic function in rodents utilized dynamic in vivo fluorescence-based imaging^1,2,37^. More recent studies in rodents, and recently in human participants, have utilized CE-MRI following either intrathecal or intravenous GBCA injection^12,14,15,30,38^. We evaluated whether device-based R_P_ measures reflect glymphatic function by measuring overnight and morning R_P_, EEG, and the movement of GBCA into brain tissue over a four-hour timeframe. In the analysis, both overnight and morning R_P_ emerged as robust explanatory variables of net contrast movement, in a regression model that included CSF and blood contrast levels, EEG neurophysiological parameters, age, gender and APOE e4 confounders. It is important to note that device-based R_P_ measures are acquired with 2-minute temporal resolution whereas CE-MRI captures overall glymphatic function over a 4-hour window. This fine temporal resolution enabled translational validation of the physiological drivers of glymphatic function in the human brain for the first time. It further revealed that while key events during sleep were necessary to lower parenchymal resistance R_P_, low R_P_ was sufficient to enhance glymphatic function independent of sleep in the morning period. In addition, it showed that sleep-related changes in RP do not shift with sleep-wake in a stepwise fashion, but rather accumulate through the course of time spent sleeping or awake. These observations are not inconsistent with prior studies in rodents that used real-time iontophoresis to measure sleep-associated increases in brain extracellular volume fraction^2^, as these observational studies did not directly test the causal effect of these changes in extracellular volume on glymphatic function, and did not monitor changes in extracellular volume fraction in a time-resolved manner over extended period. Future studies of the determinants of glymphatic function in human sleep will likely further refine existing conceptual models of sleep-wake regulation of glymphatic function.

The physiological mechanisms regulating glymphatic function have been defined almost exclusively through experimental rodent studies^1,2,10,11,30,37,39,40^, due in large part to invasiveness and poor temporal resolution of contrast-based measures of glymphatic function^12–15,30^. It is noteworthy that these rodent studies are not all in agreement. While multiple studies have reported more rapid glymphatic exchange under conditions of sleep, or under certain anesthetic regimes^2,11,41–45^, two studies^46,47^ have reported apparently slower clearance during sleep or under anesthesia. In the present human clinical studies, we demonstrate within-participant longitudinal declines in brain R_P_ during overnight sleep, but not in awake participants. During morning recovery sleep, we demonstrate within-participant longitudinal declines in brain R_P_ and during morning wake we observe increases in brain R_P_. This pattern is consistent with sleep-wake changes in extracellular volume fraction measured in mice and reported by Xie et al.^2^. Using overnight PSG followed by CE-MRI in human participants, we confirmed that glymphatic function in the human brain is associated with lower EEG beta power and heart rate, and in a simplified model with increasing EEG delta power. These findings sustain the physiological findings in mice from Hablitz et al.^11^. More broadly, these human CE-MRI-based imaging studies demonstrate that glymphatic solute exchange is more rapid under conditions of natural sleep, at least in human participants, and that these sleep-wake differences are regulated by sleep-related changes in R_P_, heart rate, and EEG spectral band power.

Linking changes in R_P_ with sleep EEG features, we further demonstrate that the investigational device-based measure of R_P_ exhibits the same dependence upon EEG delta power, EEG beta power, and heart rate during sleep. Conducting parallel time-resolved sleep EEG and R_P_ measurements, we observed that rapid changes in R_P_ occurred at sleep stage transitions between REM and NREM, and between sleep and wake. Transitions between REM and NREM exhibit large changes in EEG delta and beta powerbands, and in HR, consistent with our findings. Large changes in an EEG power within a frequency band can occur because of sudden bursts of oscillatory activity within the band’s frequency range. In rodents, recent experiments support a role for such synchronized oscillatory activity as a mechanism driving glymphatic flow by creating propagating ionic waves within the brain interstitium^41^. To our knowledge, this is the first time that these physiological drivers of glymphatic function have been confirmed in human participants, within an experimental setting permitting validation against current benchmark CE-MRI-based measures of glymphatic function well established preclinical features.

There are clear limitations both of the investigational device, and of the present study design. Existing MRI-based measures of glymphatic function remain poorly time-resolved but provide excellent cranium-wide anatomical resolution. In contrast, the present investigational device captures a measure of global brain glymphatic flow resistance with high temporal resolution. While finer anatomical resolution may have value in future iterations of the investigational device, it is important to note that the present design was sufficient to capture sleep-wake changes in glymphatic function, and to define novel neurophysiological drivers of these processes in the human brain. Study methodological limitations include protocol differences between the Benchmarking Study and Replication Study. The Benchmarking Study included CE-MRI as part of the neuroimaging package and gold-standard PSG concurrent with the device usage whereas the Replication Study did not include CE-MRI and used the device’s single-derivative EEG. Furthermore, the Benchmarking Study enrolled participants 55-65 years old living in Florida whereas the Replication Study enrolled participants 50-65 years old living on the West Coast. Despite these differences, after adjusting for age, study site and age-site interaction effects, we confirmed that the device’s single-derivative EEG showed good agreement with the PSG of the Benchmarking Study. Additionally, the pooled analysis and study-specific analyses of R_P_, EEG and HR were all in good agreement between studies. The Benchmarking Study was powered to detect a low to moderate correlation between R_P_ and CE-MRI measures of glymphatic function, which was confirmed. Quality control of the investigational device filtered out measurements that did not satisfy the Kramers-Kronig relations^31,32^. Failed measurements were more common in the Benchmarking Study, particularly during the awake conditions. This is attributable to excessive motion, likely due to the technical challenges associated with conducting concurrent PSG and device-based sleep EEG measures in participants being kept awake by study staff. The low rate of quality-control failures in the Replication Study that did not involve PSG instrumentation supports this conclusion.

The characterization of the glymphatic system in rodents beginning in 2012 has spurred a great deal of interest into its potential mechanistic role linking sleep to cognitive performance, and sleep disruption to a wide range of neurological and psychiatric conditions including Alzheimer’s disease, Parkinson’s disease, chronic traumatic encephalopathy, stroke, traumatic brain injury, headache and others^42–45^. Preclinical studies have demonstrated that glymphatic function is impaired in the setting of aging^48^, sleep disruption^2^, cerebrovascular injury^49,50^, and traumatic brain injury^3^, all risk factors for neurodegenerative conditions including Alzheimer’s disease. Experimental impairment of glymphatic function is sufficient to promote the development of the amyloid beta^51–53^ and tau pathology^3–5^ characteristic of Alzheimer’s disease. In clinical populations, genetic and histological associations support a link between glymphatic dysfunction and the development of clinical disease^52,54–56^. Yet the technical challenges of real-time dynamic measurements of glymphatic function in human clinical populations have proven a challenging barrier to defining the causal role of glymphatic function and dysfunction in the development of neurological and psychiatric conditions in these populations. Whether glymphatic dysfunction contributes to development of conditions such as Alzheimer’s disease remains to be directly tested. The present investigational device may permit the continuous and time-resolved assessment of glymphatic function in naturalistic settings necessary to define whether glymphatic impairment contributes to risk and progression of Alzheimer’s disease and its underlying pathological processes. The ability to assess glymphatic function over short timescales may enable target-engagement studies to identify and test pharmacological, device-based and lifestyle/behavioral interventions for modulating glymphatic function in humans. Assessment of these processes may also permit the identification of clinical populations with impaired glymphatic function, at risk for the development of Alzheimer’s disease, and who would thus be ideal candidates for therapeutic approaches targeting glymphatic clearance. Similar avenues now remain open outside the realm of neurodegenerative conditions as glymphatic dysfunction is implicated in an ever-increasing list of sleep-related neurological and psychiatric conditions.

## METHODS

### Investigational Device

#### System Architecture

The Applied Cognition device is a wearable multi-sensor acquisition system that uses an STM32WB5MMG microcontroller (ST Microelectronics, NV). It consists of a primary module housing the main electronics board and a 430 mAh Li-ion battery, two earpieces each housing one electrode, one accelerometer and one photoplethysmogram sensor, and two separate mastoid electrodes. A device schematic and visualization of signal outputs is provided in **Figure 2a-f**. Sensor data is stored in on-board FLASH memory and is downloaded via a USB port for offline analysis. The relevant sub-systems are further described below.

#### Electrical Impedance Spectroscopy

Electrical impedance spectroscopy (EIS) was used to measure electrical impedance using the AFE4500 analog front-end (AFE) from Texas Instruments. Four electrodes – two in-ear and two mastoid – were used to deliver the excitation current across the head and sense the resulting voltage. The two mastoid current injection sources were initially co-located inside the ear next to the Ag/AgCl sensors but because of salt-bridging were externalized to the mastoids. In a departure from traditional four-electrode measurements, where excitation and sense electrodes are fixed, the AFE4500 was programmed to implement a technique developed by Texas Instruments where multiple excitation and sense configurations are used to determine the contact impedance and body impedance accurately.

Measurements were successively performed at eighteen frequencies ranging from 1 kHz to 256 kHz following a logarithmic series, all with excitation current below 50 μA root-mean-square (rms). Frequencies were in alternating order from high-low to minimize any impact on impedance drift during each scan. Sensed voltages were quadrature-demodulated, filtered and digitized internally by the AFE4500 to provide in-phase and out-of-phase components. Measurement time for each frequency was adjusted to give consistent signal-to-noise ratio across the frequency range, resulting in a scan time of 106 seconds.

#### Electroencephalogram

Electroencephalogram (EEG) was measured with an ADS1299-4 AFE from Texas Instruments, between the two in-ear electrodes (differential measurement). A third electrode (left mastoid) was used to drive the common mode. While these electrodes were shared with the EIS, the two measurements were time-multiplexed and a pair of analog multiplexers (TMUX1136, Texas Instruments, Inc.) effectively decoupled EEG and EIS sub-circuits and respective measurements. For EEG, both in-ear electrodes were also buffered (OPA376, Texas Instruments, Inc.) in the earpiece. These buffers were bypassed (analog multiplexer TMUX1136, Texas Instruments, Inc.) for EIS or electrode impedance measurement.

The ADS1299-4 was programmed to have a sampling rate of 250 samples-per-second (24-bit samples) and an input range of ±180 mV. The common mode drive was actively derived from the in-ear electrodes. Both mastoid electrodes were also connected to an input channel so that the low-frequency (31.25 Hz) impedance of each electrode could be measured using the internal current source of the ADS1299-4. Such impedance measurements were performed before starting a recording session, and then automatically between EEG periods and EIS. EEG measurement periods were 170 seconds long, and the sequence EEG-EIS-Electrode impedance repeated until the end of the recording session. A sample EEG spectrogram and hypnogram, derived from the device EEG is shown in **Figure 2b**.

#### Impedance Plethysmogram

In addition to impedance spectroscopy, real-time bioimpedance was acquired simultaneously with the EEG, using the MAX30001 AFE (Analog Devices, Inc.). The four-electrode configuration shared electrodes with the EEG, with the two mastoid electrodes used for current injection and the two in-ear electrodes for voltage sensing. A 96 μA current at 82 kHz was used for excitation. Such high frequency excitation is effectively filtered by the ADS1299-4 and does not corrupt the EEG signal. The MAX30001 built-in synchronous demodulator filtered and ADC recovered the in-phase component of the sensed voltage and digitized it at 64 samples-per-second (20-bit resolution). The resulting impedance measurements had a noise floor of 3 mΩ rms (100 Ω load), enabling the detection of blood pulsations. A sample trace from a study participant, derived from the device IPG is shown in **Figure 2d**. Note that the impedance shifts measured in either the respiratory or cardiac frequency bands range between 50 to 150 mΩ. In contrast, impedance shifts detected by EIS were at least an order of magnitude greater.

#### Photoplethysmogram and Acceleration

In-ear reflective photoplethysmogram (PPG) was measured with two miniature, fully-integrated optical sensors MAXM86161 (Analog Devices, Inc.) located in each earpiece, facing the anterior wall of the ear canal. One side was programmed to use green light (530 nm), while the other used alternatively red (660 nm) and infrared (880 nm) lights. To achieve a tight synchronization with EEG samples, the sensors ran at 1024 Hz for each color, and the most recent samples available at the time of the EEG read were averaged, resulting in a 250 Hz output rate and a phase variation <1 ms. A sample trace of the left (red) and right PPG (green) from a study participant is provided in **Figure 2f**.

In-ear acceleration (X,Y,Z) was measured using two miniature MEMS Inertial Measurement Unit (IMU) LSM6DSOX (ST Microelectronics, NV) located in each earpiece. Similar to the PPG, the sensors were run at 833 Hz (each axis) and averaged to ensure tight time synchronization between sensors. A noise floor of 1 mg rms enabled the resolution of cardiac/blood pulsations, in addition to posture and activity. From these accelerometer data, a ballistocardiogram is derived, as shown for a study participant in **Figure 2e**.

#### Electrodes

In-ear electrodes were laser-cut from silver sheets 100 microns thick and 99.9% purity. The electrodes were 4.5 mm diameter plates with a 1 mm-wide stem. The electrodes were abraded and sonicated in distilled water, dip-coated with Ag/AgCl ink provided by Creative Materials Inc (SKU EXP 2653-138-1) and cured at 100°C for 60 min. Following curing, they were sintered at 427°C for 60 minutes under low-flow argon gas.

Once fabricated, all electrodes were characterized according to the ANSI/AAMI EC12 standard for AC impedance^57^, DC offset voltage, combined offset instability and internal noise, and bias current tolerance (**Supplemental Table 7**). Pairs of electrodes were also connected to the earpiece circuits with clip-on wires and tested on an agar phantom as described in the *Device Testing* section.

#### Electrical Impedance Spectroscopy Validation

Measurement accuracy was first evaluated using a test fixture simulating the load and the four electrodes (**Supplemental Figure 2**). The load was either purely resistive (*R*, 20-100 Ω), or slightly reactive (*2R1C*, 20-100 Ω in parallel with 470 Ω and 22 nF in series) to mimic the brain parenchyma. The electrodes were implemented as either (i) low impedance (270 Ω resistor in series with a 1 kΩ resistor and 180 nF capacitor in parallel), or (ii) high impedance (620 Ω resistor in series with a 3.3 kΩ resistor and 82 nF capacitor in parallel), in order to simulate the frequency-dependence of typical Ag/AgCl electrodes, but also to ensure a minimal impedance at high frequencies. Two 10 Ω resistors were added between the electrodes connected to the same side of the load to simulate the tissue impedance between these electrodes as well. **Supplemental Table 8** describes the benchtop tests performed and **Supplemental Table 9** and **Supplemental Table 10** contain the results.

#### Device Testing

In addition to the EIS test fixture, stability and reproducibility of EIS measurements over twelve hours were evaluated with an agar phantom. A slab (5-10 mm thick) of conductive agar was first cast using a solution of 2.5% weight agar (Living Jin USA, Inc.), 0.64% weight NaCl (RND Center, Inc.) and distilled water. Electrodes (mastoids and earpieces for the final testing) were then coated with conductive gel (Electro-Gel, Electro-Cap International) to reduce the contact resistance, and placed on the agar slab in a pattern geometrically similar to the electrode location on the head. A 12-hour recording was performed, resulting in a total of 450 minutes of EEG recording, 159 EIS cycles and 160 electrode impedance checks. The results of a 12 hr agar phantom EEG test is shown in **Supplemental Figure 3**.

#### Signal Processing – Parenchymal Resistance

A python neuroprocessing pipeline developed by Applied Cognition performed the data analysis of the signals captured by the investigational device. The neuroprocessing pipeline runs in the Amazon Web Services cloud on a distributed architecture allowing for fast parallel execution of participant device readings.

An EIS scan by the investigational device measured impedances Z(ω) at 18 frequencies *f* (ω = 2*πf*) ranging from 1,000 Hz to 256,000 Hz and took 106 seconds. The frequencies were measured in the following permuted order for each scan (all in Hz): 2276, 102400, 1600, 64000, 4551, 51200, 12800, 85333, 3200, 25600, 1000, 32000, 6400, 8533, 128000, 204800, 256000. During the EIS scan, all other device sensors were powered off except the left-in-ear IMU that sampled at 1Hz to detect head motion and position. The EIS scans were duty-cycled throughout the recording period with 165 second neurophysiology scans that captured data from the other sensors: EEG, PPGs and IMUs. Four electrode impedance checks at 30Hz and lasting 7 seconds were performed during the neurophysiology scan and immediately preceding the EIS scan.

The Cole-Cole model^58–60^ is commonly used to analyze EIS data. The analysis is based on the four parameters contained in the Cole equation R_0_, R_∞_, α and τ.

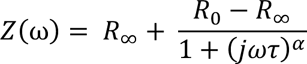

As the excitation frequency ω (ω = 2*πf*) increases to large values, the capacitive cell membranes are invisible to the excitation frequency and the impedance Z(ω) approaches R_∞_. The four excitation and response electrodes are positioned trans-cranially and at large ω measure total intracranial brain volume. As the excitation frequency ω decreases approaching DC values, the capacitive cell membranes prevent any transmembrane conduction and the impedance Z(ω) approaches R_0_ the measure of electrical resistance through extracellular fluid.

The value of τ in the Cole-Cole model is the inverse of tissue characteristic frequency 2*πf_c_*. The Cole-Cole α describes the divergence of a measured dielectric dispersion from the ideal dispersion exhibited by a Debye type of dielectric relaxation, and is widely assumed to be related to a distribution of the relaxation times in the system involved. The value of α ranges from 0.5 to 1 with a value of 1 reducing the Cole-Cole model to the Debye model.

For each EIS scan, the four parameters were estimated using a non-linear least-squares fit (Python SciPy’s least_squares) with the Trust Region Reflective algorithm that is a robust method suitable for large sparse problems with bounds. The α parameter was found to be close to one in the estimations and thereafter set to one, reducing the Cole-Cole model to the Debye model. The remaining three parameters were estimated with bounds of 0 to 90Ω for *R*_0_, 0 to 70Ω for *R* and bounds for 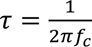 determined by requiring the characteristic frequency *f* to lie between 16kHz and 160kHz. The parameter bounds where confirmed to encompass neurophysiological values in all participants by plotting all EIS scan resistance (real values of impedances Z(ω)) and reactance (imaginary values of impedance Z(ω)). Furthermore, no parameter solutions lay on a boundary for each scan across the range of dynamic impedance spectroscopy measurements for all participants.

Prior to fitting the Debye model to the EIS scan data, to compensate for stray capacitance, time delay effects from signal transmission and electrical components, the impedances Z from each scan were rotated by multiplying by the factor *e*^*j*ω*T*^ with the value *T* chosen so that the reactance at the highest frequency of the scan was zero^59^. This rotation was independently validated using EIS scans recorded on the benchtop tests using known impedance loads (**Supplemental Table 10**). The root-mean-squared-error (RMSE) of the Debye model fits to the benchtop tests (**Supplemental Table 8)** are reported in **Supplemental Table 11**.

Each EIS scan measures R_P_ at a point in time and to evaluate glymphatic function the Debye model fits need to detect small changes in R_P_. With the device time-multiplexing settings, the device completes approximately 84 scans over a 7-hour night of sleep. The statistical models require significance testing of the mean change in R_P_ between sleep and wake, and testing significance of predictors of R_P_ in linear mixed models. These tests depend on the SD of the Debye model dispersion and the number of EIS scans (**Supplemental Table 12**). Details of these analyses are found in **Supplemental 4**.

During EIS, tissue behaves as a linear, time-invariant and causal system allowing the use of Kramers-Kronig relations to identify electrical impedance measures that are corrupted by artifacts from motion, electrode impedances and other sources^31,32^. The Kramers-Kronig transform allows the real part of the electrical impedance measure to be derived from the imaginary part and vice versa^31^. These relations are used as validity tests for measured spectrums. When the real and imaginary impedance measures do not satisfy the Kramers-Kronig relation the measure is discarded. The Python PyEIS1.0.10 repository was used for Kramers-Kronig impedance validation. The validation uses the method of Schönleber^32^ to avoid ambiguities in the linear Kramers-Kronig validity tests due to under- and over-fitting.

In addition to the Kramers-Kronig validity tests, EIS measures were discarded if for any of the four electrode impedances, the difference across successive time measures exceeded 1kΩ or if the EIS scan was collected during a wake interval on a participant randomized to sleep. The first condition ensured that the electrode impedances were maintained within a narrow range during the EIS scans and the second condition ensured that the EIS scans were representative of sleep physiology for participants randomized to sleep.

#### Signal Processing – Electroencephalography

The raw EEG tracings from the device and the commercial PSG (Philips Respironics Alice 6 LDx Diagnostic Sleep System) were time-synchronized by time-aligning a 100-ms square pulse generated by the device every 10 seconds and stored both in the device flash memory and in the PSG recording through as an unused auxiliary analog input channel. Each raw tracing was notch filtered at 60 Hz using a second-order infinite impulse response (IIR) notch digital filter (Python SciPy *iirnotch*). The digital filter was applied to the signal forward and backward with a combined filter phase of zero (Python SciPy *filtfilt*). The signal was then bandpass filtered between 0.3 Hz and 50 Hz using a finite impulse response (FIR) filter with a Hann window (Python SciPy *firwin*). The length of the low-pass filter was 501 and the length of the high-pass filter was 2,401.

During the observation periods, participants were not allowed to touch devices that were plugged to power outlets. Other non-physiologic sources included excessive head motion that led to signal artifacts from the electrode-skin interface. Physiologic sources of signal artifact included electrooculogram, electrocardiogram and electromyogram. The signals where partitioned into time-aligned 30-second epochs. Epochs containing one or more peak-to-peak signal amplitude exceeding 350 µV or maximum power in the Welch power spectrum exceeding 1,000 µV^2^/Hz were removed. In addition, the device left in-ear and right-in ear IMU signals were used to filter excessive motion by filtering epochs with peak-to-peak values on an x-, y-, or z-axis on either ear that exceeded 100 milli-g during sleep and 200 milli-g during wake.

The power spectral density of each 30-second epoch was computed using Welch’s method^61^ (Python Scipy *welch*) using 10-second segments and 50% overlap across successive segments. Relative power bands were computed for delta power (1-4 Hz), theta power (4-8 Hz), alpha power (8-12 Hz), sigma power (12-15 Hz), beta power (15-30 Hz) and low gamma power (30-50 Hz), each normalized to the total power in the power spectral density. Simpson integration of the power spectral density was used for computing the total power and respective band powers.

Hypnogram staging of each 30-second epoch used automated scoring (Python yasa 0.6.3 *SleepStaging*) trained and validated on 3,000 nights of data from the National Sleep Research Resource^62^. The automated scoring uses a single EEG derivative and a single EOG channel. For device hypnogram staging the single in-ear transcranial derivative was used and for the commercial PSG, hypnogram staging using C3-A2 and C4-A1 derivatives were recorded separately. The PSG left EOG channel was used for commercial PSG hypnogram staging. An additional T3-T4 transcranial PSG derivative was used for hypnogram staging as another comparison to the device’s transcranial in-ear derivative. The internal agreement of commercial PSG and agreement between investigational device-based EEG and PSG are presented in **Supplemental Table 6**.

#### Signal Processing – Heart Rate

The photoplethysmography (PPG) left and right raw signals were bandpass filtered between 0.5 Hz and 10 Hz using a finite impulse response (FIR) filter with a Hann window (Python SciPy *firwin*). Empirical mode decomposition was used to identify peaks in the raw signals (Python emd 0.6.2) using a box-car rolling window of 100 observations^63^. Heart rate variability metrics were computed after removing outliers, ectopic beats and interpolating missing values (Python Aura-healthcare *hrvanalysis*). Both time-domain features (standard deviation (SD) of the inter-beat-interval of normal sinus beats, *sdnn*) and frequency-domain features using the Lomb-Scargle periodogram (low-frequency signal power *lf* between 0.04-0.15 Hz considered a mixture of sympathetic and parasympathetic activity, high-frequency signal power *hf* between 0.15-0.4 Hz representing beat-to-beat changes from parasympathetic vagal activity, and the low-frequency to high-frequency ratio (*lf-hf-ratio*) were computed^64^.

The device inertial-measurement units (IMU) were located inside the left and right ear canal and provided left and right cartesian coordinates of acceleration measured in milli-g where 1,000 milli-g is the acceleration caused by earth’s gravitational force. Each in-ear component from the device is uniquely and reproducibly oriented on account of the ellipsoidal shape of the human ear-canal and the matching shape of the in-ear component. From this orientation, head position and motion (acceleration) were resolved from each set of IMU samples that included gross positions such as *supine*, *reclined*, *upright*, *left* and *right*.

### Clinical Study Design

The Investigational Device was evaluated in two studies, a Benchmarking Study conducted in The Villages community in Florida in partnership with the UF Health PHRC, and a Replication Study conducted at the University of Washington in Seattle. These studies were reviewed and approved by University of Florida Institutional Review Board (IRB No. 202201364) and Western Institutional Review Board (IRB No. 20225818), respectively. Written informed consent was obtained from all study participants during a screening visit, prior to any study activities.

#### Participant Recruitment

Following distribution of study flyers in public-facing locations in senior recreation and/or local medical centers, the Benchmarking Study enrolled 34 healthy participants 56-66 years of age and the Replication Study enrolled 14 healthy participants 49-63 years of age. Participants were cognitively intact and had no history of clinical depression, confirmed during screening using the Montreal Cognitive Assessment (MOCA) and Geriatric Depression Scale (GDS). Other exclusion criteria included a history of diabetes, hypertension, coronary artery disease, pulmonary disease, neurological disease, or anxiety. Participants planning travel to alternate time zones within two weeks of study participation were also excluded. Consolidated Standards of Reporting Trials (CONSORT) diagram for the Benchmarking Study and Replication Study is provided in **Figure 3** (bottom), and participant exclusions are detailed in the Results section.

Demographic data including age, sex, APOE4 allele status, MOCA and GDS for the Benchmarking Study, Replication Study, and Combined Studies are provided in **Supplemental Table 1.**

#### Study Protocol

As shown in the study schematic (**Figure 3**, top), this is a cross-over study in which participants took part in two overnight study visits, one undergoing normal sleep and the other undergoing overnight sleep deprivation, at a 2-4 week interval. Each overnight study visit consisted of three study periods beginning at 1900 hrs, 0700 hrs, 1330 hrs separated by two intervals – an overnight interval (2300-0630 hrs), and a morning interval (0800-1130 hrs).

##### Benchmarking Study

Beginning with arrival to the study suite at 1900, participants underwent a blood draw for APOE genotyping and assessment of amyloid β and tau biomarkers (Aβ_1-40_, Aβ_1-42_, phospho-tau181, phosho-tau217, non-phospho-tau181, non-phospho-tau217; C_2_N Diagnostics^65–67^) and 60 min of non-contrast magnetic resonance imaging (MRI) scanning (sequences detailed below). On completion, participants were instrumented for commercial polysomnography (PSG, Philips Respironics Alice 6 LDx Diagnostic Sleep System) that included a 10-20 EEG montage with electro-oculogram (EOG) and submental electromyogram (EMG). Electrode impedances were required to be below 5 kΩ. Participants were last instrumented with the Investigational Device, and device electrode impedances required to be below 5 kΩ.

The Overnight interval lasted from 2300-0630 hrs the following morning. To support study planning, participants were randomized to undergo either normal sleep or sleep deprivation for their initial overnight visit following completion of their informed consent, however this information was not shared with the participants. To reduce likelihood of preparation bias on the part of participants (i.e. altering sleep schedules or taking naps in anticipation of sleep deprivation visits) participants were not informed of their initial visit assignment until 4pm the day of their arrival. Both PSG and Investigational Device recordings took place continuously throughout the Overnight interval. Throughout the overnight period, sleep-assigned participants were allowed to sleep normally without interruption. Participants assigned to the sleep deprivation were monitored by study staff to ensure adherence to the sleep deprivation protocol.

Between 0700-0745 hrs the next morning, all participants were administered a cognitive battery that included the 5-minute psychomotor vigilance test for sustained attention^68,69^, symbol-digit modality test of processing speed, trail making test part A and part B for assessing executive function^70^, and digit span forward test of working memory^71^. Following the cognitive battery, participants underwent a blood draw for amyloid β and tau biomarkers and a second non-contrast MRI scanning session (denoted T_0_ in Results section). Immediately following the second non-contrast MRI scan, participants underwent intravenous gadolinium-based contrast agent (GBCA) injection (Gadavist, 0.1 mmol/kg), followed by T1- and T2-weighted contrast-enhanced MRI scans 7-10 min after GBCA injection (denoted T_10_ in Results section).

Participants were escorted back to the study suite where they were re-instrumented with the commercial PSG and the Investigational Device. The Morning Interval ran from 0800 to 1130. Participants who assigned to the sleep condition in the Overnight Interval were required to remain awake and were monitored by study staff to ensure adherence; those assigned to the sleep deprivation condition in the Overnight were allowed to sleep. During this interval both the PSG and Interventional Device recorded data. The third assessment period commenced at the end of the Morning Interval. Participants underwent a final blood draw for amyloid β and tau biomarker assessment and then had a third MRI scanning session, including non-contrast sequences and T1- and T2-weighted contrast-enhanced sequences at approximately 4 hrs post-GBCA injection (denoted T_240_ in Results section).

##### Replication Study

The Replication protocol was an abridged version of the Benchmarking protocol. The inclusion/exclusion criteria were identical to the Benchmarking study except for the lower age cut-off. Participants were similarly randomized to a night of sleep or a night of sleep deprivation on their first visit, with a crossover to the alternative sleep/sleep deprivation assignment on their second visit. The protocol for the Replication Study was identical to that of the Benchmarking Study excepting that 1) Participants did not undergo commercial PSG, sleep was assessed by Investigational Device EEG; 2) Participant sleep deprivation adherence in the Overnight Interval was assessed retrospectively using the sleep EEG; 3) The cognitive battery did not include the digits forward working memory test; 4) Participants underwent only non-contrast, and not contrast-enhanced MRI.

##### Cognitive Assessments

The 5-minute PVT from Texas A&M University System CSE^72^ and digits forward recall from Cambridge Cognition^71^ were administered on an iPad. The paper-based SDMT from Western Psychological Services^73^, the TMT A and TMT B^74^, were administered by a clinical coordinator trained in administering these tests. The cognitive battery was administered in a quiet room with minimal distractions on the morning following sleep/wake and prior to the neuroimaging and phlebotomy. The administration of each test was preceded by the corresponding practice test. We have included a description of these tests within the overall protocol for completeness. The results of these cognitive tests, and their relationship to Investigational Device outputs, sleep parameters, fluid biomarkers, and MRI measures of glymphatic function will be reported elsewhere.

##### Plasma Biomarkers

The APOE genotyping, amyloid β and tau plasma biomarkers were analyzed using mass spectrometry by C_2_N Diagnostics^75^. The sample collection procedure was provided by C_2_N Diagnostics. Venipuncture and blood draw from the antecubital fossa were performed using a 22-gauge butterfly needle to minimize red blood cell hemolysis. A total of 10 ml of blood was drawn into a K_2_ EDTA Vacutainer. The blood was centrifuged for 15 min using a swinging bucket rotor at 500-700 x g with the brake on. Immediately after centrifugation, four 1.0 mL plasma samples were aliquoted into four Sarstedt 2.0 ml Micro Tube without disrupting the plasma/cell interface when transferring plasma. A calibrated air-displacement hand-held pipette with a polypropylene pipette tip was used. After aliquoting plasma into the Sarstedt Micro Tubes, the tubes were immediately capped and frozen at −40°C. When the tubes were ready to be shipped to C_2_N Diagnostics, they were packed into a plastic zip-lock bag with plenty of dry ice, placed in an absorbent towel and cryobox, and express couriered to C_2_N Diagnostics priority overnight. In the present report, we utilize APOE4 allele status as a covariate in our analysis, and we have included a description of plasma biomarker assessment within the overall protocol for completeness. However, the fluid biomarker levels, and their relationship to Investigational Device outputs, sleep parameters, cognitive tests, and MRI measures of will be reported elsewhere.

#### Magnetic Resonance Imaging Approach

MRI scanning at The Villages site was conducted on a 3T Siemens Vida system at Lake Medical Imaging, while scanning at the University of Washington site was conducted on a 3T Philips Ingenia Elition X at the Diagnostic Imaging Sciences Center. In the present report, to define the relationship between brain R_P_ and glymphatic function, we utilize CE-MRI from Benchmarking Study participants following the overnight sleep condition. We have included a description of the other MRI-based approaches within the overall protocol for completeness. However, the relationship between these different MRI measures of glymphatic function, Investigational Device outputs, sleep parameters, cognitive tests, and fluid biomarker levels will be reported elsewhere.

##### Contrast-Enhanced MRI

Contrast-enhanced MRI involves intravenous administration of a Gadolinium chelate that shortens T1, T2, and T2* relaxation parameters of the tissue it traverses through. Specifically, in this study, shortening of the T1 relaxation parameter enhances MRI signal that is proportional to the amount of Gadolinium and its characteristic relaxivity. Gadolinium passes quickly through the cerebral and peripheral vasculature within the first 5-30 minutes. A fraction of the contrast enters the CSF and ISF^14,15^ over many hours and finally almost 99% of the contrast exits the body by renal clearance. The passage of intravenous GBCA from the vasculature to the CSF and ISF was measured in terms of a delayed signal enhancement at approximately 4 hours after administration. Two prior studies have shown that delayed enhancement following contrast injection between 3 and 6 hours is indicative of solute (in this case, GBCA) transport via the CSF-ISF glymphatic exchange^14,15^. At the Villages®, we performed 3D T1-weighted magnetization-prepared rapid acquisition with gradient echo (MPRAGE) with TR/TI/TE = 2300/900/2 ms, resolution = 1×1×1 mm^3^, field-of-view = 256×256×208 mm^3^, total acquisition time = 5:12 min before contrast injection. The post-contrast MRI image (T_10_) was acquired approximated 7-10 minutes after contrast injection and its acquisition was identical to the pre-contrast image. A second post-contrast scan was acquired about 4 hours (T_240_) later using identical imaging parameters.

##### Structural MRI

Structural T1 MRI was used for classifying the brain tissue into lobar gray matter, lobar white matter, and CSF as well as for segmenting regions-of-interests including the hippocampus, sagittal sinus, internal carotid arteries, lateral ventricle, and subarachnoid space. The T1 images were also used for registration purposes for all other imaging modalities. The T1 images were used for determining PVS morphometry^76,77^. Scanning parameters for the Benchmarking Study are identical to the contrast-enhanced MRI. Scanning parameters for the Replication Study were TR/TI/TE = 12/1000/4.5 ms, resolution = 0.8×0.8×0.8 mm^3^, field-of-view = 256×240×166 mm^3^, total acquisition time = 5:24 min. Since inclusion of T2 MRI significantly improves PVS detection, a T2 MRI with TR/TE = 3200/561 ms, resolution = 0.8×0.8×0.8 mm^3^, field-of-view = 256×240×166 mm^3^, total acquisition time = 4:32 min was also added.

##### Intravoxel Incoherent Motion MRI

Intravoxel incoherent motion (IVIM) MRI is a multi-shell diffusion imaging approach to measure water movement at different scales of distance and time. Glymphatic transport includes movement of CSF in the subarachnoid space and the interstitium at a slow speed (∼1 mm/s) over large distances (∼100-200 um)^1,30,40^. This perivascular movement of CSF through the subarachnoid compartment is upstream of its exchange with the ISF along the penetrating intraparenchymal vasculature and hence could serve as an important correlate of glymphatic function^78^. The imaging parameters for the Benchmarking Study were: TR/TE = 5500/124 ms, resolution = 1×1×5 mm^3^, field-of-view = 250×250×150 mm^3^, parallel imaging factor = 2, multi-band factor = 3, fat suppression with Spectral Presaturation with Inversion Recovery (SPIR), b values = 0, 50, 100, 150, 200, 250, 300, 350, 400, 500, 700, 800, and1000 s/mm^2^, and total acquisition time = 7:01 min. The b = 0 s/mm^2^ was acquired with reverse phase encoding to correct for field distortions. The imaging parameters for the Replication Study were TR/TE = 2978/90 ms, resolution = 1×1×5 mm^3^, parallel imaging factor = 2.2, field-of-view = 230×196×115 mm^3^, fat suppression with SPIR, b values = 0, 10,40,80,100,150,200, 300,500,700, 800, 900,1000 s/mm^2^, and total acquisition time = 8:08 min. A Split echo (SPLICE) acquisition was used to eliminate the need for distortion correction.

##### Multi-Echo, Multi-Delay Arterial Spin Labeling (ASL) MRI

Multi-echo, multi-delay ASL measures perfusion and time for exchange of water across the endothelium. This time metric is dependent on perfusion, endothelial permeability, and the glial vascular unit including the AQP4 water channels that are critical to the perivascular CSF-ISF exchange^78,79^. This protocol was implemented only for the Replication Study as a Hadamard-encoded ASL acquisition, TR/TE = 5000 ms, resolution = 1×1×5 mm^3^, field-of-view = 230×196×115 mm^3^, post labeling delay = 650, 950,1210,1510,2083,2383 ms with background suppression, label duration = 3400 ms, and TE = 0,40,80, 120 ms. Total acquisition time = 12:40 min. A conventional arterial spin labeling (ASL) protocol was implemented in the Benchmarking Study due to the lack of availability of the multi-echo component. The parameters were: TR/TE = 4420/22 ms, resolution = 1.7×1.7×4 mm^3^, field-of-view = 220×220×96 mm^3^, post labeling delay = 500, 900,1000,1100,1200,1400,1600,1800, label duration = 1800 ms. Total acquisition time = 2:37 min. The sequence was acquired twice. A reference M0 images was acquired in both studies with identical parameters but without background suppression or labeling.

##### Fast Functional MRI

A sub-second functional MRI sequence was implemented to separate and regress physiological noise (cardiac and respiratory signals) as well as to capture slow CSF oscillations in the ventricles^80^. The protocol for the Benchmarking Study was TR/TE = 400-500/30 ms, resolution = 3.6×3.6×3.6 mm^3^, flip angle = 70°, multi-band factor = 4, field-of-view = 230×2430×101 mm^3^, fat suppression, no. of volumes = 365 using SPIR, total acquisition time = 2:37 min. Heart rate and respiration were monitored using the BIOPAC MR160 and synchronized with MRI acquisition. The Replication Study used a gradient-echo, echo planar imaging sequence with TR/TE = 400-500/30 ms, resolution = 3×3×4 mm^3^, flip angle = 42°, multi-band factor = 6, field-of-view = 240×240×123 mm^3^, slice gap = 0.4 mm, fat suppression using SPIR, no. of volumes = 450, total acquisition time = 3:00 min. Two single volume, spin echo acquisitions were also acquired with opposite phase encoding to allow for distortion correction. Heart rate and respiration were monitored using the sensors built into the Philips scanner and synchronized with MRI acquisition. Glymphatic CSF-ISF exchange is at least partly driven by vasomotion, which may be reflected by periodic motion in the CSF spaces. This CSF motion correlates well with sleep stages, with power of the slow CSF oscillation increasing from wakefulness to the N1 and finally to the N2 stage^80^.

##### Phase-Contrast MRI

The phase contrast MRI is used to quantify CSF efflux from the Aqueduct of Sylvius. The peristaltic motion of CSF at the Aqueduct is measured by gating the MRI signal to the participants and calculating the CSF motion into and out of the brain over a cardiac signal. Variations in CSF efflux velocity appear to follow the cardiac and respiratory signals and may impact glymphatic CSF-ISF exchange. The Benchmarking Study protocol for this MRI was the following: a single slice acquisition perpendicular to the Aqueduct of Sylvius, TR/TE = 21/6.6 ms, resolution = 30.6×0.6×6 mm^3^, flip angle = 10°, 40 phases per cardiac cycle, and velocity encoding for 12 cm/s, total acquisition time: 2:00 min. The Replication Study protocol included the following: a single-slice acquisition perpendicular to the Aqueduct of Sylvius, TR/TE = 12/7.9 ms, resolution = 30.6×0.6×4 mm^3^, flip angle = 10°, 15 phases per cardiac cycle, and velocity encoding for 12 cm/s, total acquisition time: 2:02 min

#### Statistical Approach

The Benchmarking study was designed to have 80% power at a significance level of 5% to detect a low-to-moderate correlation *r* (*r*=0.5) between the device measurements and CE-MRI measurement of glymphatic function. The Replication study was designed to have 80% power at a significance level of 20% to detect a low-to-moderate correlation *r* (*r*=0.5) between the device measurements and non-contrast MRI measurement of glymphatic function and to detect overnight sleep/wake differences in brain parenchymal resistance.

The parenchymal resistances R_P_ computed from each EIS scan from the device were normalized to the sleep/wake onset values to allow comparison across visits and participants. To reduce measurement variability of the sleep/wake onset value, the value was inferred from a linear regression to the initial R_P_ values. The overnight change in R_P_ per participant visit was taken as the average of the normalized values for that visit. For example, a value of 1 indicated that there was no overnight change from the onset value and a value of 0.8 indicated a 20% overnight reduction from onset. A Student’s T-test group comparison between sleep/wake visits, group mean and SD were calculated.

Contrast-enhanced MRI of sleep-related glymphatic function was analyzed using a random intercept linear mixed effect model (R, lme in the lmne package). Glymphatic function was defined by measuring brain parenchymal contrast enhancement, the % change in T1-weighted signal intensity between 10-240 min post-GBCA injection (100% * (T_240_-T_10_)/T_0_) at each of eight regions of interest (ROIs): frontal cortical gray and white matter, parietal cortical gray and white matter, temporal cortical gray and white matter, and occipital gray and white matter. Only sleep visits were analyzed leading to a one level of grouping, the eight ROIs, for the participant sleep visit. All models included regressors in the mixed model for the T_10_-T_240_ change in blood signal within the internal carotid artery, and the T_10_-T_240_ change in CSF signal within the cerebral lateral ventricles to compensate for the influence of vascular and CSF contrast on parenchymal enhancement. Potentially confounding biological variables age, gender, and APOE ε4 status were also included in all models.

To evaluate different sleep-related contributions to glymphatic function, overnight changes from sleep onset for each of the four recorded EEG relative band powers (delta, theta, alpha, beta) and for HR were computed. The recorded values were normalized to their respective value at sleep onset and a single statistic, the average of the overnight normalized values, computed per participant visit. The four hypnogram sleep stages (N1, N2, N3, REM) and WASO were each summed into single total sleep stage durations and total WASO duration per participant visit. The device R_P_ normalization and averaging has been previously described. Models were run separately for each of the four EEG relative band powers (delta, theta, alpha, beta), for each of the four hypnogram sleep stages (N1, N2, N3, REM), for WASO, for HR and for device R_P_. Two full models were then fit to the data, with the EEG band power and the hypnogram sleep stages separated into distinct models because of collinearity across those predictors. The variance inflation factor (R ‘vif’ function from the ‘car’ package) was used to test for predictor collinearity using a threshold of 5. The EEG powerbands included were limited to delta, theta and beta because of collinearity among the four relative band powers. HR and the device R_P_ were included in both full models. Each full model was reduced to a selected model using backward elimination of predictors with a threshold Wald’s p-value of 0.05.

Model selection stability, or robustness of the selected model to perturbations in the dataset, was tested using resampling-based multi-model inference^29^. The dataset was bootstrapped resampled with replacement to generate 500 bootstrapped datasets. For each of these datasets the two full models were estimated and reduced to a selected model using backward elimination. The bootstrap inclusion frequency (the percent of bootstrap datasets that included the predictor), the RMSD ratio (the RMSD of the predictor bootstrap estimate divided by the standard error of that coefficient in the full model and expressing the variance inflation or deflation caused by variable selection), the relative conditional bias (quantifying the variable-selection-induced bias to expect if a predictor is selected) and the bootstrap median of each predictor coefficient were reported.

The analyses of device R_P_ values with EEG band powers, heart rate and hypnogram staging were performed on the overnight sleep recordings. Analyses were grouped into REM sleep and NREM sleep (N1, N2 and N3). The device resistances were computed repeatedly throughout the night from each EIS scan. During sleep recordings, these data were non-stationary. After normalization to sleep onset as previously described, first-order differences ΔR_P_ and ΔX for X one of the EEG band powers or HR were incorporated into individual random intercept linear mixed models to evaluate their relationship. Threshold regression models were used in the linear mixed model. A custom estimation procedure described in **Supplemental Information** was used to jointly estimate the threshold regression for the full positive and negative range of the predictors. The hinge threshold model was analytically reduced to estimation by linear regression which allowed it to be used in a random intercepts linear mixed effects regression model. The autocorrelation function and augmented Dickey Fuller tests were used to confirm stationarity of the differenced variables ΔR_P_ and ΔX and also of the fitted model residuals (R, acf and adf functions).

In addition to the analyses of ΔR_P_ and ΔX for X one of the EEG powerbands or HR during REM sleep and NREM sleep, these were analyzed at hypnogram transitions NREM to REM, REM to NREM, sleep to wake and wake to sleep. The analysis was similar, using threshold regression models in a random intercept mixed linear model.

Univariate outliers were trimmed at the 0.5 and 99.5 percentile. For multivariate analysis, the Mahalonobis distance was computed and outliers trimmed at the 0.5 and 99.5 percentile of the Mahalonobis distance. All regressions were adjusted for potential confounding effects of age, sex and APOE genotype. All tests were considered significant at a 5% level if the 95% confidence interval (CI) did not contain the null hypothesis. All statistical analyses were performed in R version 4.2.1 (2022-06-23) or Graphpad Prism 8.0.0.

We reported the pooled findings of the Benchmarking and Replication study for both the investigational device transcranial resistance measurements and EEG parameters.

## SUPPLEMENTAL INFORMATION

### Supplemental 1: Analysis of device EEG and commercial polysomnography

The single derivative device EEG was validated against the commercial PSG (Philips Respironics Alice 6 LDx Diagnostic Sleep System) in participants that underwent concurrent device and PSG sleep recording as part of the Benchmark study. Both hypnogram sleep staging and sleep parameters were validated using a confusion matrix between the device prediction and the PSG prediction both per participant and pooled across participants. Prediction metrics computed included sensitivity, specificity, precision, accuracy and F1 score. The device EEG with EOM and the PSG C3-A2, C4-A1 derivatives were individually input into the automated hypnogram scoring (Python yasa 0.6.3 *SleepStaging*). Manual scoring of the PSG by a sleep physician (YC) was used as the gold-standard. The results of these comparisons are provided in **Supplemental Table 6.**

### Supplemental 2: Threshold regression model and estimation

Threshold regression models were fit on data that spanned the full positive and negative range of abscissa values. These models are defined by a positive turning point *t_p_* ≥ 0 where for *x* ≥ *t_p_* the regression line goes from *f*(*x*) = 0 to *f*(*x*) = *m_p_*(*x* – *t_p_*) and a negative turning point *t_n_* ≤ 0 where for *x* ≤ *t_n_* the regression line goes from *f*(*x*) = 0 to *f*(*x*) = *m_n_*(*x* – *t_n_*). These models can be reduced to linear regression. First transform the data (*x*, *y*) → (*x*, *y*) were

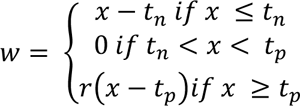

For values of *t_p_*, *t_n_* and *r*, a least squares estimator can be used to fit a linear model to the transformed data values (*w*, *y*). With 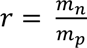, the transformed data is linear, through the origin and has slope *m_n_*. When *r* = 1 both slopes are identical; for *r* ≠ 1 the positive half-space slope equals the regression estimate multiplied by *r*. Hence, for *r* < 1 the positive half-space slope is less than the negative half-space slope and for *r* > 1 the positive half-space slope is greater than the negative half-space slope.

The linear fit was estimated using an F-statistics, the ratio of the explained variance (due to the model) and the unexplained variance (residuals). Grid search over the parameter space *t_p_*, *t_n_* and *r* was carried out and the values that minimized the mean sum of squared residuals (and hence maximized the F-statistic) was reported^81^. To avoid edge effects, *t_p_* and *t_n_* were set to *±1.5* standard deviations^82^. Random intercept linear mixed models were similarly fit to the transformed data and grid search used to solve for the parameters that minimized the mean sum of squared residuals for the fixed effects. The autocorrelation function was used to confirm no residual autocorrelation following model estimation and fit.

### Supplemental 3: Physiological basis for electrical impedance spectroscopy change with glymphatic function

In normal adult men (women) between the ages of 60-86 years, the GM, WM and total intracranial volumes (TIV) are 614.3 ± 60.1 (562.6 ± 50.1 ml), 534.0 ± 57.2 ml (476.6 ± 44.7 ml) and 1603.0 ± 133.0 ml (1425.9 ± 109.0 ml)^83^. Average adult cerebral blood flow (CBF) is approximately 750 ml/min or 12.5 ml/sec. Assuming HR of 60 bpm, 12.5 ml of blood injected into the brain causes an approximate 50 mΩ decrease in the device IPG sampled at 80 kHz (see for example **Figure 2d**, IPG-Impedance Plethysmograph data taken from a female study participant). At 80 kHz, EIS measures the extracellular compartment and most of the intracellular compartment. Thus, the 12.5 ml increase in blood volume, representing a 12.5/1425.9 = 0.88% change in TIV, would be measured as a 50 mΩ decrease in resistance by EIS.

At low frequencies, EIS measures changes in extracellular volume. The interstitial cortical volume fraction α in awake rodents has been shown to be 14.1% and in sleeping rodents 23.4% whereas the tortuosity λ of the interstitial space remained unchanged across sleep and wake^2,84^. This suggests that in rodents, the sleep-induced volume expansion of the interstitial space occurs from a change in the cross-sectional area and not length. This is noteworthy because resistance of a conductor is proportional to 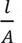 where 𝑙 is the length and *A* the cross-sectional area. Therefore, the increase in interstitial space fluid with glymphatic function will increase the cross-sectional area of the interstitial space channels and proportionally decrease the transcranial electrical resistance. In humans, the measurements range from 15-20%^85^ but changes with sleep/wake have not been assessed. Using the rodent data as a proxy to what we might expect in humans, we would expect a 66% increase in interstitial volume with sleep to occur from of a 66% increase in cross-sectional area of the interstitial channels. The remaining extracellular compartment volumes contributing to the resistance at low EIS frequencies are CSF and blood plasma that occupy a total volume comparable to the awake interstitial volume. Therefore, the net extracellular compartment volume increase, or effective cross-sectional area increase, with sleep is 33%. Using the IPG relationship between fluid volume change and transcranial impedance, a 0.88% increase in fluid volume was measured as a 50 mΩ decrease in transcranial electrical resistance implying that a 33% volume increase would be measured as an 1875 mΩ decrease. This analysis uses the conclusion that the interstitial volume increase of glymphatic function is an increase in the interstitial channel cross-sectional area and similarly the assumption that the intravascular volume increase of cardiac output is from an increase in the intravascular cross-sectional area. The preceding analysis assumed a 66% increase in interstitial volume fraction as seen in rodents. Assuming a smaller 25% volume fraction increase and using the same analysis, we would expect a 710 mΩ decrease in transcranial resistance with sleep.

### Supplemental 4: Benchtop and agar phantom characterization of electrodes and device

Electrodes were characterized according to the ANSI/AAMI EC12 standard for AC impedance, DC offset voltage, combined offset instability and internal noise, and bias current tolerance^57^. Defibrillation overload recovery was not tested as it is not relevant to the present application. Pairs of electrodes were immersed in a 140 mM NaCl solution, facing each other and separated by 3-4 mm. AC impedance at 10 Hz was measured using a multifunction DAQ module (Analog Discover 2, Digilent) configured for 2-point impedance measurement. An excitation voltage of 100 millivolts peak-to-peak (mVpp) and a 10 kΩ series resistor guaranteed a maximum current of 10 microamps peak-to-peak (μApp). DC offset voltage was measured with a Fluke 87V multimeter. Offset instability and internal noise were measured over five minutes using a 24-bit digitizer module (ADS1299EVM, Texas Instruments, 67 nV resolution) sampling at 2000 samples per second, calibrated using a Keysight EDU33212A Signal Generator. Recorded data was subsequently filtered with a 0.15-100 Hz first order bandpass (simulating EC12 filtering) or the 0.3-50 Hz EEG bandpass filter described in **Signal Processing – Electroencephalography**. Peak-to-peak values over the 5-minute period are reported. Bias current tolerance was measured with a multifunction DAQ module using its 14-bit digitizer (0.32 mV resolution) as data logger (sampling every 10 seconds for 8 hours), and its power supply to apply 200 nA DC across the pair of electrodes (2 V across a 10 MΩ resistor in series). The maximum voltage deviation relative to the initial offset voltage before current application is reported. All results are found in **Supplemental Table 7**.

Benchtop validation of the investigational device EIS compared the measured to the theoretical resistance and reactance at each frequency on a circuit model of brain parenchymal impedance (the load) with separate models for each of the four electrode impedances. The circuit diagram for these benchtop measurements is shown in **Supplemental Figure 2**. The load was modelled using 2R1C parallel circuit and the four electrode-skin interfaces were modeled by individual 2R1C circuits with a charge transfer resistance in parallel to a double layer capacitance that was in series with a resistor embodying the electrolyte and stratum corneum resistance. The resistance and capacitance values for the load were based on observed participant resistance and reactance over the EIS frequencies. Two sets of parameters for the electrode impedance models were used, a low-impedance model and a high-impedance model. These were selected to cover the observed range of subject electrode impedances over the range of EIS frequencies. The parameters for the complete set of benchtop tests are found in **Supplemental Table 8** and included (i) calibration measurements of purely resistive loads, (ii) calibration of a 2R1C load varying the parallel resistor R_s_ to simulate changes in the interstitial space resistance, and (iii) calibration of a fixed 2R1C load while varying the electrode impedances at each electrode between low and high impedance settings to test the sensitivity to expected variations during use. The absolute error between the measured and theoretical resistance and reactance for the benchtop tests are reported in **Supplemental Table 9** and in **Supplemental Table 10**, respectively. The measured impedances reported were rotated by multiplying by the factor *e*^−*j*ω*T*^ with the value *T* chosen so that the reactance at 256kHz was zero^59^. The absolute error of the resistance reveals a systematic positive bias implicating contributions to the resistance in the device or benchtop setup that were not fully compensated by the device calibration. For the measurements of the 2R1C benchtop tests, the resistance bias is 0.278 Ω and the bias unadjusted and adjusted RMSE are 0.321 Ω and 0.144 Ω, respectively. The reactance did not show a systematic bias. The RMSE of the reactance for the 2R1C benchtop tests is 0.128 Ω. Because we are interested in the dispersion of the resistance over frequencies, or the difference between the resistance at a low frequency and at a high frequency, the bias cancels out. Using the signal processing pipeline to fit the Debye model, **Supplemental Table 11** reports the dispersion prediction error from theoretical value at each frequency relative to 128 kHz. Reported values are for the 2R1C electrode impedance sensitivity tests. The 1 kHz minus 128kHz dispersion is representative of the parenchymal resistance R_P_ computed from the difference between the extrapolated DC resistance and the infinite frequency resistance. This dispersion had an RMSE of 0.147 Ω approaching the bias-adjusted RMSE of the 2R1C benchtop tests in **Supplemental Table 9**, and represents a 3.1% measurement error in the theoretical dispersion value of 4.766 Ω.

The device EIS showed the accuracy needed to reliably measure a single R_P_ value but to evaluate glymphatic function it needs to detect changes in R_P_. These changes are small by comparison to the absolute value of R_P_. As discussed, the overnight decrease ranges between 500 mΩ and 2000 mΩ. With the current time-multiplexing settings, the device completes approximately 84 scans over a 7-hour night of sleep, or 6 to 23 mΩ average decrease between scans. The statistical models used in the analysis of the Benchmarking Study and Regression Study required significance testing of the mean change in R_P_ between sleep and wake, and testing significance of predictors of R_P_ in linear mixed models. These tests depend on the SD of the Debye model dispersion estimate which for the electrode impedance sensitivity tests was 0.113 Ω, slightly less than the bias-adjusted RMSE. **Supplemental Table 12** shows that the minimum change in R_P_ with 25 and 100 EIS scans that is statistically significant is 44 mΩ and 22 mΩ, respectively. Note that during sleep, R_P_ decreases with glymphatic function and therefore each measurement arises from a distribution with a different mean but the same SD *σ*. Using the linearity of expectation and statistical independence of each R_P_ measurement, the standard error of the mean of R_P_ is 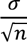 for *n* samples. Using the Fisher transform^86^ to solve for the minimum significant correlation detectable at a given sample size, **Supplemental Table 11** reports that weak to moderate correlations of R_P_ with predictors will be significant at 100 and 25 EIS scans, respectively. These estimates are optimistic since not all sources of error are accounted by the electrode impedance sensitivity tests.

Agar phantom testing over 12 hours was used as a final quality check for each fully assembled device. These tests measured the device combined noise level from the electronics, electrochemical interfaces between the electrodes, EEG gel and agar. **Supplemental Figure 3** shows the device periodogram, EEG recording and power spectral density from an agar phantom test.

## Data availability

The data supporting the results in this study are available from the corresponding author with Institutional Review Board approval and Data Use Agreement permitting non-commercial use of data for independent validation, publication and sharing of new findings.

## Code availability

Code used for the analysis and to produce figures are available upon reasonable request to the corresponding author.

---

**Supplemental Table 1.**
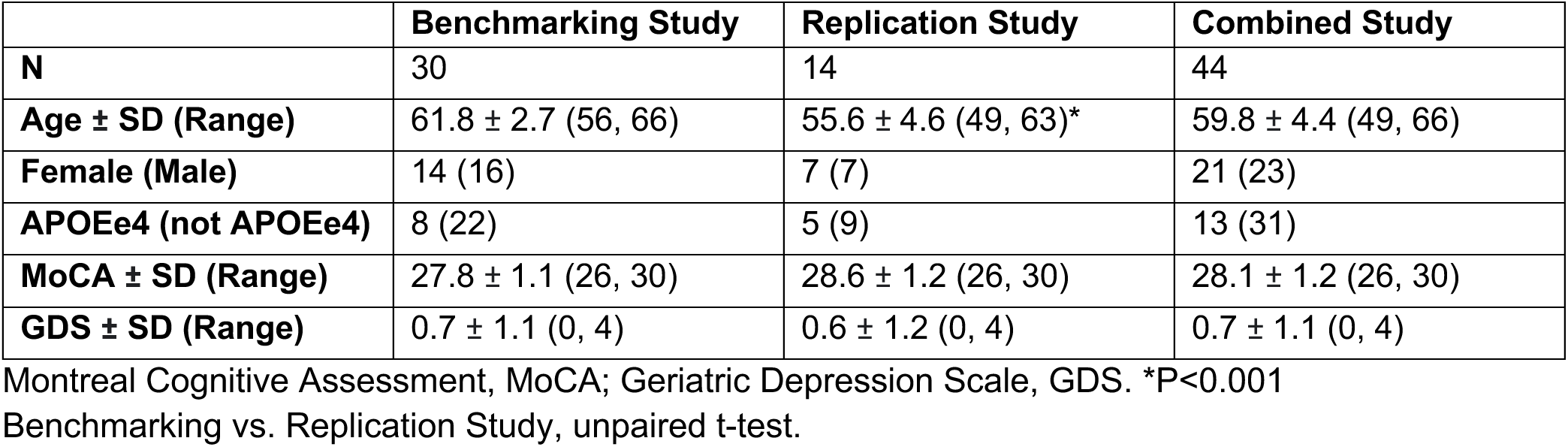
Demographic, genetic and psychometric results of the Benchmarking Study, Replication Study and Combined Study.

|  | **Benchmarking Study** | **Replication Study** | **Combined Study** |
| --- | --- | --- | --- |
| **N** | 30 | 14 | 44 |
| **Age ± SD (Range)** | 61.8 ± 2.7 (56, 66) | 55.6 ± 4.6 (49, 63)* | 59.8 ± 4.4 (49, 66) |
| **Female (Male)** | 14 (16) | 7 (7) | 21 (23) |
| **APOEε4 (not APOEε4)** | 8 (22) | 5 (9) | 13 (31) |
| **MoCA ± SD (Range)** | 27.8 ± 1.1 (26, 30) | 28.6 ± 1.2 (26, 30) | 28.1 ± 1.2 (26, 30) |
| **GDS ± SD (Range)** | 0.7 ± 1.1 (0, 4) | 0.6 ± 1.2 (0, 4) | 0.7 ± 1.1 (0, 4) |

Montreal Cognitive Assessment, MoCA; Geriatric Depression Scale, GDS. \*P<0.001 Benchmarking vs. Replication Study, unpaired t-test.

**Supplemental Table 2.** EEG band power, hypnogram, and R_P_ as individual predictions of parenchymal contrast enhancement when added to the null model following overnight sleep.

|  | Predictor | Estimate | 95% CI | P Value | LRT | LRT P Value |
| --- | --- | --- | --- | --- | --- | --- |
| Predictors Added to Null Model | Delta Band Power | 19.121 | (5.482, 32.760) | **0.007** | 7.50 | **0.006** |
|  | Theta Band Power | -3.755 | (-11.982, 4.473) | 0.378 | 0.81 | 0.368 |
|  | Alpha Band Power | -7.279 | (-12.264, -2.294) | **0.005** | 8.12 | **0.004** |
|  | Beta Band Power | -5.329 | (-7.786, -2.873) | **<0.001** | 17.49 | **<0.001** |
|  | Heart Rate | -21.624 | (-49.814, 6.566) | 0.139 | 2.28 | 0.131 |
|  | WASO Time | -0.003 | (-0.045, 0.038) | 0.881 | 0.02 | 0.879 |
|  | N1 Sleep Time | -0.128 | (-0.213, -0.042) | **0.004** | 8.46 | **0.004** |
|  | N2 Sleep Time | -0.031 | (-0.071, 0.009) | 0.139 | 2.28 | 0.131 |
|  | N3 Sleep Time | 0.025 | (-0.024, 0.075) | 0.322 | 1.02 | 0.313 |
|  | REM Sleep Time | 0.031 | (-0.023, 0.086) | 0.268 | 1.28 | 0.258 |
|  | Parenchymal Resistance | -38.080 | (-61.348, -14.812) | **0.002** | 10.15 | **0.001** |
| Null Model Fixed Effects | Blood CE | 10.29 | (5.45, 15.13) | **<0.001** | - | - |
|  | CSF CE | 148.48 | (123.40, 173.56) | **<0.001** | - | - |
|  | Age | -1.31 | (-1.91, -0.71) | **<0.001** | - | - |
|  | Male Sex | -2.69 | (-5.42, 0.063) | 0.060 | - | - |
|  | APOE ε4 | -3.29 | (-6.50, -0.091) | **0.048** | - | - |
| Null Model Random Effects | σ<sup>2</sup> | 70.50 | - | - | - | - |
|  | T00 ROI | 12.60 | - | - | - | - |

Wake After Sleep Onset, WASO; Rapid Eye Movement, REM; Confidence Interval, CI; Likelihood Ratio Test, LRT. **Bolded P values reflect significant associations at P < 0.05.** Likelihood ratio test taken between null model and model with predictor.

**Supplemental Table 3.**
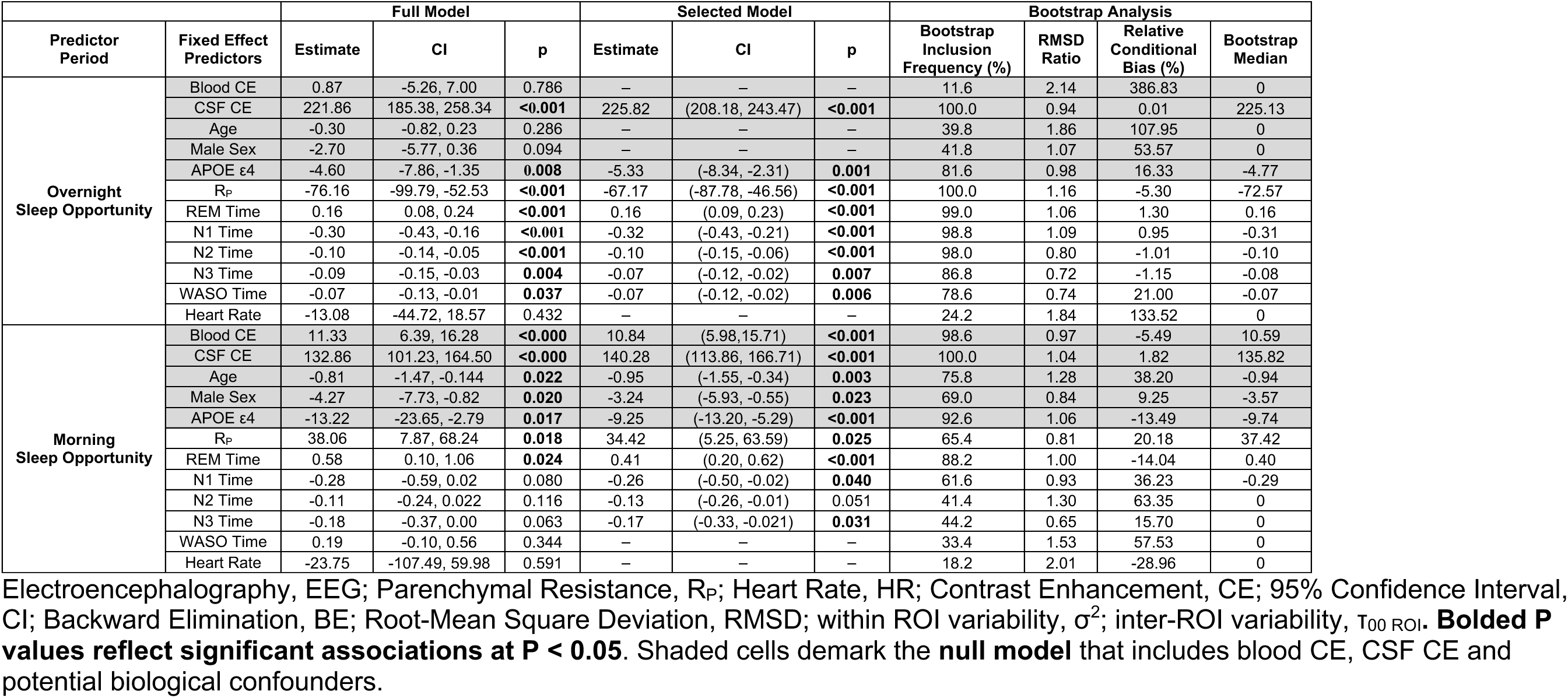
Multivariate models of morning brain parenchymal contrast enhancement using overnight and morning Rp, HR and EEG hypnogram predictors.

**Supplemental Table 4.**
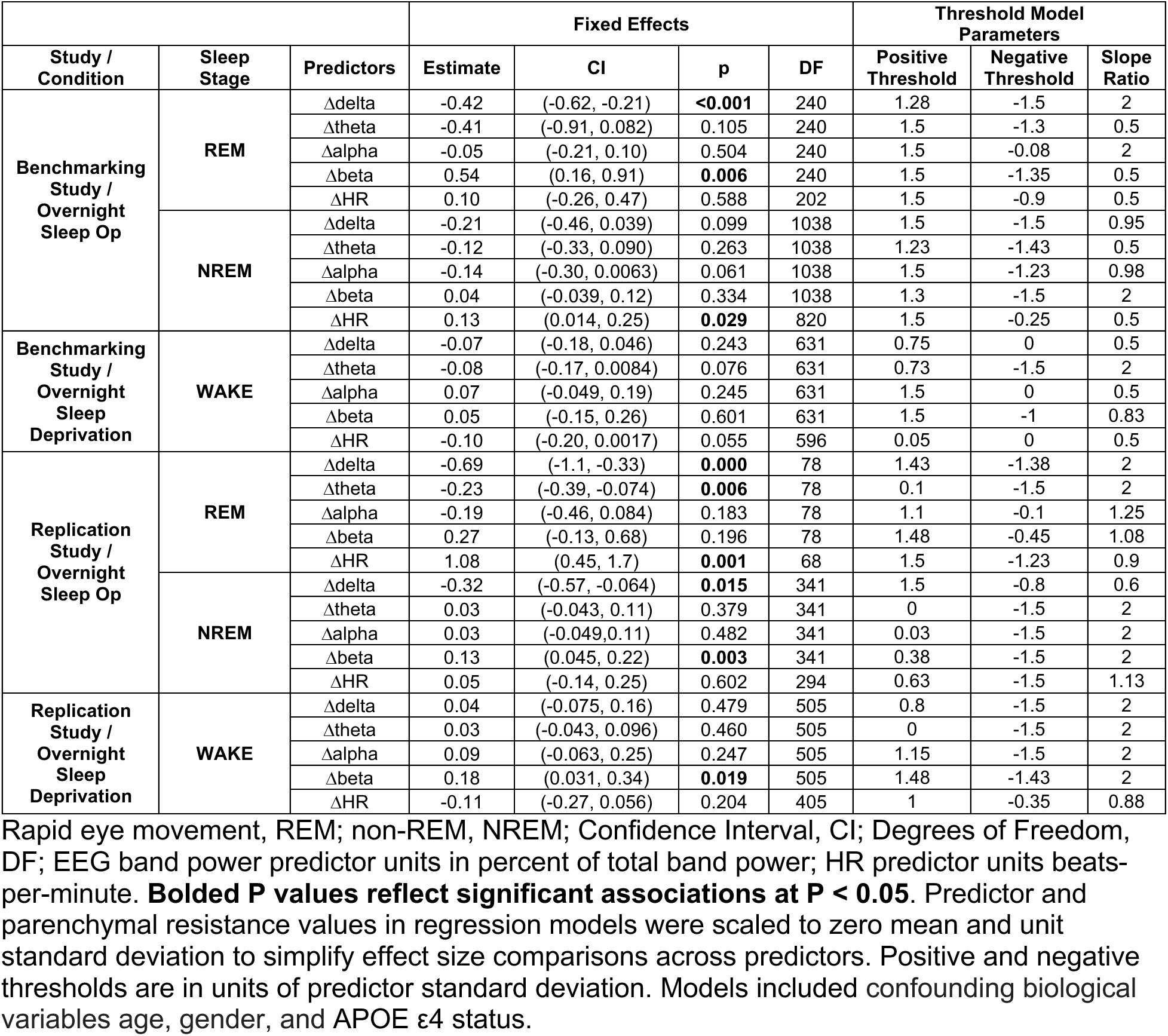
The change in parenchymal resistance resulting from large changes in EEG band power and heart rate during REM sleep, NREM sleep and wake for each study.

**Supplemental Table 5.** The change in parenchymal resistance resulting from large changes in EEG band power and heart rate at NREM-REM and sleep-wake transitions for each study.

| Study | Transition | Fixed Effects |  |  |  |  | Threshold Model Parameters |  |  |
| --- | --- | --- | --- | --- | --- | --- | --- | --- | --- |
|  |  | Predictors | Estimate | CI | P | DF | Positive Threshold | Negative Threshold | Slope Ratio |
| Benchmarking Study | NREM to REM | $\Delta\alpha$ | 0.86 | (-0.05, 1.77) | 0.073 | 52 | 0.725 | -1.5 | 0.5 |
| | | $\Delta\beta$ | 1.01 | (0.18, 1.85) | **0.023** | 52 | 0.35 | -1.5 | 0.5 |
| | | $\Delta\delta$ | -0.31 | (-0.96, 0.34) | 0.359 | 52 | 1.5 | -1.15 | 2 |
| | | $\Delta\text{HR}$ | 0.10 | (-0.27, 0.47) | 0.593 | 37 | 1.5 | 0 | 2 |
| | | $\Delta\theta$ | 0.28 | (-0.37, 0.93) | 0.405 | 52 | 0 | -1.5 | 0.5 |
| | REM to NREM | $\Delta\alpha$ | -0.17 | (-0.46, 0.13) | 0.276 | 45 | 1.5 | 0 | 2 |
| | | $\Delta\beta$ | -0.08 | (-0.29, 0.13) | 0.445 | 45 | 0.2 | -1.5 | 1.9 |
| | | $\Delta\delta$ | -0.43 | (-0.74, -0.11) | **0.011** | 45 | 1 | -0.9 | 2 |
| | | $\Delta\text{HR}$ | 0.32 | (-0.13, 0.78) | 0.180 | 34 | 1.5 | -0.625 | 2 |
| | | $\Delta\theta$ | -0.03 | (-0.17, 0.1) | 0.628 | 45 | 0 | 0 | 2 |
| | Sleep to Wake | $\Delta\alpha$ | -0.89 | (-1.71, -0.08) | **0.038** | 53 | 0.775 | -1.5 | 0.5 |
| | | $\Delta\beta$ | 0.67 | (-0.14, 1.47) | 0.114 | 53 | 0.35 | -1.5 | 0.5 |
| | | $\Delta\delta$ | 0.03 | (-0.17, 0.23) | 0.778 | 53 | 0.025 | -1.45 | 2 |
| | | $\Delta\text{HR}$ | -0.32 | (-1.01, 0.38) | 0.383 | 39 | 0 | -1.4 | 0.5 |
| | | $\Delta\theta$ | 0.04 | (-0.2, 0.29) | 0.737 | 53 | 0.25 | -1.5 | 1.75 |
| | Wake to Sleep | $\Delta\alpha$ | -0.31 | (-0.98, 0.37) | 0.377 | 89 | 1.5 | -0.975 | 0.525 |
| | | $\Delta\beta$ | 0.45 | (-0.07, 0.96) | 0.095 | 89 | 1.5 | -1.25 | 2 |
| | | $\Delta\delta$ | -0.22 | (-0.6, 0.17) | 0.273 | 89 | 0.2 | -1.5 | 0.9 |
| | | $\Delta\text{HR}$ | -0.11 | (-0.25, 0.03) | 0.145 | 64 | 0 | 0 | 2 |
| | | $\Delta\theta$ | -0.20 | (-0.68, 0.27) | 0.409 | 89 | 0.05 | -1.3 | 0.5 |
| Replication Study | REM to NREM | $\Delta\alpha$ | 0.26 | (-0.46, 0.97) | 0.493 | 8 | 1.5 | -0.1 | 2 |
| | | $\Delta\beta$ | -0.75 | (-2.45, 0.94) | 0.400 | 8 | 0.175 | -1.325 | 0.5 |
| | | $\Delta\delta$ | 2.23 | (-0.7, 5.16) | 0.167 | 8 | 1.15 | -1.475 | 1.8 |
| | | $\Delta\text{HR}$ | -0.75 | (-2.21, 0.71) | 0.342 | 7 | 0.825 | -0.675 | 0.5 |
| | | $\Delta\theta$ | 0.11 | (-0.73, 0.95) | 0.803 | 8 | 1.5 | -0.675 | 1.9 |
| | REM to NREM | $\Delta\alpha$ | -2.79 | (-5.55, -0.04) | 0.073 | 12 | 1.5 | -1.5 | 0.5 |
| | | $\Delta\beta$ | 0.05 | (-0.4, 0.51) | 0.825 | 12 | 0.4 | -0.625 | 2 |
| | | $\Delta\delta$ | -2.05 | (-4.21, 0.12) | 0.092 | 12 | 1.05 | -1.5 | 0.5 |
| | | $\Delta\text{HR}$ | -0.73 | (-6.16, 4.7) | 0.782 | 3 | 0.9 | -0.975 | 0.5 |
| | | $\Delta\theta$ | -2.04 | (-2.97, -1.11) | **0.001** | 12 | 0.05 | -1.5 | 0.5 |
| | Wake to Sleep | $\Delta\alpha$ | 0.23 | (-0.5, 0.96) | 0.557 | 17 | 1.5 | -0.225 | 2 |
| | | $\Delta\beta$ | 1.65 | (0.54, 2.76) | **0.011** | 17 | 0.1 | -1.5 | 0.5 |
| | | $\Delta\delta$ | 2.55 | (-3.56, 8.66) | 0.436 | 17 | 1.425 | -1.5 | 0.5 |
| | | $\Delta\text{HR}$ | 0.36 | (-0.2, 0.91) | 0.236 | 17 | 1.5 | -0.025 | 2 |
| | | $\Delta\theta$ | 0.03 | (-0.66, 0.73) | 0.931 | 17 | 1.5 | 0 | 2 |

Rapid eye movement, REM; non-REM, NREM; Confidence Interval, CI; Degrees of Freedom, DF; EEG band power predictor units in percent of total band power; HR predictor units beats-per-minute. **Bolded P values reflect significant associations at $P < 0.05$ .** Predictor and parenchymal resistance values in regression models were scaled to zero mean and unit standard deviation with coefficient estimates representing effect size in standard deviations. Positive and negative thresholds are in units of predictor standard deviation.

**Supplemental Table 6.**
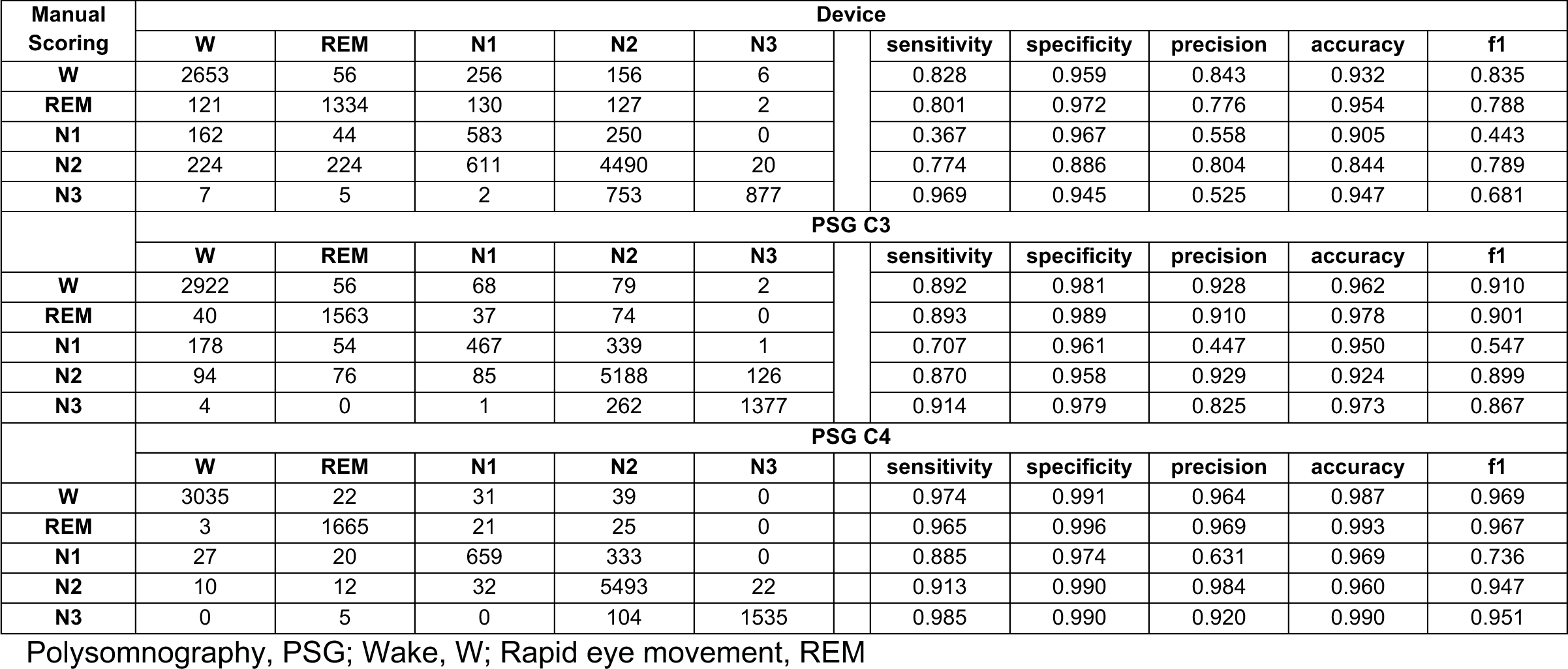
Comparison of investigational device and polysomnography hypnogram staging to manual scoring.

**Supplemental Table 7:**
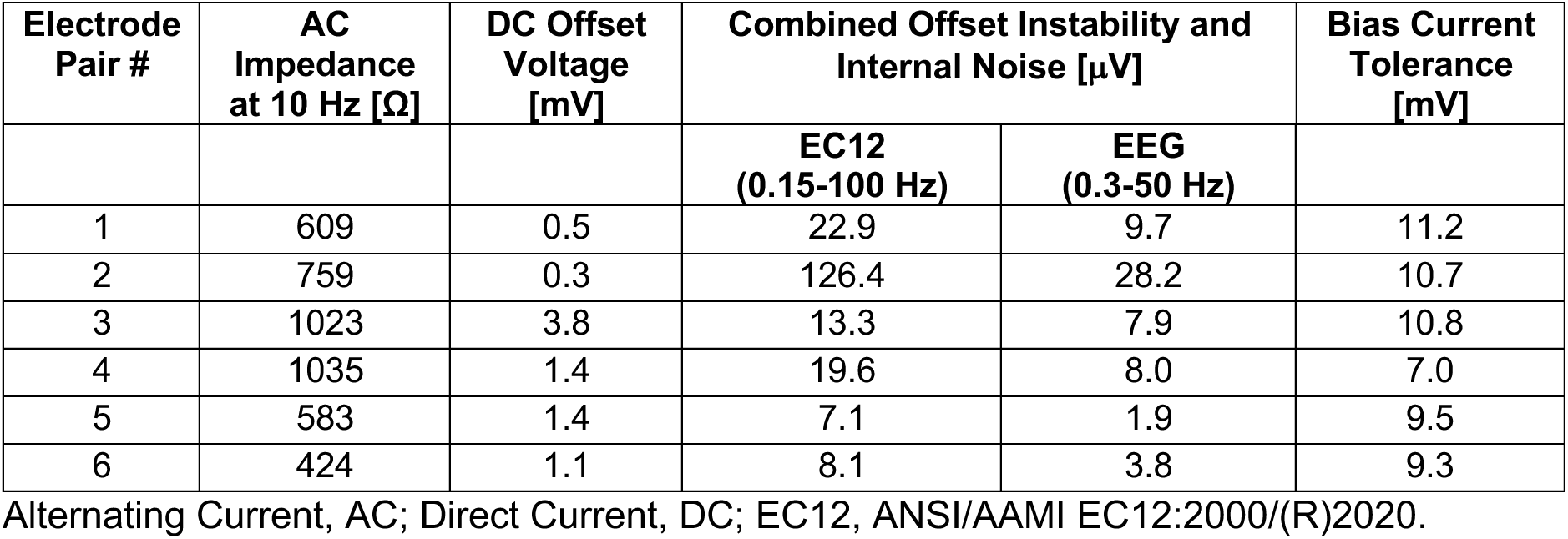
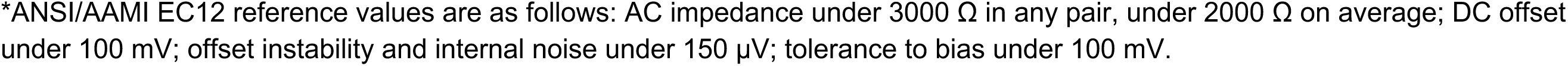
Electrode characterization according to ANSI/AAMI EC12*.

| Electrode Pair # | AC Impedance at 10 Hz [$\Omega$] | DC Offset Voltage [mV] | Combined Offset Instability and Internal Noise [μV] | | Bias Current Tolerance [mV] |
| --- | --- | --- | --- | --- | --- |
|  |  |  | EC12 (0.15-100 Hz) | EEG (0.3-50 Hz) |  |
| 1 | 609 | 0.5 | 22.9 | 9.7 | 11.2 |
| 2 | 759 | 0.3 | 126.4 | 28.2 | 10.7 |
| 3 | 1023 | 3.8 | 13.3 | 7.9 | 10.8 |
| 4 | 1035 | 1.4 | 19.6 | 8.0 | 7.0 |
| 5 | 583 | 1.4 | 7.1 | 1.9 | 9.5 |
| 6 | 424 | 1.1 | 8.1 | 3.8 | 9.3 |

Alternating Current, AC; Direct Current, DC; EC12, ANSI/AAMI EC12:2000/(R)2020.
\*ANSI/AAMI EC12 reference values are as follows: AC impedance under 3000 $\Omega$ in any pair, under 2000 $\Omega$ on average; DC offset under 100 mV; offset instability and internal noise under 150 μV; tolerance to bias under 100 mV.

**Supplemental Table 8.** Benchtop tests for assessing calibration and sensitivity to electrode impedance.

|  |  | Electrode Configuration |  |  |  | Load | Measured Load Components |
| --- | --- | --- | --- | --- | --- | --- | --- |
|  |  | Left |  | Right |  | [Ω] | Measured [Ω] |
| Setting |  | MASTOID | IN-EAR | IN-EAR | MASTOID |  |  |
|  | Cal R 20 | Low | Low | Low | Low | 20 | 20.0 |
|  | Cal R 40 | Low | Low | Low | Low | 40.2 | 40.2 |
|  | Cal R 60 | Low | Low | Low | Low | 60.4 | 60.3 |
|  | Cal R 80 | Low | Low | Low | Low | 80.6 | 80.5 |
|  | Cal R 100 | Low | Low | Low | Low | 100 | 100.4 |
| Calibration 2R1C | Cal 2R1C 20 | Low | Low | Low | Low | 20\|(470 + 22n) | Rs = 20, Rp=468.2, Cp = 23.3n |
|  | Cal 2R1C 40 | Low | Low | Low | Low | 40.2\|(470 + 22n) | Rs = 40.2, Rp=468.2, Cp = 23.3n |
|  | Cal 2R1C 60 | Low | Low | Low | Low | 60.4\|(470 + 22n) | Rs = 60.3, Rp=468.2, Cp = 23.3n |
|  | Cal 2R1C 80 | Low | Low | Low | Low | 80.6\|(470 + 22n) | Rs = 80.5, Rp=468.2, Cp = 23.3n |
|  | Cal 2R1C 100 | Low | Low | Low | Low | 100\|(470 + 22n) | Rs = 100.4, Rp=468.2, Cp = 23.3n |
| Sensitivity to Electrode Impedance | Low All | Low | Low | Low | Low | 50\|(470 + 22n) | Rs = 49.9, Rp=468.2, Cp = 23.3n |
|  | High Right Mastoid | Low | Low | Low | High | 50\|(470 + 22n) | Rs = 49.9, Rp=468.2, Cp = 23.3n |
|  | High Right | Low | Low | High | High | 50\|(470 + 22n) | Rs = 49.9, Rp=468.2, Cp = 23.3n |
|  | High Right In-Ear | Low | Low | High | Low | 50\|(470 + 22n) | Rs = 49.9, Rp=468.2, Cp = 23.3n |
|  | High In-Ear | Low | High | High | Low | 50\|(470 + 22n) | Rs = 49.9, Rp=468.2, Cp = 23.3n |
|  | High Left In-Ear | Low | High | Low | Low | 50\|(470 + 22n) | Rs = 49.9, Rp=468.2, Cp = 23.3n |
|  | High Left | High | High | Low | Low | 50\|(470 + 22n) | Rs = 49.9, Rp=468.2, Cp = 23.3n |
|  | High Left Mastoid | High | Low | Low | Low | 50\|(470 + 22n) | Rs = 49.9, Rp=468.2, Cp = 23.3n |
|  | High Mastoids | High | Low | Low | High | 50\|(470 + 22n) | Rs = 49.9, Rp=468.2, Cp = 23.3n |
|  | High All | High | High | High | High | 50\|(470 + 22n) | Rs = 49.9, Rp=468.2, Cp = 23.3n |

Element to change from one benchtop test to the next

**Supplemental Table 9.** Absolute error of resistance for benchtop tests (values in ohms)

|  | Setting | 1kHz | 1.6kHz | 2.28kHz | 3.2kHz | 4.55kHz | 6.4kHz | 8.53kHz | 12.8kHz | 16kHz | 25.6kHz | 32kHz | 51.2kHz | 64kHz | 85.3kHz | 102kHz | 128kHz |
| --- | --- | --- | --- | --- | --- | --- | --- | --- | --- | --- | --- | --- | --- | --- | --- | --- | --- |
| Calibration R | Cal R 20 | 0.125 | 0.151 | 0.157 | 0.119 | 0.127 | 0.154 | 0.182 | 0.190 | 0.220 | 0.308 | 0.320 | 0.274 | 0.094 | 0.379 | 0.700 | 0.702 |
|  | Cal R 40 | 0.220 | 0.161 | 0.152 | 0.192 | 0.221 | 0.094 | 0.164 | 0.117 | 0.127 | 0.151 | 0.200 | 0.301 | 0.134 | 0.438 | 0.344 | 0.396 |
|  | Cal R 60 | 0.333 | 0.231 | 0.325 | 0.307 | 0.301 | 0.413 | 0.411 | 0.208 | 0.308 | 0.326 | 0.276 | 0.456 | 0.327 | 0.412 | 0.268 | 0.301 |
|  | Cal R 80 | 0.104 | 0.175 | 0.242 | 0.205 | 0.314 | 0.140 | 0.154 | 0.257 | 0.258 | 0.332 | 0.333 | 0.257 | 0.284 | 0.298 | 0.317 | 0.230 |
|  | Cal R 100 | 0.301 | 0.288 | 0.268 | 0.251 | 0.261 | 0.323 | 0.236 | 0.271 | 0.458 | 0.380 | 0.199 | 0.157 | 0.387 | 0.324 | 0.196 | 0.134 |
| Calibration 2R1C | Cal 2R1C 20 | 0.232 | 0.277 | 0.244 | 0.105 | 0.130 | 0.125 | 0.141 | 0.087 | 0.211 | 0.247 | 0.383 | 0.355 | 0.228 | 0.371 | 0.522 | 0.756 |
|  | Cal 2R1C 40 | 0.191 | 0.117 | 0.112 | 0.305 | 0.207 | 0.042 | 0.207 | 0.149 | 0.268 | 0.157 | 0.134 | 0.240 | 0.314 | 0.242 | 0.421 | 0.311 |
|  | Cal 2R1C 60 | 0.372 | 0.213 | 0.356 | 0.292 | 0.202 | 0.209 | 0.263 | 0.276 | 0.333 | 0.475 | 0.260 | 0.301 | 0.522 | 0.434 | 0.380 | 0.318 |
|  | Cal 2R1C 80 | 0.215 | 0.166 | 0.249 | 0.175 | 0.186 | 0.252 | 0.231 | 0.168 | 0.348 | 0.384 | 0.303 | 0.211 | 0.333 | 0.331 | 0.280 | 0.143 |
|  | Cal 2R1C 100 | 0.384 | 0.304 | 0.256 | 0.295 | 0.195 | 0.159 | 0.294 | 0.283 | 0.428 | 0.423 | 0.339 | 0.379 | 0.208 | 0.172 | 0.193 | 0.021 |
| Sensitivity to Electrode Impedance | Low All | 0.284 | 0.159 | 0.181 | 0.318 | 0.227 | 0.171 | 0.280 | 0.263 | 0.136 | 0.152 | 0.165 | 0.279 | 0.237 | 0.192 | 0.454 | 0.330 |
|  | High Right Mastoid | 0.123 | 0.003 | 0.165 | 0.220 | 0.176 | 0.210 | 0.071 | 0.120 | 0.128 | 0.085 | 0.001 | 0.300 | 0.242 | 0.650 | 0.362 | 0.725 |
|  | High Right | 0.131 | 0.242 | 0.079 | 0.279 | 0.232 | 0.253 | 0.274 | 0.144 | 0.202 | 0.186 | 0.167 | -0.022 | 0.383 | 0.586 | 0.620 | 0.911 |
|  | High Right In-Ear | 0.259 | 0.256 | 0.249 | 0.297 | 0.252 | 0.347 | 0.183 | 0.182 | 0.279 | 0.256 | 0.212 | 0.315 | 0.139 | 0.529 | 0.457 | 0.576 |
|  | High In-Ear | 0.384 | 0.263 | 0.109 | 0.233 | 0.350 | 0.236 | 0.237 | 0.292 | 0.266 | 0.259 | 0.312 | 0.327 | 0.379 | 0.415 | 0.446 | 0.450 |
|  | High Left In-Ear | 0.210 | 0.169 | 0.236 | 0.313 | 0.262 | 0.319 | 0.229 | 0.269 | 0.280 | 0.332 | 0.324 | 0.168 | 0.303 | 0.255 | 0.361 | 0.310 |
|  | High Left | 0.266 | 0.337 | 0.372 | 0.241 | 0.271 | 0.281 | 0.252 | 0.167 | 0.388 | 0.501 | 0.416 | 0.266 | 0.128 | 0.225 | 0.218 | 0.006 |
|  | High Left Mastoid | 0.341 | 0.325 | 0.217 | 0.225 | 0.222 | 0.193 | 0.185 | 0.242 | 0.430 | 0.363 | 0.269 | 0.242 | 0.143 | 0.097 | 0.268 | 0.018 |
|  | High Mastoids | 0.307 | 0.227 | 0.235 | 0.364 | 0.286 | 0.192 | 0.348 | 0.363 | 0.339 | 0.277 | 0.238 | 0.146 | 0.498 | 0.455 | 0.492 | 0.487 |
|  | High All | 0.517 | 0.412 | 0.447 | 0.374 | 0.418 | 0.420 | 0.342 | 0.397 | 0.425 | 0.461 | 0.375 | 0.418 | 0.527 | 0.727 | 0.852 | 0.843 |

**Supplemental Table 10.** Absolute error of reactance for benchtop tests (values in ohms)

|  | Setting | 1kHz | 1.6kHz | 2.28kHz | 3.2kHz | 4.55kHz | 6.4kHz | 8.53kHz | 12.8kHz | 16kHz | 25.6kHz | 32kHz | 51.2kHz | 64kHz | 85.3kHz | 102kHz | 128kHz |
| --- | --- | --- | --- | --- | --- | --- | --- | --- | --- | --- | --- | --- | --- | --- | --- | --- | --- |
| Calibration R | Cal R 20 | 0.087 | -0.031 | 0.055 | -0.046 | 0.010 | -0.016 | 0.104 | -0.030 | 0.142 | 0.237 | 0.172 | 0.188 | 0.018 | 0.250 | 0.197 | 0.288 |
|  | Cal R 40 | 0.032 | 0.042 | 0.011 | 0.055 | -0.054 | -0.057 | 0.126 | 0.035 | 0.018 | -0.027 | 0.027 | 0.087 | -0.070 | 0.057 | 0.102 | 0.040 |
|  | Cal R 60 | 0.103 | 0.007 | -0.025 | 0.070 | -0.052 | -0.017 | 0.025 | 0.044 | 0.069 | -0.071 | 0.090 | -0.041 | -0.032 | -0.129 | -0.050 | -0.107 |
|  | Cal R 80 | 0.036 | 0.073 | 0.015 | -0.132 | -0.042 | 0.064 | 0.012 | -0.010 | 0.047 | 0.072 | 0.040 | -0.010 | -0.061 | -0.092 | -0.162 | -0.181 |
|  | Cal R 100 | -0.066 | -0.054 | 0.028 | -0.019 | 0.011 | 0.153 | 0.057 | -0.051 | -0.002 | 0.022 | 0.045 | -0.003 | -0.133 | -0.024 | -0.050 | -0.121 |
| Calibration 2R1C | Cal 2R1C 20 | 0.044 | -0.039 | -0.037 | 0.019 | 0.043 | 0.033 | 0.031 | 0.051 | 0.091 | 0.143 | 0.210 | 0.211 | 0.237 | 0.285 | 0.413 | 0.380 |
|  | Cal 2R1C 40 | 0.001 | -0.070 | -0.029 | 0.084 | -0.049 | 0.030 | -0.050 | -0.033 | 0.026 | 0.000 | -0.008 | 0.020 | 0.117 | 0.074 | 0.097 | 0.114 |
|  | Cal 2R1C 60 | 0.019 | 0.006 | -0.043 | -0.017 | 0.009 | 0.032 | 0.120 | -0.120 | -0.005 | 0.034 | -0.149 | 0.089 | 0.106 | 0.041 | 0.204 | 0.122 |
|  | Cal 2R1C 80 | 0.095 | -0.005 | 0.018 | 0.145 | -0.018 | -0.073 | 0.001 | 0.017 | 0.013 | 0.010 | 0.145 | 0.058 | 0.076 | -0.012 | -0.028 | 0.104 |
|  | Cal 2R1C 100 | -0.030 | -0.118 | -0.014 | 0.065 | -0.066 | 0.025 | 0.070 | 0.055 | 0.074 | 0.037 | 0.174 | 0.158 | 0.012 | 0.092 | 0.041 | 0.105 |
| Sensitivity to Electrode Impedance | Low All | 0.015 | -0.066 | 0.047 | -0.052 | 0.046 | 0.016 | 0.068 | -0.040 | -0.032 | -0.112 | 0.006 | 0.107 | 0.121 | 0.006 | 0.093 | 0.112 |
|  | High Right Mastoid | 0.113 | 0.084 | 0.030 | 0.095 | 0.131 | -0.034 | 0.043 | 0.043 | 0.103 | 0.139 | 0.160 | 0.251 | 0.309 | 0.388 | 0.389 | 0.306 |
|  | High Right | 0.029 | 0.059 | -0.024 | 0.159 | 0.000 | 0.098 | 0.028 | 0.016 | 0.059 | 0.124 | 0.186 | 0.136 | 0.243 | 0.204 | 0.459 | 0.335 |
|  | High Right In-Ear | 0.015 | -0.074 | -0.001 | -0.019 | 0.073 | 0.044 | 0.076 | 0.042 | 0.011 | 0.073 | 0.047 | 0.131 | 0.123 | -0.009 | 0.150 | 0.133 |
|  | High In-Ear | 0.071 | -0.015 | 0.000 | -0.074 | 0.026 | -0.104 | -0.007 | 0.028 | 0.087 | 0.045 | 0.021 | 0.103 | 0.203 | 0.111 | 0.043 | 0.160 |
|  | High Left In-Ear | -0.007 | 0.001 | 0.015 | 0.013 | -0.042 | -0.168 | 0.045 | 0.010 | -0.049 | 0.006 | -0.032 | 0.038 | 0.237 | 0.150 | 0.070 | 0.137 |
|  | High Left | -0.150 | -0.117 | -0.154 | -0.178 | 0.012 | -0.104 | -0.025 | -0.124 | -0.172 | -0.159 | -0.127 | -0.253 | -0.173 | -0.147 | -0.124 | -0.099 |
|  | High Left Mastoid | -0.173 | -0.093 | -0.169 | -0.045 | -0.104 | -0.073 | 0.001 | -0.104 | -0.032 | -0.211 | -0.271 | -0.187 | -0.212 | -0.076 | -0.251 | -0.091 |
|  | High Mastoids | 0.034 | -0.002 | 0.036 | -0.023 | -0.100 | -0.010 | -0.054 | 0.032 | -0.042 | -0.103 | 0.008 | -0.006 | 0.083 | 0.082 | 0.176 | 0.058 |
|  | High All | -0.041 | -0.114 | -0.083 | -0.109 | -0.091 | -0.066 | -0.051 | 0.065 | 0.083 | 0.103 | 0.013 | 0.138 | 0.278 | 0.296 | 0.389 | 0.401 |

**Supplemental Table 11.**
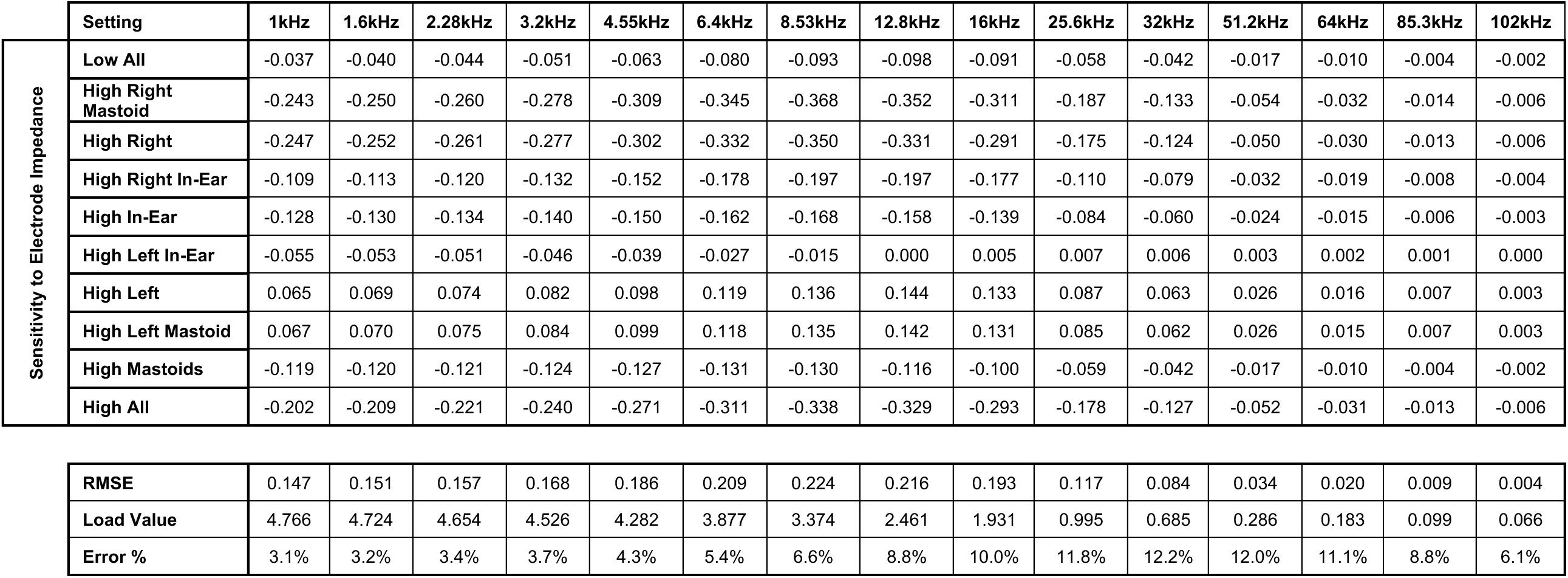
Debye model absolute prediction error (ohms) of dispersion relative to 128kHz.

|  | Setting | 1kHz | 1.6kHz | 2.28kHz | 3.2kHz | 4.55kHz | 6.4kHz | 8.53kHz | 12.8kHz | 16kHz | 25.6kHz | 32kHz | 51.2kHz | 64kHz | 85.3kHz | 102kHz |
| --- | --- | --- | --- | --- | --- | --- | --- | --- | --- | --- | --- | --- | --- | --- | --- | --- |
| Sensitivity to Electrode Impedance | Low All | -0.037 | -0.040 | -0.044 | -0.051 | -0.063 | -0.080 | -0.093 | -0.098 | -0.091 | -0.058 | -0.042 | -0.017 | -0.010 | -0.004 | -0.002 |
|  | High Right Mastoid | -0.243 | -0.250 | -0.260 | -0.278 | -0.309 | -0.345 | -0.368 | -0.352 | -0.311 | -0.187 | -0.133 | -0.054 | -0.032 | -0.014 | -0.006 |
|  | High Right | -0.247 | -0.252 | -0.261 | -0.277 | -0.302 | -0.332 | -0.350 | -0.331 | -0.291 | -0.175 | -0.124 | -0.050 | -0.030 | -0.013 | -0.006 |
|  | High Right In-Ear | -0.109 | -0.113 | -0.120 | -0.132 | -0.152 | -0.178 | -0.197 | -0.197 | -0.177 | -0.110 | -0.079 | -0.032 | -0.019 | -0.008 | -0.004 |
|  | High In-Ear | -0.128 | -0.130 | -0.134 | -0.140 | -0.150 | -0.162 | -0.168 | -0.158 | -0.139 | -0.084 | -0.060 | -0.024 | -0.015 | -0.006 | -0.003 |
|  | High Left In-Ear | -0.055 | -0.053 | -0.051 | -0.046 | -0.039 | -0.027 | -0.015 | 0.000 | 0.005 | 0.007 | 0.006 | 0.003 | 0.002 | 0.001 | 0.000 |
|  | High Left | 0.065 | 0.069 | 0.074 | 0.082 | 0.098 | 0.119 | 0.136 | 0.144 | 0.133 | 0.087 | 0.063 | 0.026 | 0.016 | 0.007 | 0.003 |
|  | High Left Mastoid | 0.067 | 0.070 | 0.075 | 0.084 | 0.099 | 0.118 | 0.135 | 0.142 | 0.131 | 0.085 | 0.062 | 0.026 | 0.015 | 0.007 | 0.003 |
|  | High Mastoids | -0.119 | -0.120 | -0.121 | -0.124 | -0.127 | -0.131 | -0.130 | -0.116 | -0.100 | -0.059 | -0.042 | -0.017 | -0.010 | -0.004 | -0.002 |
|  | High All | -0.202 | -0.209 | -0.221 | -0.240 | -0.271 | -0.311 | -0.338 | -0.329 | -0.293 | -0.178 | -0.127 | -0.052 | -0.031 | -0.013 | -0.006 |

|  |  |  |  |  |  |  |  |  |  |  |  |  |  |  |  |
| --- | --- | --- | --- | --- | --- | --- | --- | --- | --- | --- | --- | --- | --- | --- | --- |
| RMSE | 0.147 | 0.151 | 0.157 | 0.168 | 0.186 | 0.209 | 0.224 | 0.216 | 0.193 | 0.117 | 0.084 | 0.034 | 0.020 | 0.009 | 0.004 |
| Load Value | 4.766 | 4.724 | 4.654 | 4.526 | 4.282 | 3.877 | 3.374 | 2.461 | 1.931 | 0.995 | 0.685 | 0.286 | 0.183 | 0.099 | 0.066 |
| Error % | 3.1% | 3.2% | 3.4% | 3.7% | 4.3% | 5.4% | 6.6% | 8.8% | 10.0% | 11.8% | 12.2% | 12.0% | 11.1% | 8.8% | 6.1% |

**Supplemental Table 12:** Minimum significant change in mean R_p_ and correlation with a predictor using the R_p_ standard deviation of the electrode impedance sensitivity tests.

| Number of Samples | 25 | 100 |
| --- | --- | --- |
| Standard Deviation [$\Omega$] | 0.113 | 0.113 |
| Standard Error [$\Omega$] | 0.023 | 0.011 |
| Minimum Significant Change [$\Omega$] | 0.044 | 0.022 |
| Minimum Significant Correlation | 0.39 | 0.20 |

Brain parenchymal resistance, $R_p$ .

**Supplemental Figure 1.**
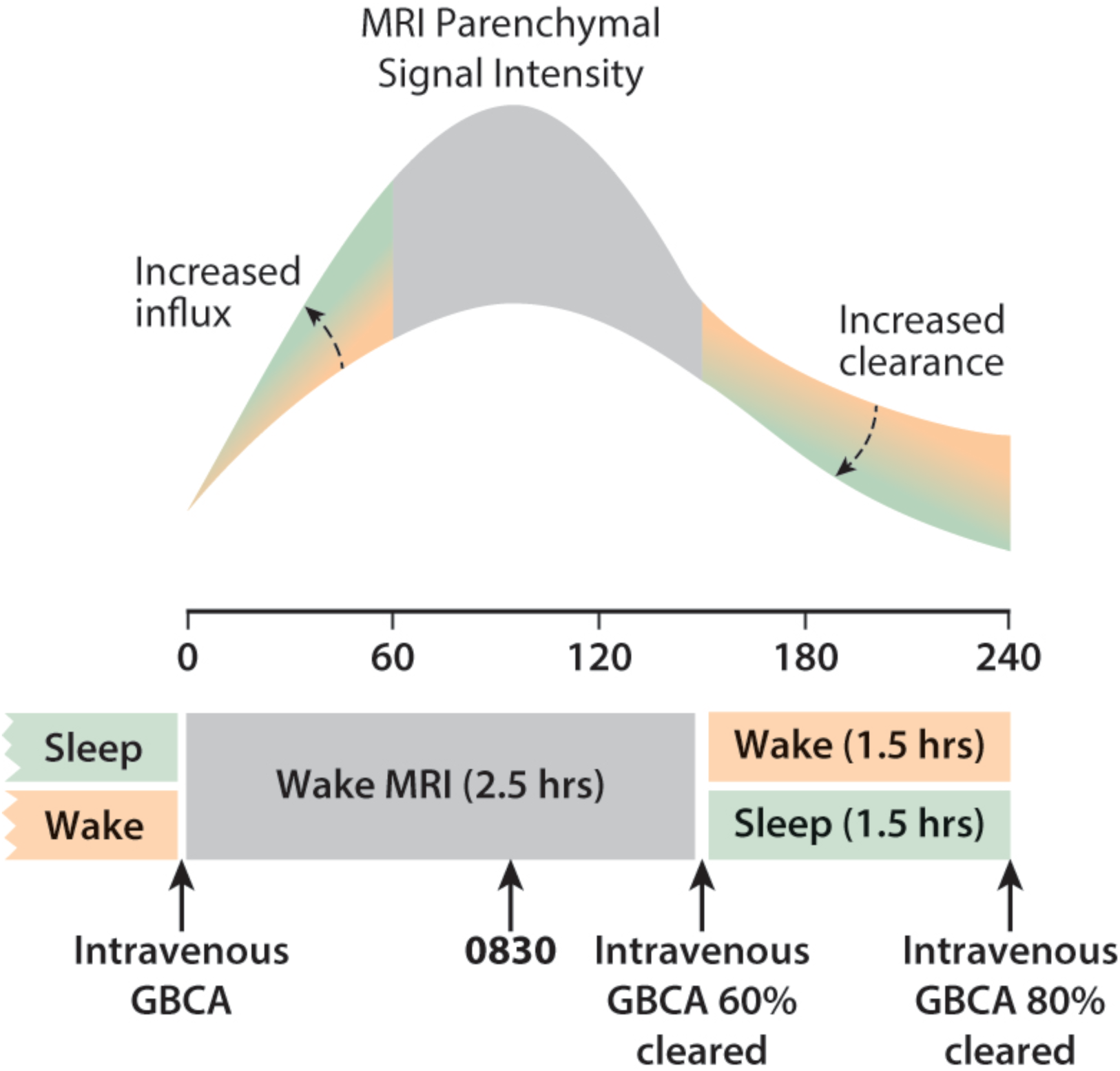
An illustration of the effect of increased CSF influx into the brain parenchyma and clearance from the parenchyma on contrast-enhanced MRI signal intensity in the Benchmarking study. An increase in CSF influx into the brain parenchyma following intravenous GBCA leads to greater contrast delivery to the brain parenchyma and a corresponding increase in MRI signal intensity. MRI signal intensity of the brain parenchyma has been shown to peak at approximately 90 minutes post-injection^15^ which occurred at time 0830 in the Benchmarking study. With a terminal half-life of 109 minutes^28^, intravenous contrast is 60% cleared at 140 minutes post-injection when the morning sleep/wake intervention begins and is 80% cleared at the end of the morning intervention when the second post-contrast MRI scan is acquired. During this morning intervention, an increase in CSF clearance from the brain parenchyma leads to a corresponding decrease in MRI signal intensity.

**Supplemental Figure 2.**
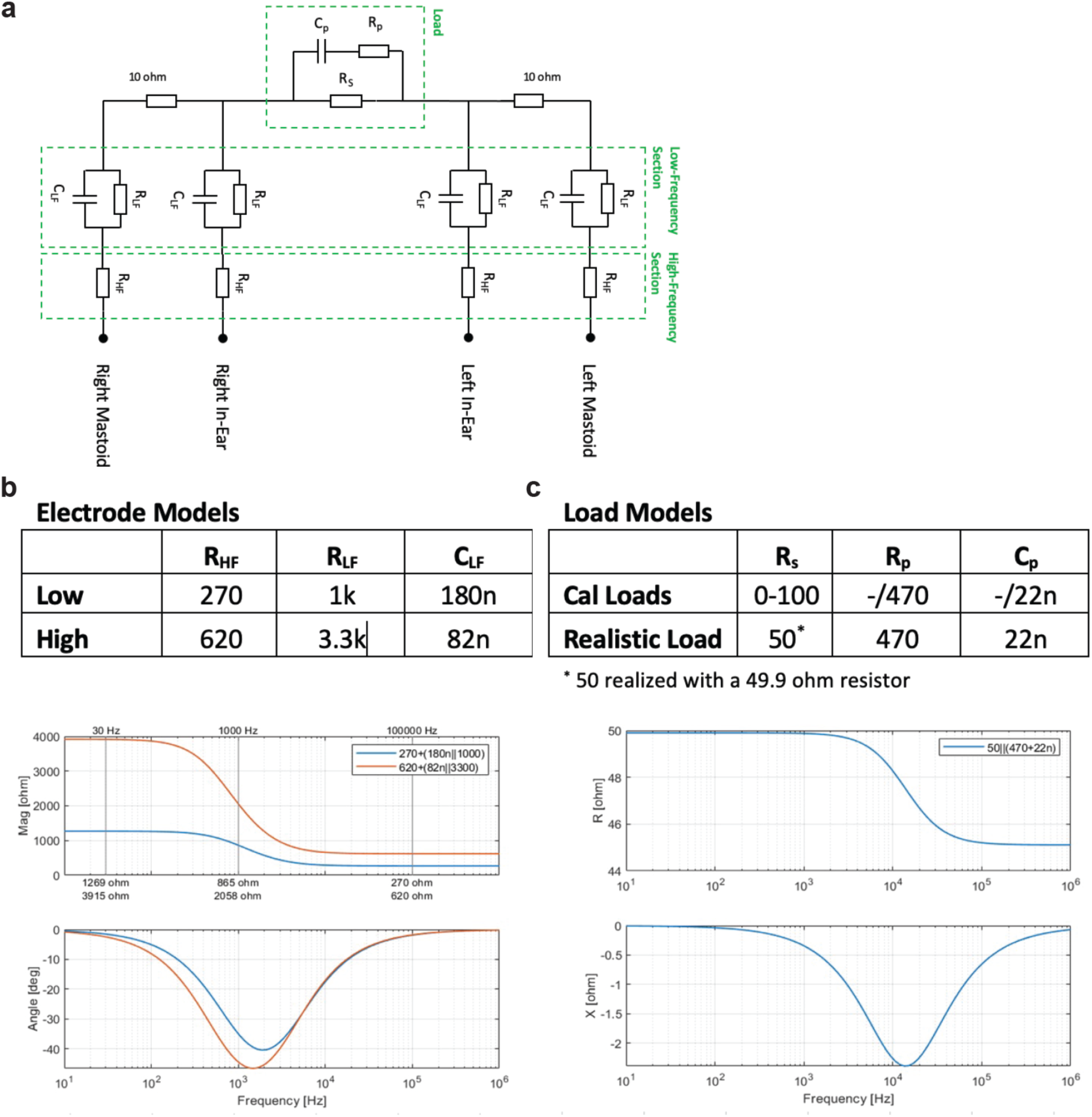
**a**, Circuit model of transcranial measurements using four-point electrical impedance spectroscopy and models for each of the four electrode contact impedances. **b**, Electrode impedance models comprised a low impedance and a high impedance model, representing the range of in-human impedances. **c**, Calibration loads varied the resistance **R_s_** of the free fluid or interstitial fluid compartment from 20 to 100 Ω in increments of 20 Ω, with and additional R_p_C_p_ in parallel to simulate complex loads. The realistic load parameters were chosen to create a frequency dispersion that approximated the in-human measured dispersions. **d**, Electrode impedance dispersion over the measured frequencies for the low and high impedance model. **e**, Load impedance dispersion over the measured frequencies using the Real Load model.

**Supplemental Figure 3.**
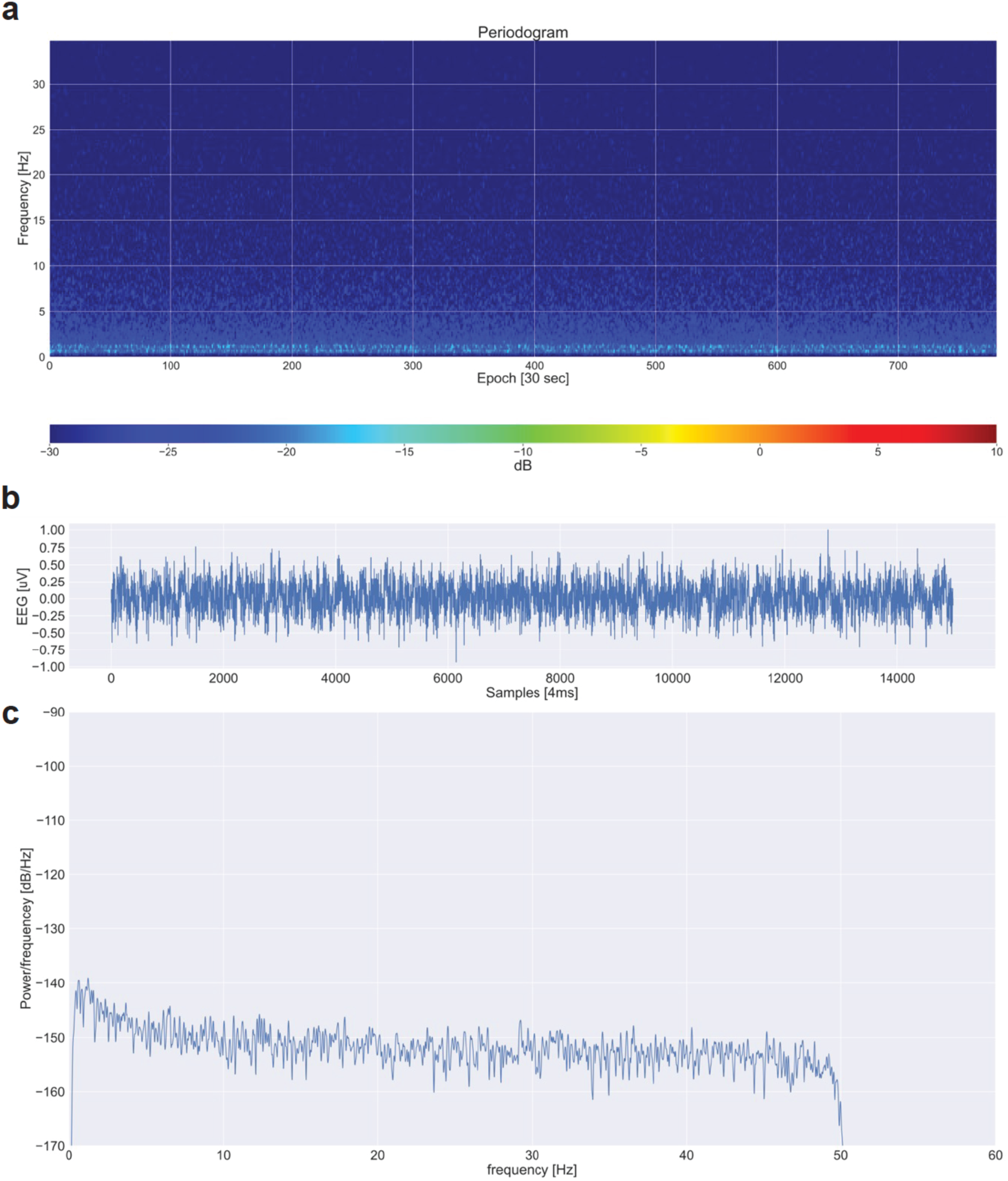
**a**, Periodogram of a device over a 12 hour test using four Ag/AgCl sintered disk electrodes on an Agar phantom with EEG gel. For reference, human EEG beta power during NREM sleep is above −10dB on the device. **b**, EEG recording over an epoch shows 0.5 µV peak-to-peak noise. **c**, Power spectral density shows a noise floor at −150dB/Hz with a 1/f increase to −140db/Hz below 10Hz.

## REFERENCES

1 Iliff, J. J. et al. A paravascular pathway facilitates CSF flow through the brain parenchyma and the clearance of interstitial solutes, including amyloid β. Sci Transl Med 4, 147ra111 (2012). 10.1126/scitranslmed.3003748

2 Xie, L. et al. Sleep drives metabolite clearance from the adult brain. Science 342, 373–377 (2013). 10.1126/science.1241224

3 Iliff, J. J. et al. Impairment of glymphatic pathway function promotes tau pathology after traumatic brain injury. J Neurosci 34, 16180–16193 (2014). 10.1523/JNEUROSCI.3020-14.2014

4 Harrison, I. F. et al. Impaired glymphatic function and clearance of tau in an Alzheimer’s disease model. Brain 143, 2576–2593 (2020). 10.1093/brain/awaa179

5 Ishida, K. et al. Glymphatic system clears extracellular tau and protects from tau aggregation and neurodegeneration. J Exp Med 219 (2022). 10.1084/jem.20211275

6 Cui, H. et al. Decreased AQP4 Expression Aggravates ɑ-Synuclein Pathology in Parkinson’s Disease Mice, Possibly via Impaired Glymphatic Clearance. J Mol Neurosci 71, 2500–2513 (2021). 10.1007/s12031-021-01836-4

7 Zou, W. et al. Blocking meningeal lymphatic drainage aggravates Parkinson’s disease-like pathology in mice overexpressing mutated α-synuclein. Transl Neurodegener 8, 7 (2019). 10.1186/s40035-019-0147-y

8 Hablitz, L. M. & Nedergaard, M. The Glymphatic System: A Novel Component of Fundamental Neurobiology. J Neurosci 41, 7698–7711 (2021). 10.1523/JNEUROSCI.0619-21.2021

9 Klostranec, J. M. et al. Current Concepts in Intracranial Interstitial Fluid Transport and the Glymphatic System: Part II-Imaging Techniques and Clinical Applications. Radiology 301, 516–532 (2021). 10.1148/radiol.2021204088

10 Hablitz, L. M. et al. Circadian control of brain glymphatic and lymphatic fluid flow. Nat Commun 11, 4411 (2020). 10.1038/s41467-020-18115-2

11 Hablitz, L. M. et al. Increased glymphatic influx is correlated with high EEG delta power and low heart rate in mice under anesthesia. Sci Adv 5, eaav5447 (2019). 10.1126/sciadv.aav5447

12 Eide, P. K., Vinje, V., Pripp, A. H., Mardal, K. A. & Ringstad, G. Sleep deprivation impairs molecular clearance from the human brain. Brain 144, 863–874 (2021). 10.1093/brain/awaa443

13 Ringstad, G. et al. Brain-wide glymphatic enhancement and clearance in humans assessed with MRI. JCI Insight 3 (2018). 10.1172/jci.insight.121537

14 Deike-Hofmann, K. et al. Glymphatic Pathway of Gadolinium-Based Contrast Agents Through the Brain: Overlooked and Misinterpreted. Invest Radiol 54, 229–237 (2019). 10.1097/RLI.0000000000000533

15 Richmond, S. B. et al. Quantification approaches for magnetic resonance imaging following intravenous gadolinium injection: A window into brain-wide glymphatic function. Eur J Neurosci 57, 1689–1704 (2023). 10.1111/ejn.15974

16 Bera, T. K. Bioelectrical Impedance Methods for Noninvasive Health Monitoring: A Review. J Med Eng 2014, 381251 (2014). 10.1155/2014/381251

17 Abasi, S., Aggas, J. R., Garayar-Leyva, G. G., Walther, B. K. & Guiseppi-Elie, A. Bioelectrical Impedance Spectroscopy for Monitoring Mammalian Cells and Tissues under Different Frequency Domains: A Review. ACS Meas Sci Au 2, 495–516 (2022). 10.1021/acsmeasuresciau.2c00033

18 Stupin, D. D. et al. Bioimpedance Spectroscopy: Basics and Applications. ACS Biomater Sci Eng 7, 1962–1986 (2021). 10.1021/acsbiomaterials.0c01570

19 Gabriel, S., Lau, R. W. & Gabriel, C. The dielectric properties of biological tissues: II. Measurements in the frequency range 10 Hz to 20 GHz. Phys Med Biol 41, 2251–2269 (1996). 10.1088/0031-9155/41/11/002

20 Gabriel, C. Dielectric properties of biological tissue: variation with age. Bioelectromagnetics Suppl 7, S12–18 (2005). 10.1002/bem.20147

21 Jessen, N. A., Munk, A. S., Lundgaard, I. & Nedergaard, M. The Glymphatic System: A Beginner’s Guide. Neurochem Res 40, 2583–2599 (2015). 10.1007/s11064-015-1581-6

22 Nakamura, K. et al. Diurnal fluctuations in brain volume: Statistical analyses of MRI from large populations. Neuroimage 118, 126–132 (2015). 10.1016/j.neuroimage.2015.05.077

23 Elvsåshagen, T. et al. Cerebral blood flow changes after a day of wake, sleep, and sleep deprivation. Neuroimage 186, 497–509 (2019). 10.1016/j.neuroimage.2018.11.032

24. Dagum, P. Non-invasive assessment of glymphatic flow and neurodegeneration from a wearable device. (2022).

25. Dagum, P. Non-invasive assessment of glymphatic flow and neurodegeneration from a wearable device. (2022).

26. Nasreddine, Z. S., et al. The Montreal Cognitive Assessment, MoCA: a brief screening tool for mild cognitive impairment. J Am Geriatr Soc 53, 695-699 (2005). 10.1111/j.1532-5415.2005.53221.x

27 Brown, L. M. & Schinka, J. A. Development and initial validation of a 15-item informant version of the Geriatric Depression Scale. Int J Geriatr Psychiatry 20, 911–918 (2005). 10.1002/gps.1375

28 Administration., U. F. a. D. Gadavist (gadobutrol) injection, for intravenous use, <https://www.accessdata.fda.gov/drugsatfda_docs/label/2011/201277s000lbl.pdf> (2011).

29 Heinze, G., Wallisch, C. & Dunkler, D. Variable selection - A review and recommendations for the practicing statistician. Biom J 60, 431–449 (2018). 10.1002/bimj.201700067

30 Iliff, J. J. et al. Brain-wide pathway for waste clearance captured by contrast-enhanced MRI. J Clin Invest 123, 1299–1309 (2013). 10.1172/JCI67677

31 Boukamp, B. A. A Linear Kronig-Kramers Transform Test for Immittance Data Validation. Journal of The Electrochemical Society 142, 1885 (1995).

32 Schonleber, M., Klotz, D. & Ivers-Tiffee, E. A Method for Improving the Robustness of linear Kramers-Kronig Validity Tests. Electrochimica Acta 131, 20–27 (2014).

33 Song, J. et al. Electrical Impedance Changes at Different Phases of Cerebral Edema in Rats with Ischemic Brain Injury. Biomed Res Int 2018, 9765174 (2018). 10.1155/2018/9765174

34 Abboud, T., Mielke, D. & Rohde, V. Mini Review: Impedance Measurement in Neuroscience and Its Prospective Application in the Field of Surgical Neurooncology. Front Neurol 12, 825012 (2021). 10.3389/fneur.2021.825012

35 Witkowska-Wrobel, A., Aristovich, K., Crawford, A., Perkins, J. D. & Holder, D. Imaging of focal seizures with Electrical Impedance Tomography and depth electrodes in real time. Neuroimage 234, 117972 (2021). 10.1016/j.neuroimage.2021.117972

36 Romsauerova, A. et al. Multi-frequency electrical impedance tomography (EIT) of the adult human head: initial findings in brain tumours, arteriovenous malformations and chronic stroke, development of an analysis method and calibration. Physiol Meas 27, S147–161 (2006). 10.1088/0967-3334/27/5/S13

37 Iliff, J. J. et al. Cerebral arterial pulsation drives paravascular CSF-interstitial fluid exchange in the murine brain. J Neurosci 33, 18190–18199 (2013). 10.1523/JNEUROSCI.1592-13.2013

38 Ringstad, G., Vatnehol, S. A. S. & Eide, P. K. Glymphatic MRI in idiopathic normal pressure hydrocephalus. Brain 140, 2691–2705 (2017). 10.1093/brain/awx191

39 Mestre, H. et al. Aquaporin-4-dependent glymphatic solute transport in the rodent brain. Elife 7 (2018). 10.7554/eLife.40070

40 Mestre, H. et al. Flow of cerebrospinal fluid is driven by arterial pulsations and is reduced in hypertension. Nat Commun 9, 4878 (2018). 10.1038/s41467-018-07318-3

41 Jiang-Xie, L. F. et al. Neuronal dynamics direct cerebrospinal fluid perfusion and brain clearance. Nature 627, 157–164 (2024). 10.1038/s41586-024-07108-6

42 Piantino, J., Lim, M. M., Newgard, C. D. & Iliff, J. Linking Traumatic Brain Injury, Sleep Disruption and Post-Traumatic Headache: a Potential Role for Glymphatic Pathway Dysfunction. Curr Pain Headache Rep 23, 62 (2019). 10.1007/s11916-019-0799-4

43 Nedergaard, M. & Goldman, S. A. Glymphatic failure as a final common pathway to dementia. Science 370, 50–56 (2020). 10.1126/science.abb8739

44 Goldman, N., Hablitz, L. M., Mori, Y. & Nedergaard, M. The Glymphatic System and Pain. Med Acupunct 32, 373–376 (2020). 10.1089/acu.2020.1489

45 Braun, M. & Iliff, J. J. The impact of neurovascular, blood-brain barrier, and glymphatic dysfunction in neurodegenerative and metabolic diseases. Int Rev Neurobiol 154, 413–436 (2020). 10.1016/bs.irn.2020.02.006

46 Gakuba, C. et al. General Anesthesia Inhibits the Activity of the “Glymphatic System”. Theranostics 8, 710–722 (2018). 10.7150/thno.19154

47 Miao, A. et al. Brain clearance is reduced during sleep and anesthesia. Nat Neurosci 27, 1046–1050 (2024). 10.1038/s41593-024-01638-y

48 Kress, B. T. et al. Impairment of paravascular clearance pathways in the aging brain. Ann Neurol 76, 845–861 (2014). 10.1002/ana.24271

49 Wang, M. et al. Focal Solute Trapping and Global Glymphatic Pathway Impairment in a Murine Model of Multiple Microinfarcts. J Neurosci 37, 2870–2877 (2017). 10.1523/JNEUROSCI.2112-16.2017

50 Li, M. et al. Impaired Glymphatic Function and Pulsation Alterations in a Mouse Model of Vascular Cognitive Impairment. Front Aging Neurosci 13, 788519 (2021). 10.3389/fnagi.2021.788519

51 Xu, Z. et al. Deletion of aquaporin-4 in APP/PS1 mice exacerbates brain Aβ accumulation and memory deficits. Mol Neurodegener 10, 58 (2015). 10.1186/s13024-015-0056-1

52 Simon, M. et al. Loss of perivascular aquaporin-4 localization impairs glymphatic exchange and promotes amyloid β plaque formation in mice. Alzheimers Res Ther 14, 59 (2022). 10.1186/s13195-022-00999-5

53 Pedersen, T. J., Keil, S. A., Han, W., Wang, M. X. & Iliff, J. J. The effect of aquaporin-4 mis-localization on Aβ deposition in mice. Neurobiol Dis 181, 106100 (2023). 10.1016/j.nbd.2023.106100

54 Burfeind, K. G. et al. The effects of noncoding aquaporin-4 single-nucleotide polymorphisms on cognition and functional progression of Alzheimer’s disease. Alzheimers Dement (N Y) 3, 348–359 (2017). 10.1016/j.trci.2017.05.001

55 Zeppenfeld, D. M. et al. Association of Perivascular Localization of Aquaporin-4 With Cognition and Alzheimer Disease in Aging Brains. JAMA Neurol 74, 91–99 (2017). 10.1001/jamaneurol.2016.4370

56 Simon, M. J. et al. Transcriptional network analysis of human astrocytic endfoot genes reveals region-specific associations with dementia status and tau pathology. Sci Rep 8, 12389 (2018). 10.1038/s41598-018-30779-x

57 Instrumentation, A. N. S. I. A. f. A. o. M. Vol. EC12:2000/(R)2020 (2020).

58 Ayllon, D., Seoane, F. & Gil-Pita, R. Cole equation and parameter estimation from electrical bioimpedance spectroscopy measurements - A comparative study. Annu Int Conf IEEE Eng Med Biol Soc 2009, 3779–3782 (2009). 10.1109/IEMBS.2009.5334494

59 De Lorenzo, A., Andreoli, A., Matthie, J. & Withers, P. Predicting body cell mass with bioimpedance by using theoretical methods: a technological review. J Appl Physiol (1985) 82, 1542–1558 (1997). 10.1152/jappl.1997.82.5.1542

60 Gabriel, S., Lau, R. W. & Gabriel, C. The dielectric properties of biological tissues: III. Parametric models for the dielectric spectrum of tissues. Phys Med Biol 41, 2271–2293 (1996). 10.1088/0031-9155/41/11/003

61 Welch, P. The use of the fast Fourier transform for the estimation of power spectra: A method based on time averaging over short, modified periodograms. IEEE Trans. Audio Electroacoust 15, 70–73 (1967).

62 Vallat, R. & Walker, M. P. An open-source, high-performance tool for automated sleep staging. Elife 10 (2021). 10.7554/eLife.70092

63 Quinn, A. J., Lopes-Dos-Santos, V., Dupret, D., Nobre, A. C. & Woolrich, M. W. EMD: Empirical Mode Decomposition and Hilbert-Huang Spectral Analyses in Python. J Open Source Softw 6 (2021). 10.21105/joss.02977

64 Heart rate variability: standards of measurement, physiological interpretation and clinical use. Task Force of the European Society of Cardiology and the North American Society of Pacing and Electrophysiology. Circulation 93, 1043-1065 (1996).

65 Janelidze, S. et al. Head-to-Head Comparison of 8 Plasma Amyloid-β 42/40 Assays in Alzheimer Disease. JAMA Neurol 78, 1375–1382 (2021). 10.1001/jamaneurol.2021.3180

66 Roberts, K. F. et al. Amyloid-β efflux from the central nervous system into the plasma. Ann Neurol 76, 837–844 (2014). 10.1002/ana.24270

67 Liu, H. et al. Acute sleep loss decreases CSF-to-blood clearance of Alzheimer’s disease biomarkers. Alzheimers Dement (2023). 10.1002/alz.12930

68 Loh, S., Lamond, N., Dorrian, J., Roach, G. & Dawson, D. The validity of psychomotor vigilance tasks of less than 10-minute duration. Behav Res Methods Instrum Comput 36, 339–346 (2004). 10.3758/bf03195580

69 Arsintescu, L. et al. Validation of a touchscreen psychomotor vigilance task. Accid Anal Prev 126, 173–176 (2019). 10.1016/j.aap.2017.11.041

70 Arbuthnott, K. & Frank, J. Trail making test, part B as a measure of executive control: validation using a set-switching paradigm. J Clin Exp Neuropsychol 22, 518–528 (2000). 10.1076/1380-3395(200008)22:4;1-0;FT518

71 Digit Span (DGS), <https://cambridgecognition.com/digit-span-dgs/> (

72 PVT Research Tool, <https://apps.apple.com/tr/app/pvt-research-tool/id1475726298> (

73 Smith, A. Symbol Digit Modalities Test. (Western Psychological Association, 1992).

74 Reitan, R. Trail Making Test: Manual for Administration and Scoring. (1992).

75 Diagnostics, C. N. <https://c2n.com/> (

76 Piantino, J. et al. Link between Mild Traumatic Brain Injury, Poor Sleep, and Magnetic Resonance Imaging: Visible Perivascular Spaces in Veterans. J Neurotrauma 38, 2391–2399 (2021). 10.1089/neu.2020.7447

77 Boespflug, E. L. et al. MR Imaging-based Multimodal Autoidentification of Perivascular Spaces (mMAPS): Automated Morphologic Segmentation of Enlarged Perivascular Spaces at Clinical Field Strength. Radiology 286, 632–642 (2018). 10.1148/radiol.2017170205

78 Levendovszky, S. R. et al. Preliminary cross-sectional investigations into the human glymphatic system using multiple novel non-contrast MRI methods. bioRxiv (2023). 10.1101/2023.08.28.555150

79 Ohene, Y. et al. Non-invasive MRI of brain clearance pathways using multiple echo time arterial spin labelling: an aquaporin-4 study. Neuroimage 188, 515–523 (2019). 10.1016/j.neuroimage.2018.12.026

80 Fultz, N. E. et al. Coupled electrophysiological, hemodynamic, and cerebrospinal fluid oscillations in human sleep. Science 366, 628–631 (2019). 10.1126/science.aax5440

81 Fong, Y., Huang, Y., Gilbert, P. B. & Permar, S. R. chngpt: threshold regression model estimation and inference. BMC Bioinformatics 18, 454 (2017). 10.1186/s12859-017-1863-x

82 Fong, Y., Di, C. & Permar, S. Change point testing in logistic regression models with interaction term. Stat Med 34, 1483–1494 (2015). 10.1002/sim.6419

83 Farokhian, F., Yang, C., Beheshti, I., Matsuda, H. & Wu, S. Age-Related Gray and White Matter Changes in Normal Adult Brains. Aging Dis 8, 899–909 (2017). 10.14336/AD.2017.0502

84 Ding, F. et al. Changes in the composition of brain interstitial ions control the sleep-wake cycle. Science 352, 550–555 (2016). 10.1126/science.aad4821

85 Sun, Y. & Sun, X. Exploring the interstitial system in the brain: the last mile of drug delivery. Rev Neurosci 32, 363–377 (2021). 10.1515/revneuro-2020-0057

86 Hotelling, H. New light on the correlation coefficient and its transforms. Journal of the Royal Statistical Society. Series B (Methodological) 15.2, 193–232 (1953).

